# Unbiased spectral cytometry immunome characterization predicts COVID-19 mRNA vaccine failure in older adults and patients with lymphoid malignancies

**DOI:** 10.1101/2023.04.10.23288350

**Authors:** Juan H-Vazquez, Paloma Cal-Sabater, Elisa Arribas-Rodríguez, Aida Fiz-López, Candido Perez-Segurado, Álvaro Martín-Muñoz, Ángel De Prado, Ignacio de la Fuente Graciani, Sonia Pérez González, Sara Gutiérrez, Pablo Tellería, Cristina Novoa, Silvia Rojo Rello, Antonio Garcia-Blesa, Rosa Sedano, Ana María Martínez García, Sonsoles Garcinuño Pérez, Marta Domínguez-Gil, Cristina Hernán García, Mª Mercedes Guerra, Eduardo Muñoz-Sánchez, Cristina Barragan-Pérez, Soraya Diez Morales, Oriana Casazza Donnarumma, Daniel Ramos Pollo, Natalia Santamarta Solla, Paula Mª Álvarez Manzanares, Sara Bravo, Cristina García Alonso, Ángel Tesedo Nieto, Elisabet Carmen López Moreno, María Esther Cabrera Sanz, Sara Borge Olmedo, Miguel de Paula Ortiz, Alberto Castellanos Asenjo, Jenifer Gay Alonso, José A. Garrote, Eduardo Arranz, José María Eiros, Fernando Rescalvo Santiago, Carolina Quevedo Villegas, Eduardo Tamayo, Antonio Orduña, Carlos Dueñas, María Jesús Peñarrubia, Sara Cuesta-Sancho, María Montoya, David Bernardo

## Abstract

COVID-19 affects the population unequally with a higher impact on aged and immunosuppressed people. Hence, we assessed the effect of SARS-CoV-2 vaccination in immune compromised patients (older adults and oncohematologic patients), compared with healthy counterparts. While the acquired humoral and cellular memory did not predict subsequent infection 18 months after full immunization, spectral and computational cytometry revealed several subsets within the CD8^+^ T-cells, B-cells, NK cells, monocytes and CD45RA^+^CCR7^-^ Tγδ cells differentially expressed in further infected and non-infected individuals not just following immunization, but also prior to that. Of note, up to 7 subsets were found within the CD45RA^+^CCR7^-^ Tγδ population with some of them being expanded and other decreased in subsequently infected individuals. Moreover, some of these subsets also predicted COVID-induced hospitalization in oncohematologic patients. Therefore, we hereby have identified several cellular subsets that, even before vaccination, strongly related to COVID-19 vulnerability as opposed to the acquisition of cellular and/or humoral memory following vaccination with SARS-CoV- 2 mRNA vaccines.

**SUMMARY:** An in depth and unbiased spectral cytometry characterization of the immune system before and after COVID-19 vaccination can predict not just subsequent PCR-confirmed infection, but also COVID-induced hospitalization in immune compromised patients.

## INTRODUCTION

COVID-19 has been shown to affect very unequally the population, with one of the main risk factors being having a depressed immune system. In this regard, multiple types of COVID-19 vaccines have been shown to be highly effective not just in preventing SARS-CoV-2 infection, but also in reducing post-infection symptoms. Indeed, although all of these vaccines induce systemic immune responses, little is known on their alterations from the immunological point of view. Hence, although the specific mechanisms of acquired humoral and cellular memory have been largely described, it has not been possible, up to date, to determine subsequent vaccine failure^1–4^.

In this framework, defining the efficacy of SARS-CoV-2 vaccines in frail populations is of paramount relevance for the design and implementation of future vaccine strategies. However, little is known regarding long-term immunity responses triggered by SARS-CoV-2 vaccines in old people and cancer patients after repeated booster doses^5, 6^. Hence, several studies have reported that patients with lymphoid cancers have been identified as being particularly at risk of inadequate antibody response to anti-SARS-CoV-2 vaccines^7^, particularly those with non-Hodgkin lymphoma (NHL) receiving B cell-depleting agents^8^. In a similar manner, immunosenescence is probably one of the determinants of progression to severe COVID-19^9^ as aging change both adaptive and innate immunity, resulting in increased susceptibility to infections and development of chronic inflammation^10^. Hence, vaccination has demonstrated to be the most effective tool against coronavirus disease 2019 (COVID-19). Nevertheless, despite the success of COVID-19 vaccines with their high efficacy in healthy populations, concerns about the efficacy and safety of COVID-19 vaccines in immunocompromised populations remain unresolved^11, 12^.

As a consequence, there is therefore an urgent need to better understand vaccine-induced immunogenicity in the context of heterogeneous host characteristics to improve protection for these patients. To that end, we hereby have performed a deep and unbiased characterization of the circulating immune system (or immunome) using top-of-the-art spectral cytometry in immune compromised patients (including older adults and oncohematologic patients), compared with healthy counterparts. Hence, our results have revealed that while the acquired humoral and cellular memory cannot predict subsequent infection, the unbiased analysis of the circulating immunome can predict subsequent infection even before vaccination took place.

## MATERIAL AND METHODS

### Biological samples

In order to address whether mRNA vaccines are equally effective in immune compromised patients, two cohorts of these patients were recruited, including older adults and patients with lymphoid cancer (oncohematologic), to be subsequent referred to a control cohort. In all cases, individuals with a previous diagnose (PCR-confirmed) of COVID-19 were excluded from the study. Older adults (2 males, 18 females) were recruited from the Orpea residential nursing home (Valladolid, Spain). All of them were vaccinated with BNT162b2 (Pfizer-BioNTech), with an average age of 88.1 years (all over 70 years old). Thirty-nine oncohematologic patients treated at the hematology department (Hospital Clínico Universitario de Valladolid, Valladolid, Spain) were also recruited. Seven of them had Chronic lymphocytic leukemia (CLL) or Follicular Lymphoma without active treatment, 10 had non-Hodgkin’s Lymphoma treated with Rituximab, 14 had CLL treated with Ibrutinib and 8 with Myeloma treated with Lenalidomide. With the exception of 4 patients vaccinated with BNT162b2 (Pfizer-BioNTech), the rest were vaccinated with mRNA-1273 (Moderna). This cohort shown homogeneity in terms of gender with an average age of 63.5 years. Finally, a total of 27 controls with not known inflammatory, autoimmune, or malignant diseases were recruited at the occupational risk prevention service (Hospital Clínico Universitario de Valladolid, Valladolid, Spain). Controls were selected to be matched for age and gender with the other cohorts. All of them were vaccinated with BNT162b2 (Pfizer-BioNTech). In all cases, a clinical follow-up of 18 months was performed in all individuals to further address (following vaccination) subsequent SARS-Cov-2 infection (PCR-confirmed). Further information about patient demographics and subsequent infection can be found in Table 1. Ethics approval was obtained by the local ethics committee from Valladolid Este (PI 21-2098).

**Table 1:** Patient demographics.

|  |  | Healthy adults (HA) | Older adults (OA) | Oncohematologic patients |  |  |  |
| --- | --- | --- | --- | --- | --- | --- | --- |
|  |  |  |  | Untreated (UP) | Lenalidomide-treated (LP) | Ibrutinib-treated (IP) | Rituximab-treated (RP) |
| n |  | 27 | 20 | 7 | 8 | 14 | 10 |
| Age |  | 59 (50-63) | 89 (86-94) | 66 (62-70) | 63.5 (56.25-74.75) | 66 (59-71) | 64 (60-73) |
| Sex (female) |  | 15 (55.55%) | 18 (90%) | 5 (71.42%) | 5 (62.5%) | 5 (45.45%) | 2 (20%) |
| Vaccine | BNT162b2 (Pfizer-BioNTech) | 27 | 20 | 1 | - | 2 | 1 |
|  | mRNA-1273 (Moderna) | - | - | 6 | 8 | 12 | 9 |
| Oncohematologic disease | Chronic lymphocytic leukemia (CLL) | - | - | 5 | - | 14 | - |
|  | Follicular Lymphoma (FL) | - | - | 2 | - | - | - |
|  | non-Hodgkin's Lymphoma | - | - | - | - | - | 10 |
|  | Myeloma | - | - | - | 8 | - | - |
| Treatment | No treatment | - | - | 7 | - | - | - |
|  | Lenalidomide | - | - | - | 8 | - | - |
|  | Ibrutinib | - | - | - | - | 14 | - |
|  | Rituximab | - | - | - | - | - | 10 |
| SARS-CoV-2 PCR+ 18 months after immunization |  | 7 out of 23 | 3 out of 20 | 1 out of 7 | 2 out of 7 | 9 out of 13 | 2 out of 7 |
| COVID-19 Hospitalization 18 months after immunization |  | 0 out of 7 | 0 out of 3 | 0 out of 1 | 0 out of 2 | 3 out of 9 | 0 out of 1 |

### Biological samples

Blood samples from all individuals were obtained before the first dose, and 3 months following the full immunization (i.e., 3 months after the second dose). Blood was collected in LH Lithium Heparin separator tubes. Subsequently, peripheral blood mononuclear cells (PBMCs) were isolated using Cytiva Ficoll-Paque™ PLUS (Cytiva 17-1440-03). Blood was slowly poured into a centrifuge tube with Ficoll-Paque™ without mixing (3 ml of Ficoll for 5ml of blood) and centrifuged at 800g for 30min at 4°C (Fisherbrand™ GT2) with acceleration set to maximum and deceleration to minimum. PBMCs were collected from the interface between the Ficoll-Paque™ and plasma layers. PBMCs were centrifuged again in RPMI at 400g for 5 min at 4°C to wash them. The resulting pellet was suspended in freezing medium (90% Fetal Bovine Serum (FBS) +10% dimethyl sulfoxide (DMSO)) to cyopreserve the cells in liquid nitrogen until used.

### Humoral memory

The determination of specific IgG and IgA antibodies against the receptor binding domain of the S protein (Spike) of SARS-CoV-2 was performed by electrochemiluminescence immunoassay (Elecsys Anti-SARS-CoV-2 S, Roche Diagnostics, Mannheim, Germany). The results are expressed in Binding Antibody Units (BAU). In addition, the presence of specific antibodies against SARS-CoV-2 N antigen was studied by enzyme-linked immunosorbent assay (ELISA) (COVID-19 ELISA IgG, Vircell Microbiologists, Santa Fe, Granada, Spain).

### Cellular memory

The magnitude and kinetics of the cellular response to SARS-CoV-2 was tested by an ex vivo IFN-γ ELISpot assay. To that end, PBMCs (both before vaccination and 3 months following full immunization) were thawed in sterile tubes with 10 mL of RPMI 1640 without L-Glutamine (Gibco) and centrifuged at 400g 4°C 5 min. After removal of the supernatant, 1 mL of AIM-V Serum free medium with L-Glutamine, 50 μg/ml Streptomycin Sulfate and 10 μg/ml Gentamycin Sulfate (Gibco) culture medium was added to count the cells in a counting chamber BLAUBRAND® Neubauer improved in the presence of Trypan Blue. Cells were cultured in duplicate (100,000 viable cells in 200 μl of AIM-V medium) in 96 U-bottom culture plates for a total period 48h in the presence of 2 μg/ml of a pool of SARS-CoV-2 spicule S1 domain peptides (Mabtech). As a positive control, total PBMCs were stimulated with a polyclonal stimulus of anti-CD3 and anti-CD28 (Mabtech), at concentrations of 0.2 and 0.02 μg/ml respectively while unstimulated cells provided a negative control. Following culture, secreted IFN-γ was detected by adding 1 μg/ml anti-IFN-γ mAb (7-B6-1-ALP, Mabtech) and subsequent 2h incubation in the dark. Plates were revealed using BCIP/NBT-plus, according to the manufacturer’s instructions. The results were obtained as spot forming units (SFU).

### Antibody staining and spectral cytometry acquisition

After thawing, and in parallel to determining cellular memory, two million PBMCs were stained with monoclonal antibodies (Supplementary Table 1) to be subsequently characterized by spectral cytometry (CyTek Aurora 5-laser) following the OMIP-069 protocol and analysis panel with slightly variations^13^.

Briefly, before staining the PBMCs, Live/Dead fixable blue dead cell stain kit (Molecular Probes, Thermo Fisher Scientific) was added to exclude dead cells from the analysis. Brilliant Stain Buffer and True-Stain Monocyte Blocker were also added prior to staining with the antibodies to obtain optimal fluorescence of the desired cells. The PBMCs were washed with FACS buffer (500mL PBS +1 0mL filtered FCS + 0.1g NaN3 + 2.5mL sterile EDTA) and incubated in the dark at room temperature during the staining process. Finally, cells were fixed with 0.8% paraformaldehyde in FACS buffer in the dark for 10 min and washed with FACS buffer to store at 4°C. Cells were acquired within 48 hours in a 5-laser spectral cytometer (Aurora, Cytek).

### Computational cytometry analysis and statistical analysis

The OMIQ Data Science platform (© Omiq, Inc. 2022) was used following transformation of the data setting the scale, parameters and cofactors as suggested by the platform. Data cleaning algorithm FlowAI was applied in order to remove outlier events in spectral cytometry data files due to abnormal flow behaviors resulting from clogs and other common technical problems. Subsequently, a manual discard was performed to eliminate cell debris and doublets and to select viable leukocytes (CD45^+^ cells) which were used for subsequent analysis.

Due to the large amount of data obtained with this panel, it is not advisable to examine the results exclusively through traditional manual identification due to their subjectivity. Therefore, an unsupervised approach applying Uniform Manifold Approximation and Projection (UMAP) algorithm was used for the exploratory analysis. Briefly, this algorithm uses a non-linear method based on graphs constructed to represent information in multiple dimensions, and then reconstructs the results into a two-dimensional map, preserving the multidimensional structure. In this way, the algorithm finds similarities between cells in all dimensions. These dimensions are, in this case, the intensity of the markers that they express. The algorithm returns a two-dimensional map where the proximity of cells reflects their distances in multidimensional space, such that cells with similar patterns of expression were located very close. This distance/similarity relationship is respected within and between each group or islet. A prior subsampling or random selection of events was performed until the total of 4 million events was reached, so that each cohort was equally represented.

Subsequently, the FlowSOM algorithm was used to find similar cell subsets and separate them into groups in an unsupervised manner. This algorithm analyzes the expression of all the selected markers in each of the cells of each sample and then groups them into metaclusters according to their expression level. In this way, it not only allows for the visualization of cells in typical biological groupings, but also for the detection of new or unexpected subsets. However, this algorithm only displayed metaclusters that would represent the large subsets of the immune system present in the sample. The visual representation of the two algorithms, allows to further to subdivide these metaclusters into clusters representing more accurately all the phenotypic and functional subsets of the human immunome. A clustered heatmap was created using the clusters obtained in the previous point. This heatmap graphically represents the level of expression of each phenotypic marker, into each cluster. Dendrograms grouped clusters and phenotypic markers associated by similarity (distance). This approach permits to identify the immune subsets represented by each cluster based on the expression levels of their markers. In this way, if a specific cluster is associated with a condition under study, its phenotype could be elucidated to identify it using classical supervised approaches where it would otherwise have gone undetected. Finally, the refine results of FlowSOM algorithm were mapped on the UMAP in order to observe their distribution.

### Statistical analyses

For the computational cytometry data, Volcano plots were constructed by comparing differences in clusters between all studied conditions. This type of graph uses the total events into each cluster combining a measure of statistical significance from a statistical test (e.g., a p-value from an ANOVA model) with the magnitude of the change, enabling quick visual identification of clusters that display large change, that are also statistically significant. Once the clusters showing significant differences were identified, a validation of the data was performed by classical hierarchical analysis. Using a modified gating strategy of OMIP-69 panel, we obtained the percentages within the total viable leukocytes fraction (CD45⁺) of those clusters that stood out in the previous analysis, and then performed the statistical analysis using GraphPad Prism9. Quantitative variables were expressed as mean and standard deviation as followed a normal distribution, and parametric t-student test were used. One-Way ANOVA, Fisher test and t-test comparisons were also applied as detailed in the Figure Legends. In all cases, a p-value under 0.05 were considered statistically significant.

## RESULTS

### Cellular immunome identification

UMAP analysis of the 162 analyzed samples (27 healthy adults, 20 older adults and 39 oncohematologic patients at 2 different time points -before and after vaccination-) identified four major continents and three smaller islands (Figure 1A). The relative expression of each marker on the UMAP is shown in Figure 1B revealing that the main continent on the left represent cytotoxic T-cells together with Tγδ, while the main continent on the right is made of helper T-cells. In a similar manner, the smaller island on the bottom is mainly composed of B-cells and the two islands in the middle represent monocytes. NK cells are in the top island together with innate lymphoid cells (ILCs) while the small top left island is composed by mixed myeloid antigen presenting cells.

**Figure 1.**
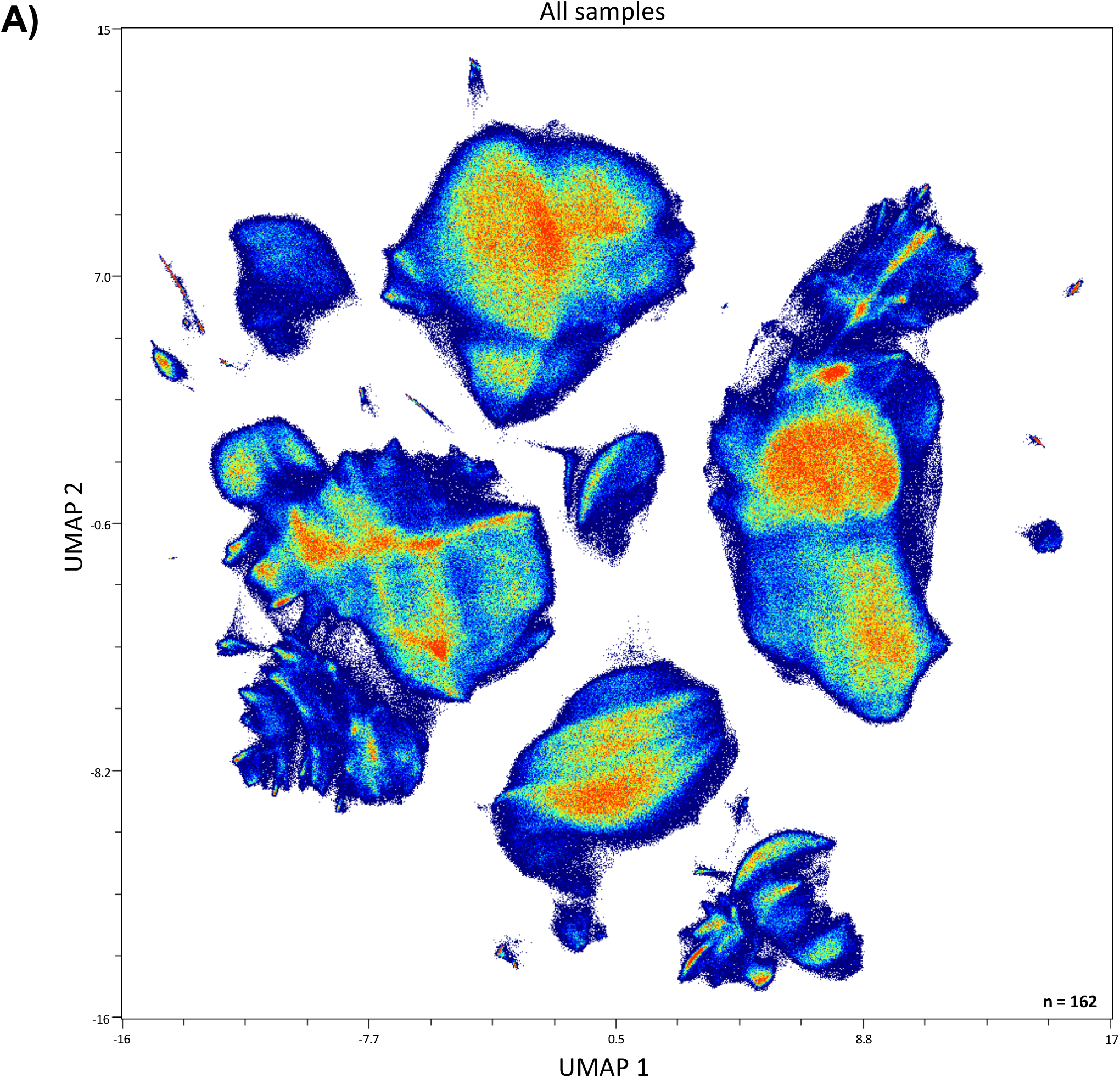
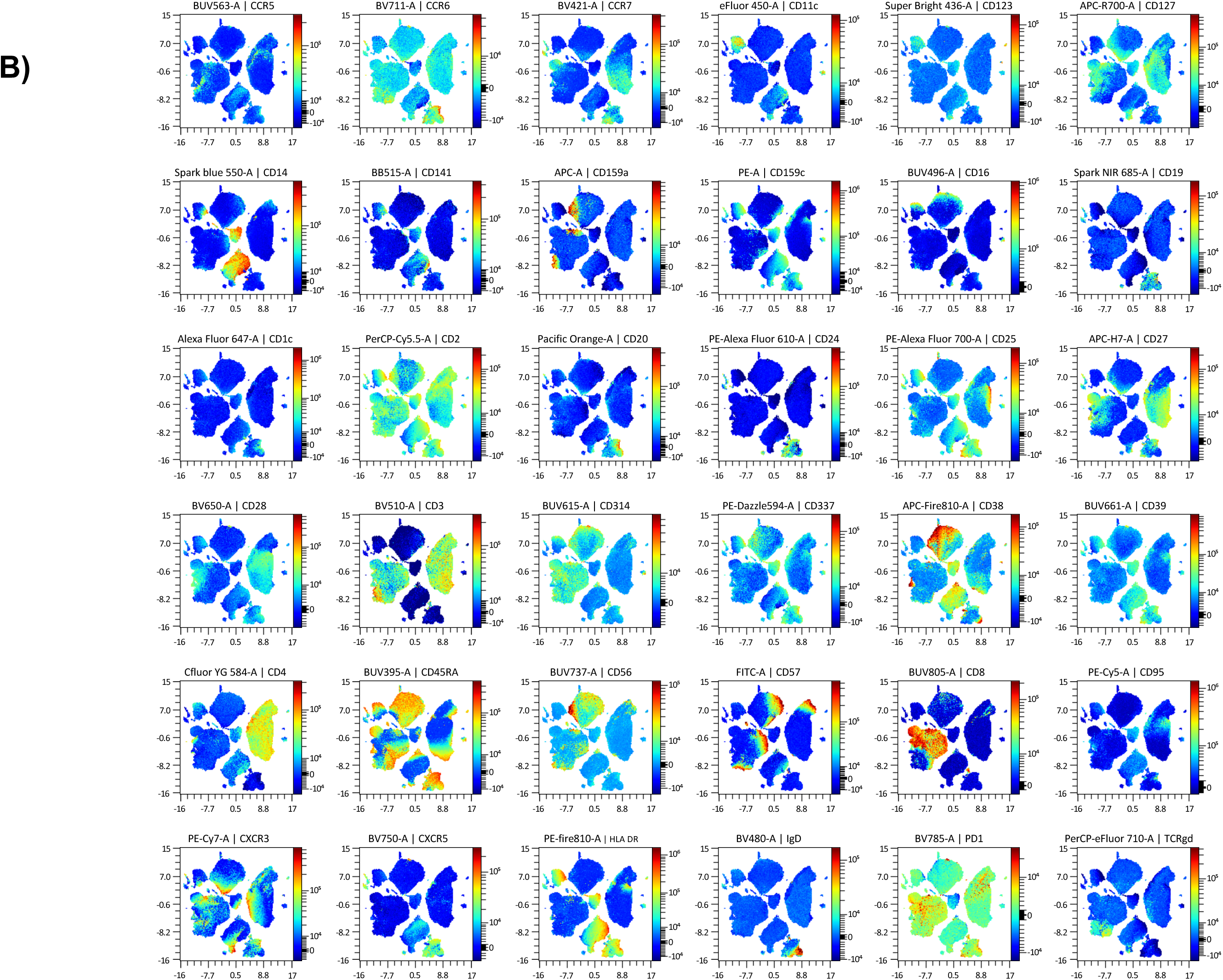
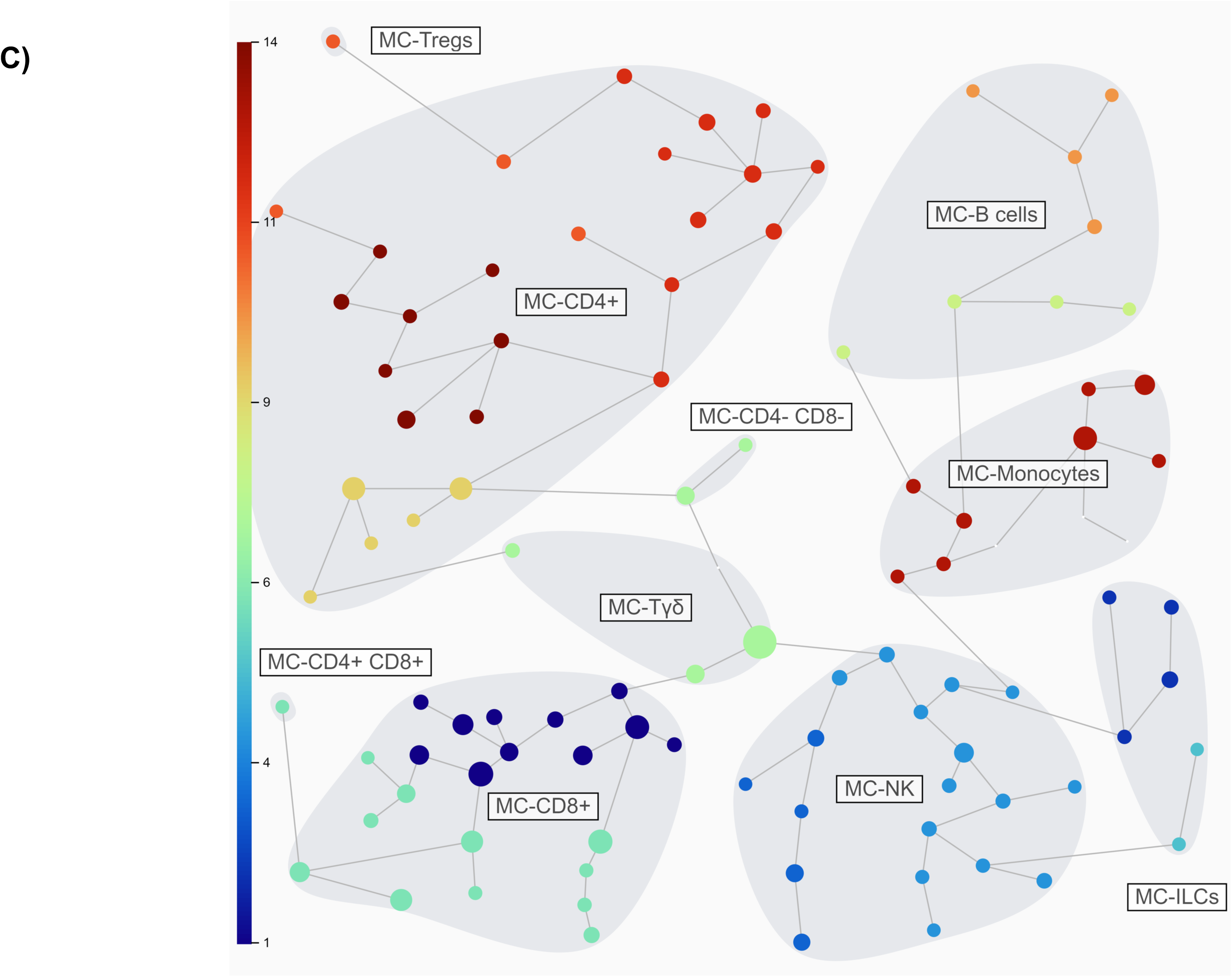
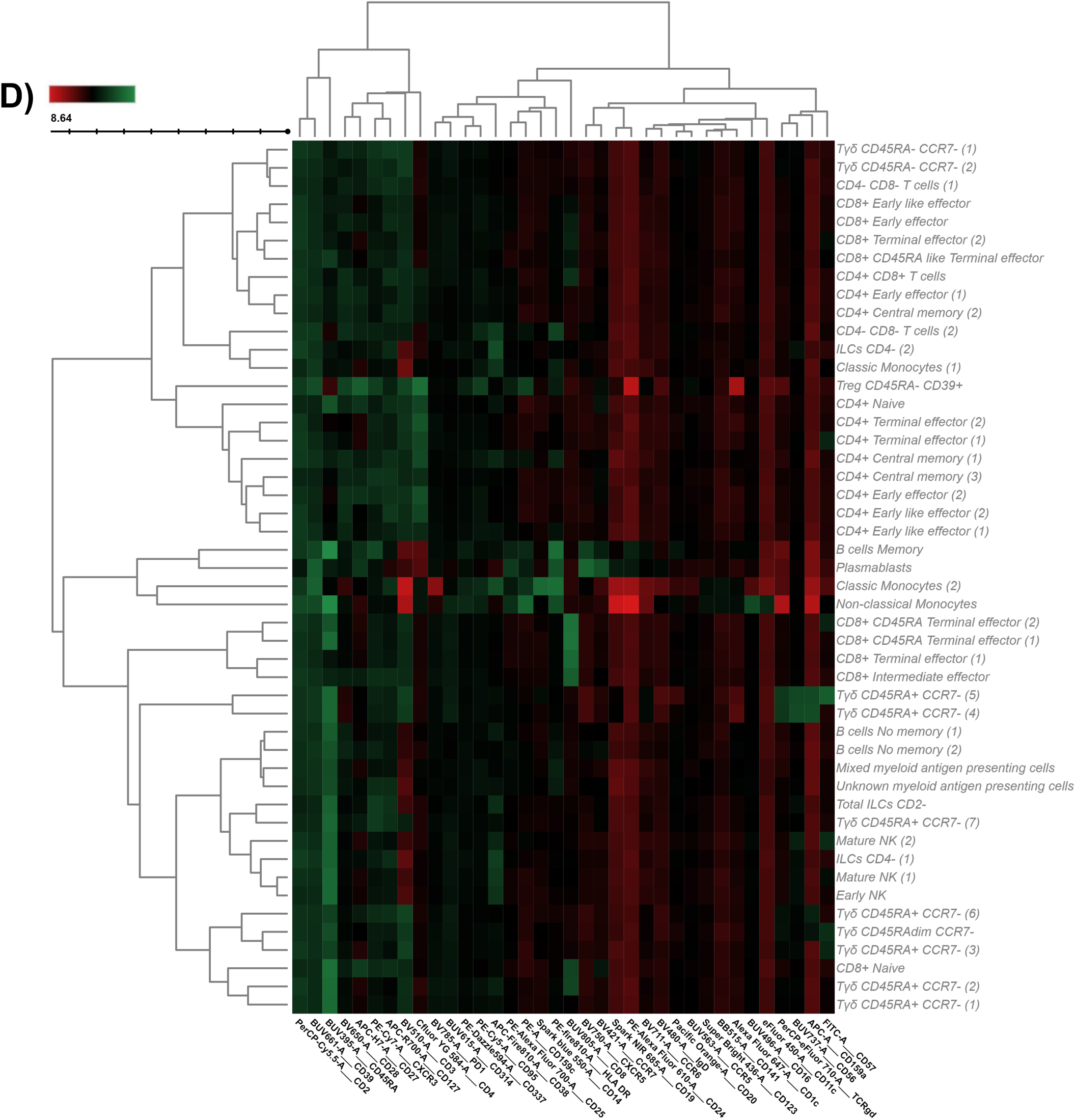
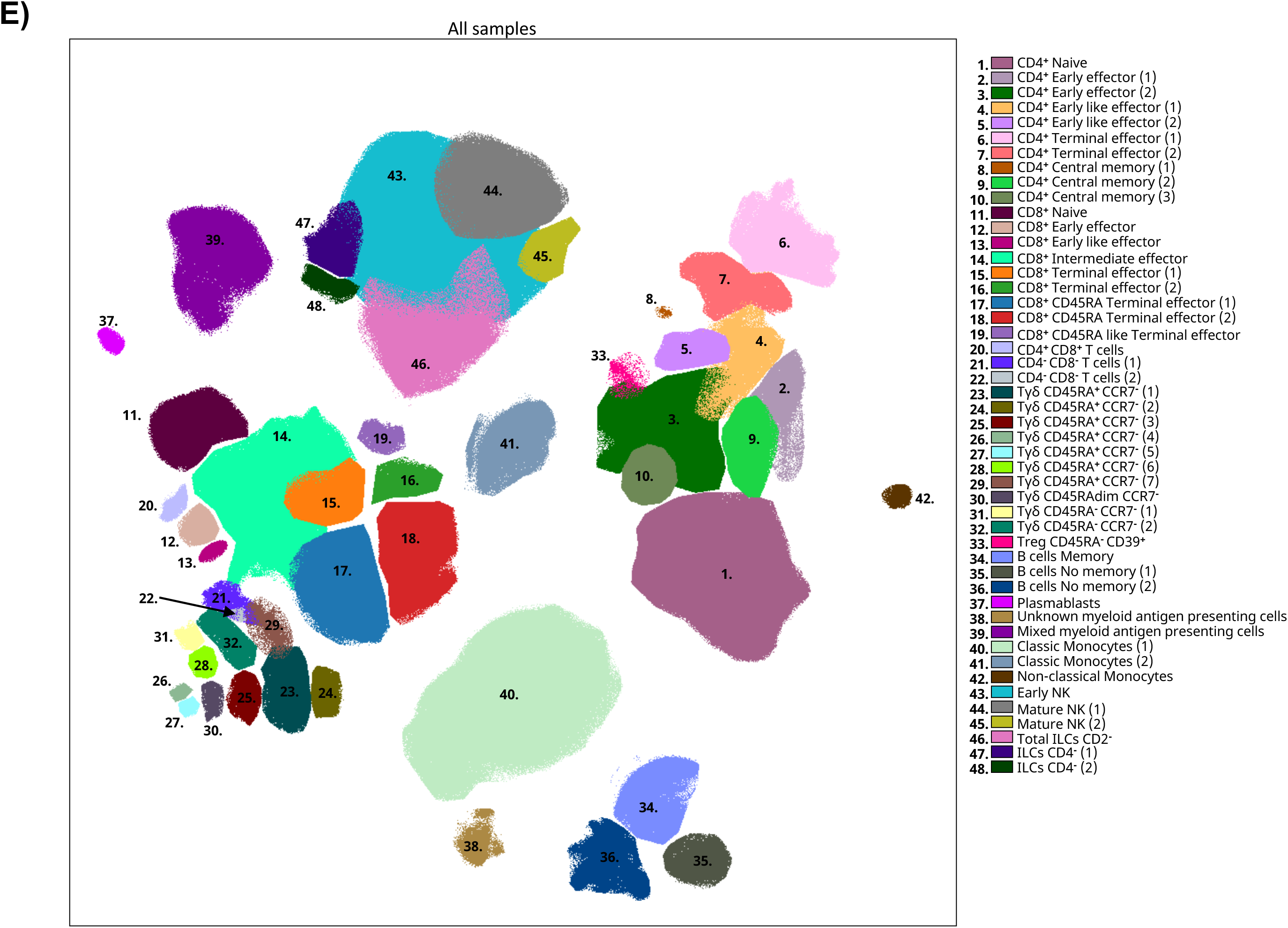
Immunome characterization following UMAP analysis. **A)** UMAP analysis was performed within total singlet viable CD45^+^ cells from all samples (n=162). Subsequent down-sampling to a total of 4 million events was performed being each cohort equally represented. Surface expression intensities of all remaining 36 analysed markers is shown in **B)** by a color code based on the intensity where red represent higher expression and blue represent lower expression. **C)** FlowSOM analysis of total singlet viable CD45^+^ cells identified the main metaclusters on dataset: B-cells, NK cells, innate lymphoid cells (ILCs), Tγδ, regulatory T-cells, CD4^+^ T-cells, CD8^+^ T- cells, CD4^+^CD8^+^ T-cells and CD4^-^CD8^-^ T-cells. **D)** Heatmap displaying the intensity levels of each marker within the 48 identified clusters. A color code based on the expression intensity where green represent higher expression and the transition to red represent lower expression was used. A dendrogram was generated by unsupervised hierarchical clustering. **E)** All 48 identified clusters were overlaid on the UMAP projection. Each identified cluster is tagged by a specific color and number as shown in the legend.

To further refine our analysis, FlowSOM algorithm was used to find similar cell subsets and separate them into metaclusters in an unsupervised manner (Figure 1C). A total of 48 clusters were identified according to the expression of the surface markers as shown in the heatmap (Figure 1D). Table 2 shows an in-depth characterization of the phenotype of all clusters, which allowed the identification of 46 of them, since clusters 38 and 39 could not be clearly identified. In the case of cluster 38, although is close to monocytes in the UMAP plot, there is no expression of CD14 or CD16. Hence, cluster 38 could be conventional dendritic cells (cDC) as they are negative for almost everything except HLADR, although they are also CD11c^-^ so their true nature remains elusive. Cluster 39 is made of intermediate (CD14^+^CD16^+^) and non-classical (CD14^+^CD16^-^) monocytes together with other HLA-DR^+^CD11c^+^ that could likely resemble cDC.

**Table 2:**
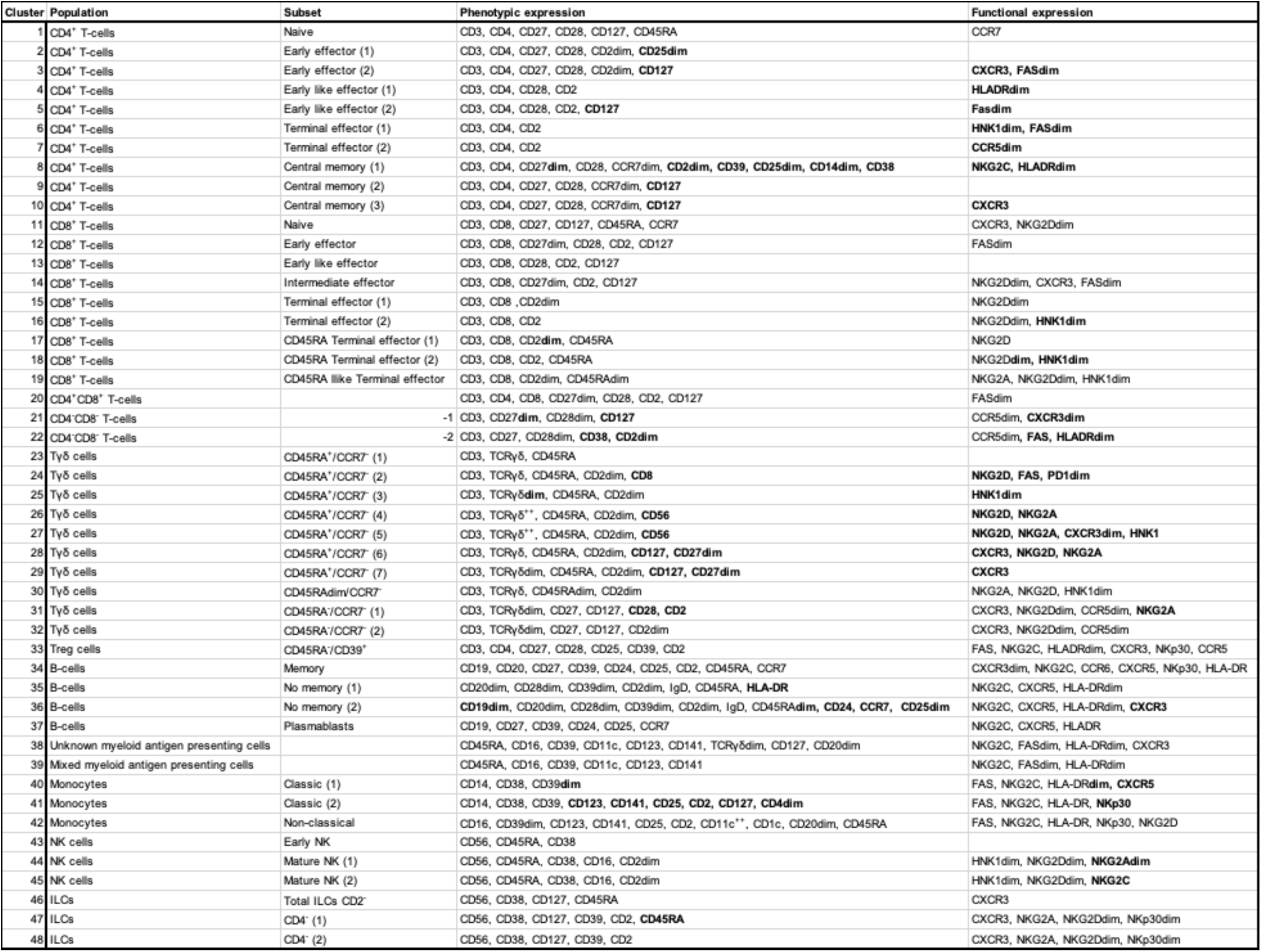
Cell cluster identification. For each for the 48 identified FlowSOM clusters (C) the cell population to which it belongs is shown, together with the specific subset, phenotype and expression of functional markers. Markers highlighted in bold denote differential expression within the same population.

Of note, the same cell population can be divided into further subsets as shown in Table 2 based on the expression of several surface markers. For instance, Tγδ CD45RA**^+^**CCR7**^-^**cells can be divided into 7 different subsets based on the surface markers expression of lineage CD56, CD127, CD27, CD8 together with NKG2D, FAS, CCR6 among others. Finally, all clusters were further uploaded into the UMAP (Figure 1E) to determine not just how the relate one to each other, but also to display their pseudoevolution.

In order to further validate these findings, the hierarchical or classical gating strategy to identify different immune subsets is shown in Figure 2A, with a particular input on T-cells being shown in Figure 2B. Given that some of the identified clusters can be found within the same subset (Table 2 and Figure 1D), Figure 2C displays the required gating strategy to identify these clusters while Figure 2D shown the identification of the 7 Tγδ CD45RA^+^CCR7^-^ subsets.

**Figure 2.**
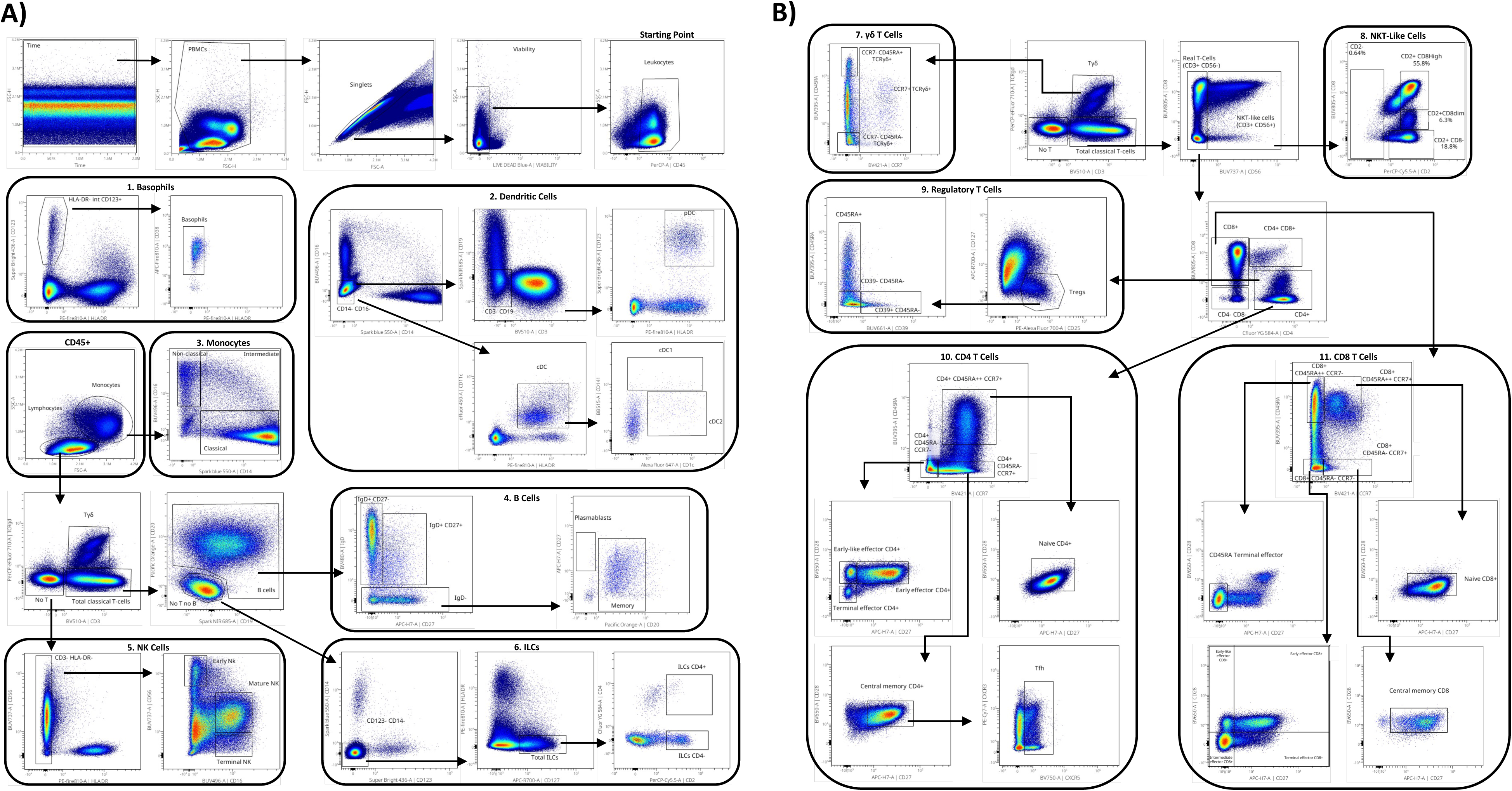
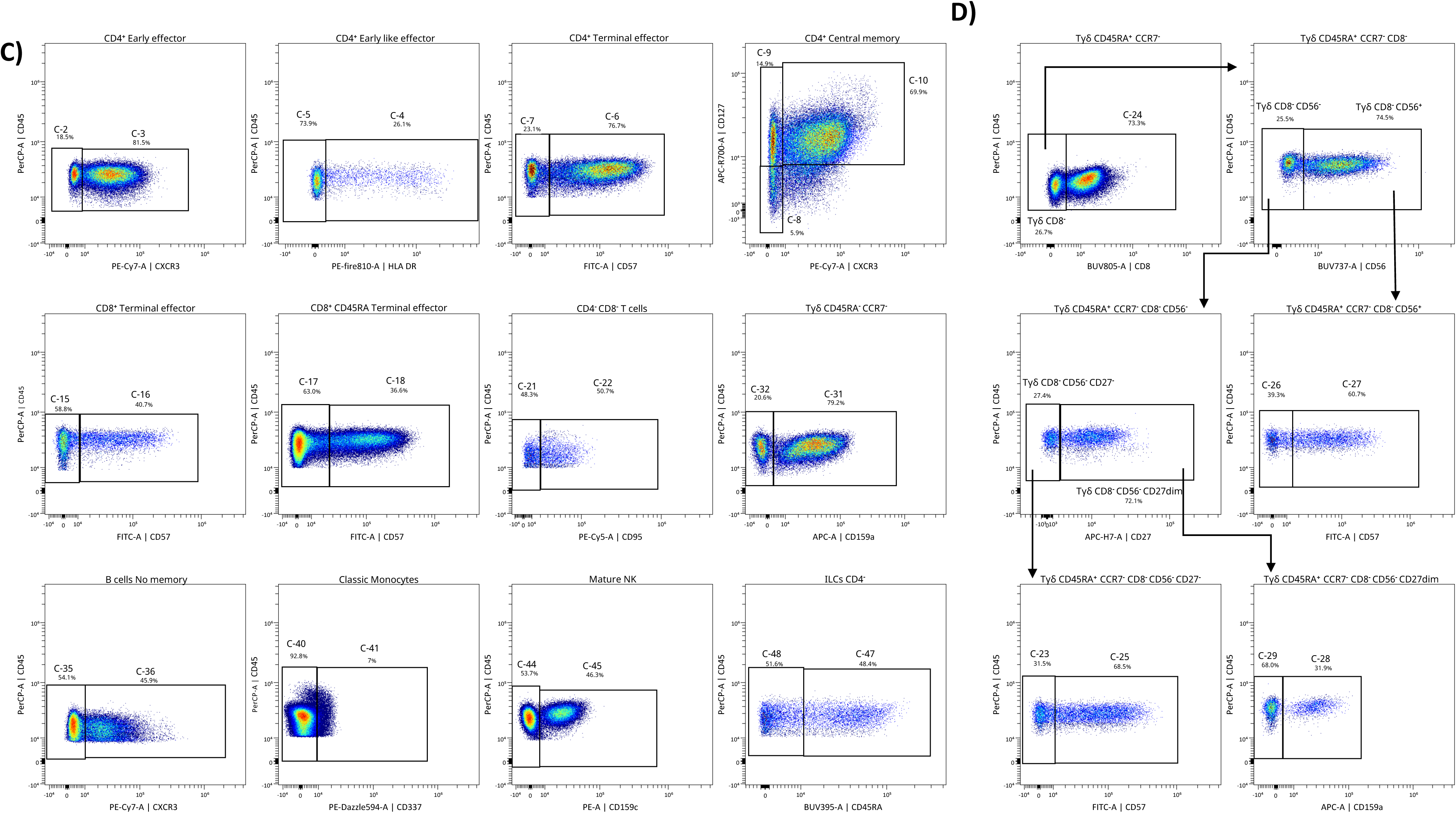
Hierarchical cell subsets identification. **A)** Representative gating strategy used to identify within total peripheral blood mononuclear cells (PBMC) the main cellular populations. Arrows are used to visualize the relationships across plots, and numbers are used to call attention to populations described here. After doublets and dead cells were excluded, basophils (1) were delineated as CD45^+^CD123^+^HLA-DR^-^. Dendritic cells (DCs, 2) were identified within CD14^−^CD16^−^ as CD3^-^CD19^-^CD123^+^HLA-DR^+^ (plasmacytoid DCs, -pDC-) and CD3^-^ CD19^-^CD11c^+^HLA-DR^+^ (classical or conventional DC -cDC-). cDC were further divided into type 1 (CD141^+^, -cDC1-) and type 2 (CD1c^+^, -cDC2-). Monocytes were gated based on FSC-A/SSC-A properties and further classified into non-classical (CD14^−^CD16^+^), intermediate (CD14^+^CD16^+/low^), and classical (CD14^+^CD16^-^). From the lymphogate, B-cells (4) were gated out of the CD3^−^TCRγδ^−^ as CD19^+^ and/or CD20^+^ cells. B-cells were further gated as IgD^+^CD27^−^, IgD^+^CD27^+^, or IgD^−^CD27^+/−^; the IgD^−^CD27^+/−^ subset was divided into plasmablasts or IgD^−^ memory B cells based on CD20 expression and CD27. NK cells (5) were defined within the lymphogate as CD3^−^TCRγδ^−^HLA-DR^−^ and classified as early NK (CD56^+^CD16^−^), mature NK (CD56^+^CD16^+^), and terminal NK (CD56^−^CD16^+^) cells. Innate lymphoid cells (ILCs, 6) were gated within the lymphogate as CD3^−^CD19^−^CD20^−^CD14^−^CD123^−^CD127^+^CD2^+^ and further divided into subsets based on the expression of CD4. **B)** T-cell were identified within the lymphogate based on the expression of CD3. Total Tγδ cells (7) were identified as CD3^+^TCRγδ^+^ and subsetted based on the expression of CD45RA and CCR7. Total NKT-like cells (8) were identified in the CD3^+^TCRγδ^-^ compartment as CD56^+^. The inclusion of CD2 and CD8 allowed further classification of the NKT-like cells. Real T-cells were defined as CD3^+^TCRγδ^-^ CD56^-^ and further divided into CD4^+^, CD8^+^, CD4^+^CD8^+^ and CD4^−^CD8^−^ T-cells. Regulatory T-cells (T_regs_, 9) were identified within total CD4^+^ T-cells as CD127^lo/-^CD25^hi^. CD39 and CD45RA were used to further classify them. Within total CD4^+^ T-cells (10) and CD8^+^ T-cells (11), CCR7, CD45RA, CD27, and CD28 were further used to divide them into different T-cell phenotypes as shown in the figure. **C)** Hierarchical gating was also used to identify the different clusters identified in Figure 1 and Table 2 within the subset described on the top of each plot. CD4 early effector were classified by CXCR3 expression as CD4 early effector (1) (CXCR3−), and CD4 early effector (2) (CXCR3^+^). CD4 early like effector were classified by HLA-DR expression as CD4 early like effector (1) (HLA-DR^+^), and CD4 early like effector (2) (HLA-DR^−^). CD4 terminal effector were classified by CD57 expression as CD4 terminal effector (1) (CD57^+^), and CD4 terminal effector (2) (CD57^−^). CD4 central memory cells were divided in CD127^+^CXCR3^+^ (CD4 central memory (3)), CD127^+^CXCR3^-^ (CD4 central memory (2)) and CD127^-^CXCR3^-^ (CD4 central memory (1)). CD8^+^ terminal effector were classified by CD57 expression as CD8^+^ terminal effector (1) (CD57^−^), and CD8^+^ terminal effector (2) (CD57^+^). CD8^+^CD45RA^+^ terminal effector were classified by CD57 expression as CD8^+^CD45RA^+^ terminal effector (1) (CD57^−^), and CD8^+^CD45RA^+^ terminal effector (2) (CD57^+^). CD4^-^CD8^-^ T-cells were classified by CD95 expression as CD4^-^CD8^-^ T cells (1) (CD95^−^), and CD4^-^CD8^-^ T-cells (2) (CD95^+^). Tγδ CD45RA^-^CCR7^-^ were classified by CD159a expression as Tγδ CD45RA^-^CCR7^-^ (1) (CD159a^+^), and Tγδ CD45RA^-^CCR7^-^ (2) (CD159a−). No memory B-cells were classified by CXCR3 expression as No memory B-cells (1) (CXCR3^−^), and No memory B-cells (2) (CXCR3^+^). Classic monocytes were classified by CD337 expression as Classic monocytes (1) (CD337^−^), and Classic monocytes (2) (CD337^+^). Mature NK were classified by CD159c expression as Mature NK (1) (CD159c^−^), and Mature NK (2) (CD159c^+^). ILCs CD4^-^ were classified by CD45RA expression as ILCs CD4^-^ (1) (CD45RA^+^), and ILCs CD4^-^ (2) (CD45RA^−^). **D)** Given the large number of identified Tγδ CD45RA^+^CCR7**^-^** clusters, the gating strategy to identify them is further shown. This population was divided in CD8^+^ (Tγδ CD45RA^+^CCR7^-^ (2)) and CD8^-^ subsets. The inclusion of CD56 enables further classification of Tγδ CD45RA^+^CCR7^-^CD8^-^. Tγδ CD45RA^+^CCR7^-^CD8^-^CD56^+^ were then divided by CD57 expression as Tγδ CD45RA^+^CCR7^-^ (4) (CD57^-^) and Tγδ CD45RA^+^CCR7^-^ (5) (CD57^+^). CD27 were used for further classification of Tγδ CD45RA^+^CCR7^-^CD8^-^CD56^-^. Tγδ CD45RA^+^CCR7^-^CD8^-^CD56^-^CD27^-^ were classified by CD57 expression as Tγδ CD45RA^+^CCR7^-^ (1) (CD57^-^) and Tγδ CD45RA^+^CCR7^-^ (3) (CD57^+^). Finally, CD159a were used to divided Tγδ CD45RA^+^CCR7^-^CD8^-^CD56^-^ CD27^dim^ on Tγδ CD45RA^+^CCR7^-^ (6) (CD159a^+^) and Tγδ CD45RA^+^CCR7^-^ (7) (CD159a^-^).

### In deep immune characterization of the cohorts at baseline

Having described the global leukocyte subsets composition (Figure 1), we next addressed the differences between cohorts at baseline (Figure 3A). Hence, the UMAP plot revealed a deficit of classical monocytes in older adults as well as in lenalidomide and ibrutinib treated oncohematologic patients. In a similar manner, and as expected, rituximab-treated-patients display a lack of B-cells. Differences according to surface markers expression within the cohorts are shown in the heatmap (Figure 3B).

**Figure 3:**
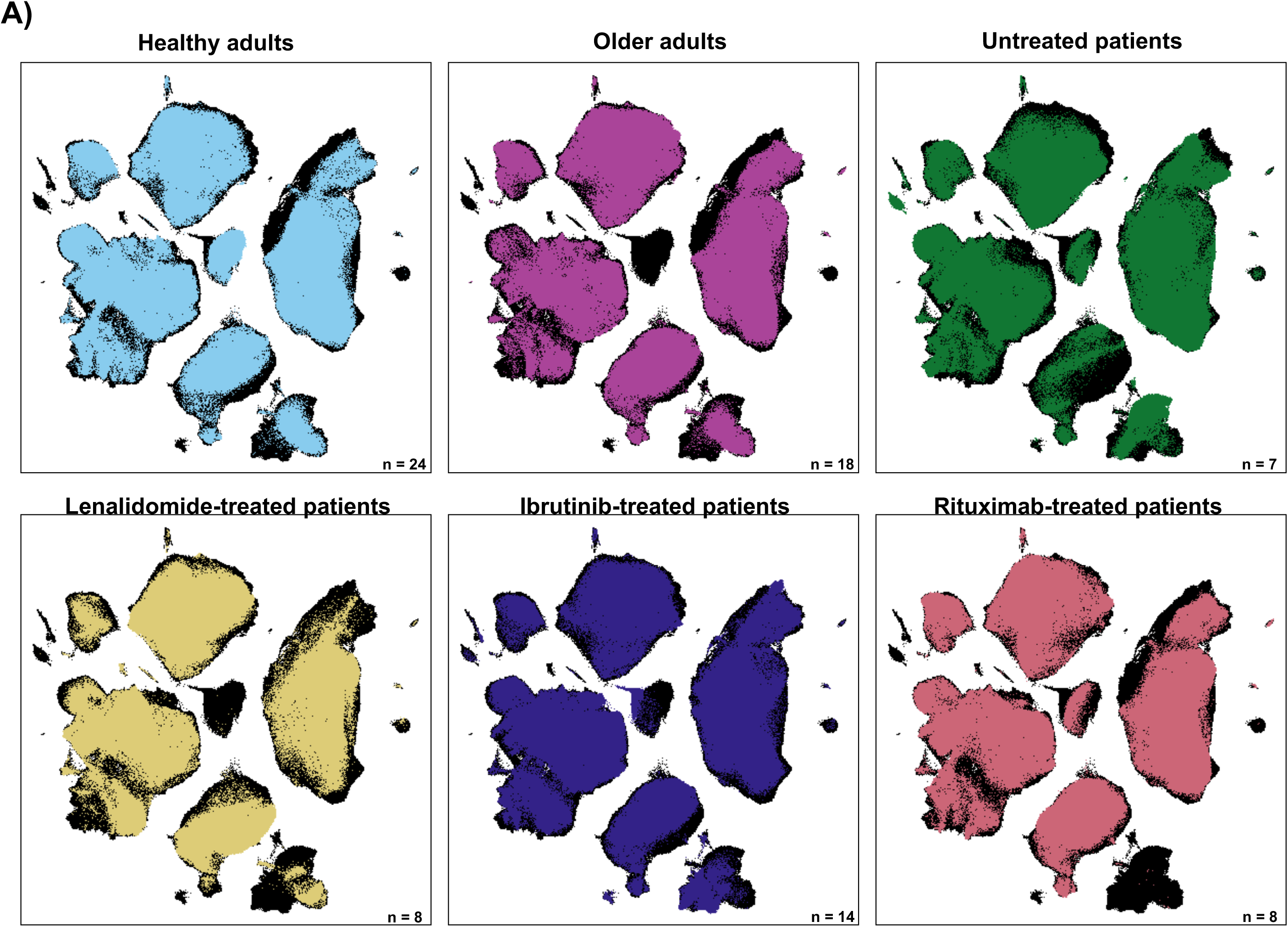
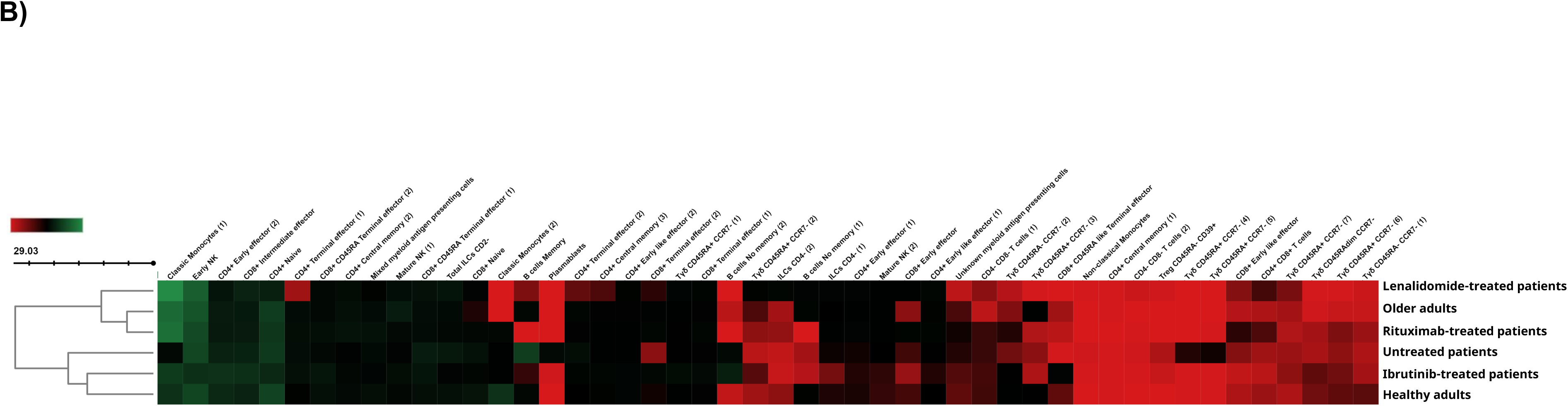
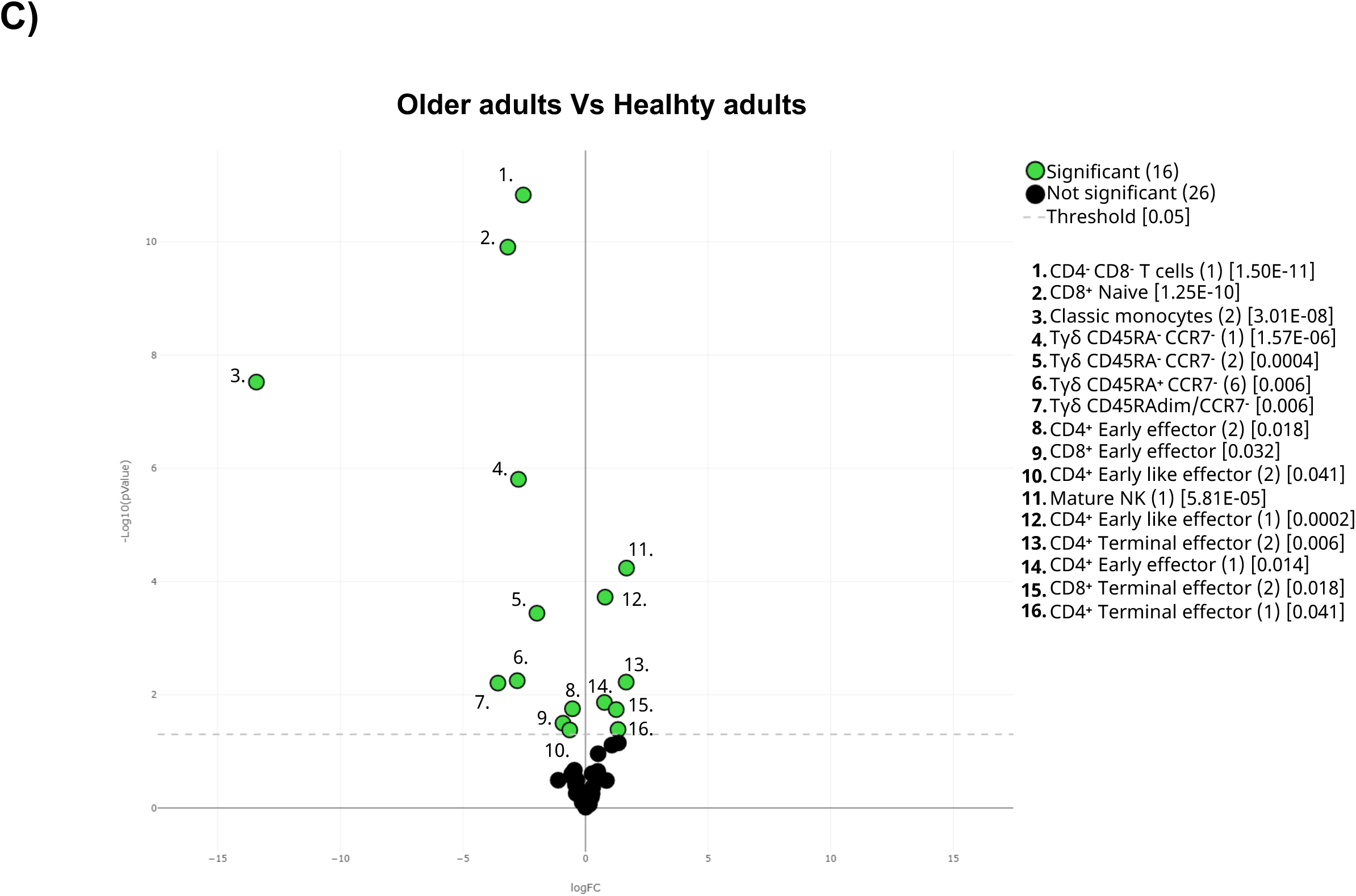
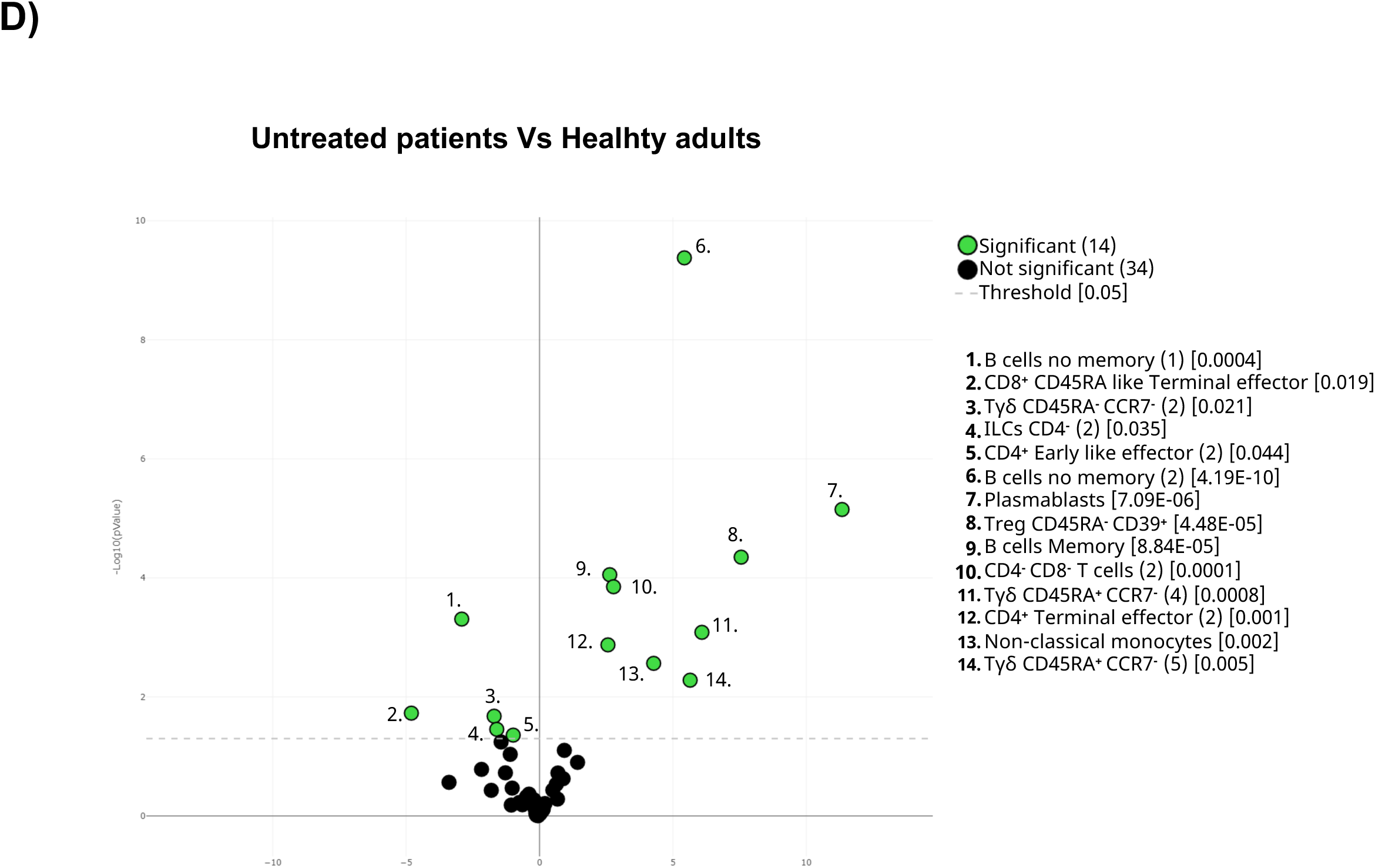
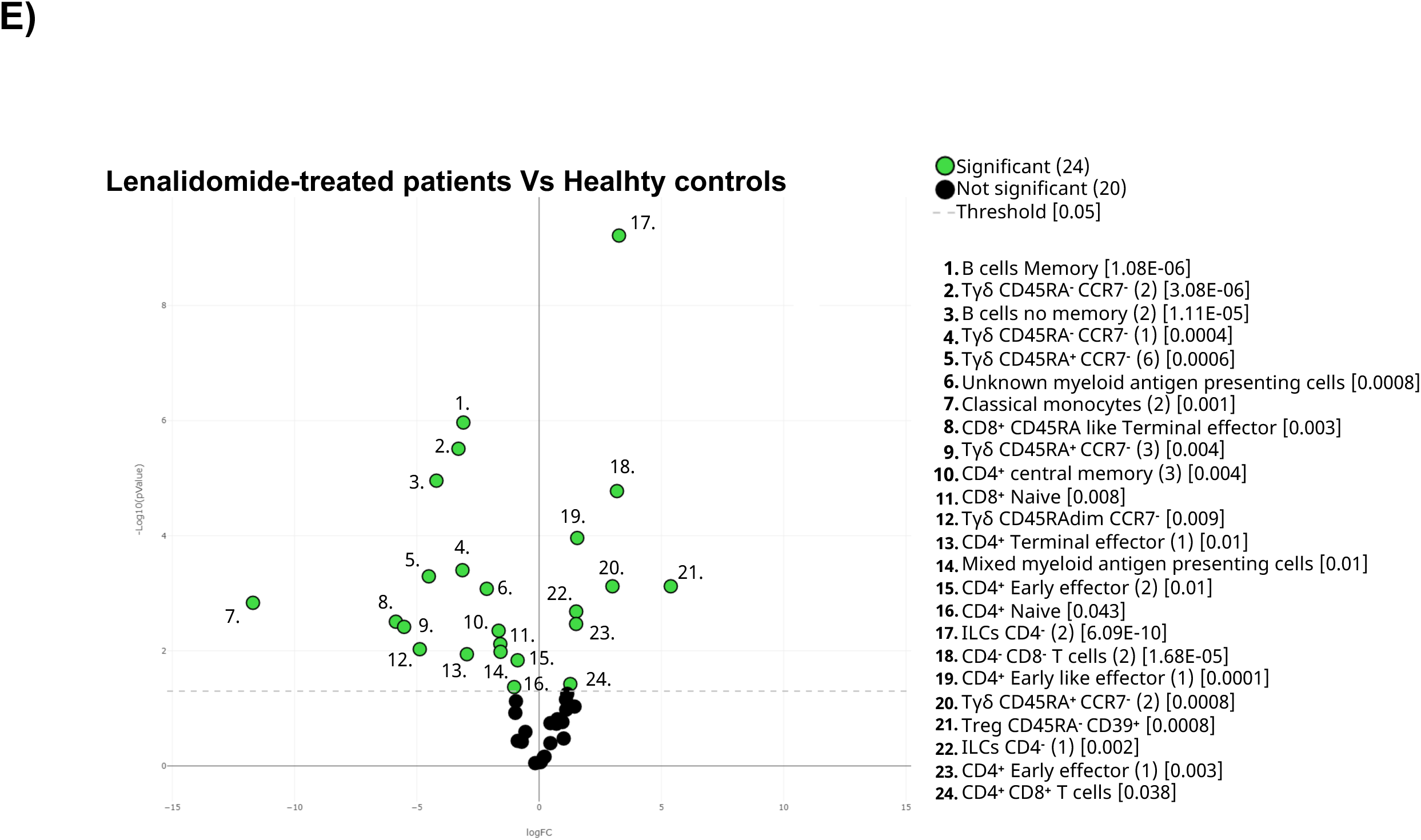
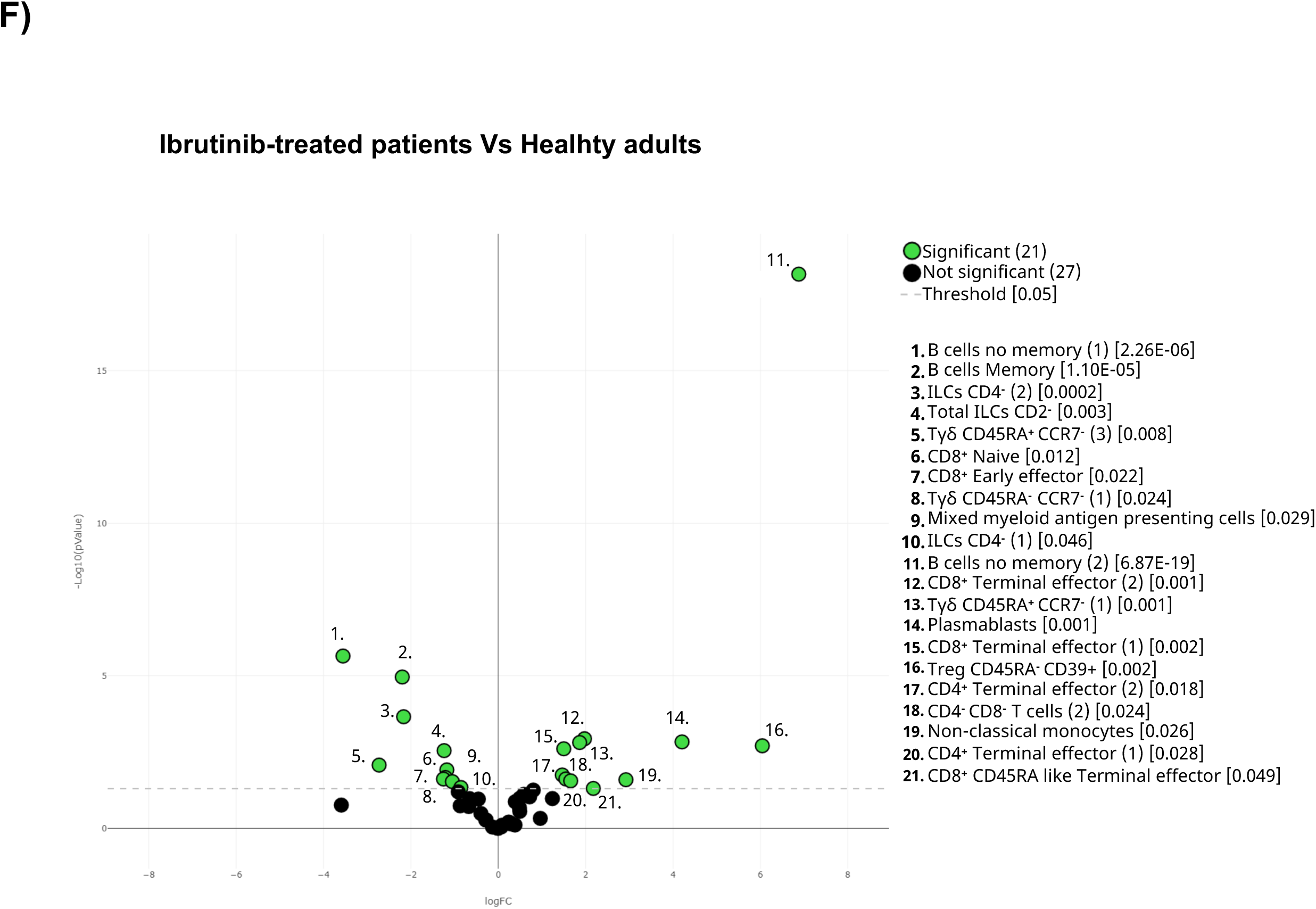
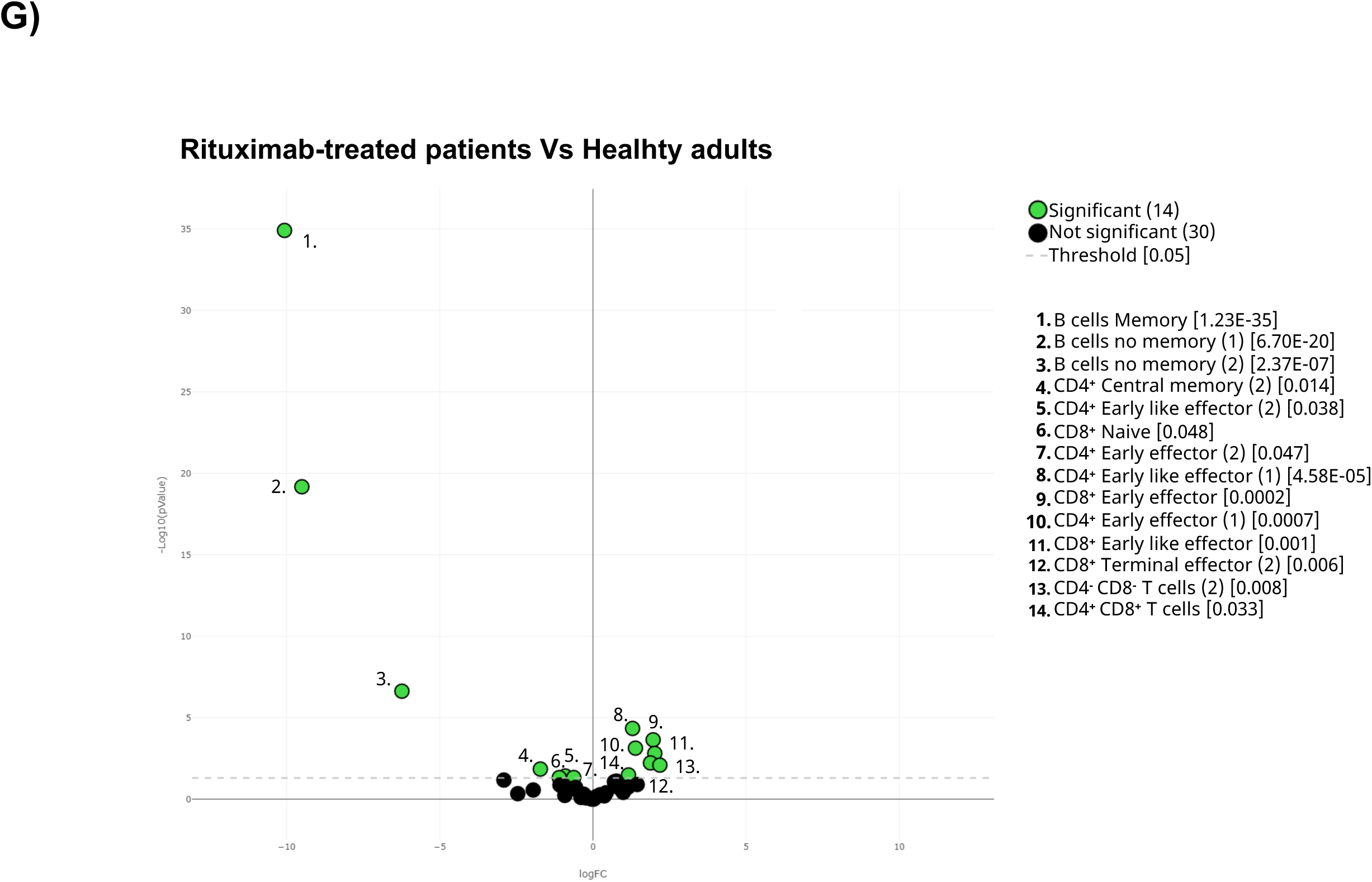
Differences of the cohorts before vaccination. **A)** The general UMAP plot was colored in blue (healthy adults), purple (older adults), green (untreated oncohematologic patients), yellow (lenalidomide-treated oncohematologic patients), navy blue (ibrutinib-treated oncohematologic patients) and light red (rituximab-treated oncohematologic patients) to display the cohorts distribution before vaccination referred to all the samples (shown in black). **B)** The heatmap displays the intensity levels of each 48 identified clusters within cohorts. A color code was based on the expression intensity, where green represent higher expression and the transition to red represent lower expression. The dendrogram was generated by unsupervised hierarchical clustering. Volcano plots comparing the clusters identified in Table 2 and Figure 1 between healthy adults and **C)** older adults; **D)** untreated patients; **E)** lenalidomide-treated patients; **F)** ibrutinib-treated patients; and **G)** rituximab-treated patients. For the volcano plots, those clusters which were differentially expressed (p-value <0.05) in the comparisons are highlighted in green. Due to the low number of events, some clusters could not be analyzed in panel C) (T_regs_ CD45RA^-^CD39^+^, Non-classical Monocytes, Plasmablasts, CD4^-^CD8^-^ T-cells (2), Tγδ CD45RA^+^CCR7^-^ (4) and Tγδ CD45RA^+^CCR7^-^(5)); and panel D (non-classical monocytes, plasmablasts, Tγδ CD45RA^+^/CCR7^-^ (4) and Tγδ CD45RA^+^/CCR7^-^ (5)) and panel G) (T_regs_ CD45RA^-^ /CD39^+^, non-classical monocytes, plasmablasts and Tγδ CD45RA^+^ /CCR7^-^ (4)).

In order to quantify these differences, a paired comparison of each cohort referred to the healthy controls one was performed. Hence, a third of the total identified clusters were differentially expressed among older adults and healthy controls since older adults displayed an expansion of CD4^+^ and CD8^+^ effector T-cells and deficit of monocytes and immature T-cells (Figure 3C). In order to further confirm these results, a classical gating strategy was performed confirming that older adults have an expansion of mature NK cells together with several subsets of effector CD4^+^ and CD8^+^ T-cells, and a deficit of immature T-cells (mainly CD4^-^CD8^-^ T-cells and naïve CD8^+^ T-cells), plasmablasts and CD45RA^+^CCR7^-^ Tγδ (Supplementary Figures 1A and 1B). Moreover, and given that the clusters identified in Figure 1 and Table 2 do not always resemble a whole population as they can be further divided into subsets, validation of the specific clusters identified in Figure 3C was performed following the gating strategy displayed in Figures 2C and 2D. Hence, although for instance early effector CD4^+^ T-cells did not differ between healthy and older adults, subset 1 of this population (cluster 2 in Table2) was expanded in older adults, while its second subset (cluster 3 in Table 2) was reduced in older adults (Supplementary Figure 1C). In a similar manner, the expansion for instance of mature NK cells shown in older adults (Supplementary Figure 1A) was specifically due to tan expansion of subset 1 on mature NK (cluster 44 in Table2) while the second subset of this population (cluster 45 in Table 2) remained unchanged. Therefore, this approach confirms the relevance of further validating not just differences between different populations, but also within them.

Referred to oncohematologic patients, analyses were performed based on their treatment status. Hence, untreated oncohematologic patients had 14 clusters differentially expressed compared to healthy adults (Figure 3D). Further analysis of this cohort revealed that these patients had a deficit of non-classical monocytes, CD45RA^+^CCR7^-^ Tγδ cells, CD45Ra^-^CD39+ T_regs_ cells and early-like effector CD4^+^ T- cells, together with an expansion of terminal effector CD4^+^ T-cells and CD4^-^CD8^-^ T-cells (Supplementary Figure 1D). Moreover, when the analysis was performed based on the specific clusters (Supplementary Figure 1E) it was revealed that it was the subset 2 of the early like effector CD4^+^ T-cells (cluster 3 in Table 2) the one that it was reduced, while cluster 2 of the terminal effector CD4^+^ T-cells (cluster 7 in Table 2) was the one expanded in this population. Further analysis also revealed a deficit of the subset 1 of the no memory B-cells (cluster 35 in Table 2) in this population together with subset 2 of the CD4^-^ ILC (cluster 47 in Table 2). Finally, the analysis of CD45RA^+^CCR7^-^ Tγδ cells revealed that subset 2 of this population (cluster 24 in Table 2) were decreased while subset 5 (cluster 27 in table 2) was expanded.

As for the lenalidomide-treated they displayed differences in half of the analyzed clusters referred to the controls (Figure 3E), with the classical gating strategy displaying a loss in several CD4^+^ and CD8^+^ T-cells populations, together with non-classical memory, memory B-cells and CD45RA^-^CCR7^-^ Tγδ cells. On the contrary, these patients and an expansion of CD45RA^-^CCR7^+^ Tγδ cells and CD4^-^ ILC (Supplementary Figures 1F and 1G). Further analyses revealed the specific subsets responsible for these differences (Supplementary Figure 1H).

Referred to ibrutinib-treated patients, 44% of the clusters were differentially expressed referred to healthy adults (Figure 3F). Hence, following hierarchical gating identification of the populations it was clear that this cohort has a deficit of memory B-cells, non- classical monocytes, naïve CD8’T-cells, CD45RA-CD39+ T_regs_, CD45RA^-^CCR7^-^ Tγδ cells and total ILC as well as CD4- ILC (Supplementary Figure 1I). On the other hand, they displayed an expansion of terminal effector CD4^+^ and CD8^+^ T-cells, early like effector CD8^+^ T-cells, CD4^-^CD8^+^ T-cells and CD45RA^-^CCR7^+^ Tγδ cells. In a similar manner, the specific subsets responsible for these differences are shown in Supplementary Figure 1J.

Last but not least, differences between rituximab-treated patients and healthy adults at baseline were performed. Results from volcano plot analysis displayed differences 29% of the total identified clusters including, as expected, a total B-cell depletion (Figure 3G and Supplementary Figure 1K). In addition, these patients also had a deficit of CD8^+^ naïve T-cells and an expansion of CD8^+^ early-like and terminal effector T-cells, together with CD45RA-CD39^+^ T_regs_. Further analysis revealed that although CD4+ T-cell populations were not altered in these patients, several of their specific subsets were nevertheless altered in this cohort (Supplementary Figure 1M).

### Acquired humoral and cellular memory

Having described the cohorts at baseline, we next studied the acquired humoral and cellular immunity 3 months after full immunization. SARS-CoV-2 neutralizing spike antibodies (IgG and IgA anti S) and nucleocapsid antibodies (IgG anti N) were determined in plasma before and over 3 months after vaccination (Figure 4A). A small fraction of healthy adults (14%), older adults (5%), and oncohematologic treated with lenalidomide (11.1%) and ibrutinib (7%) had IgG anti-N antibodies before vaccination suggesting an unnoticed previous asymptomatic infection. Of note, adult and older patients triggered IgG anti-S antibodies confirming vaccine success, although the percentage was lower in the oncohematologic patients, and virtually absent in Rituximab treated patients (Figure 4A) Also, given that vaccination was intramuscular, IgA antibodies production was only induced in healthy adults while it was not triggered in the older adults or the oncohematologic cohorts. Finally, we also assessed induced cellular memory revealing a strong response in all cohorts (including Rituximab-treated patients) although that was smaller in Lenalidomide- and absent Ibrutinib-treated patients (Figure 4B).

**Figure 4:**
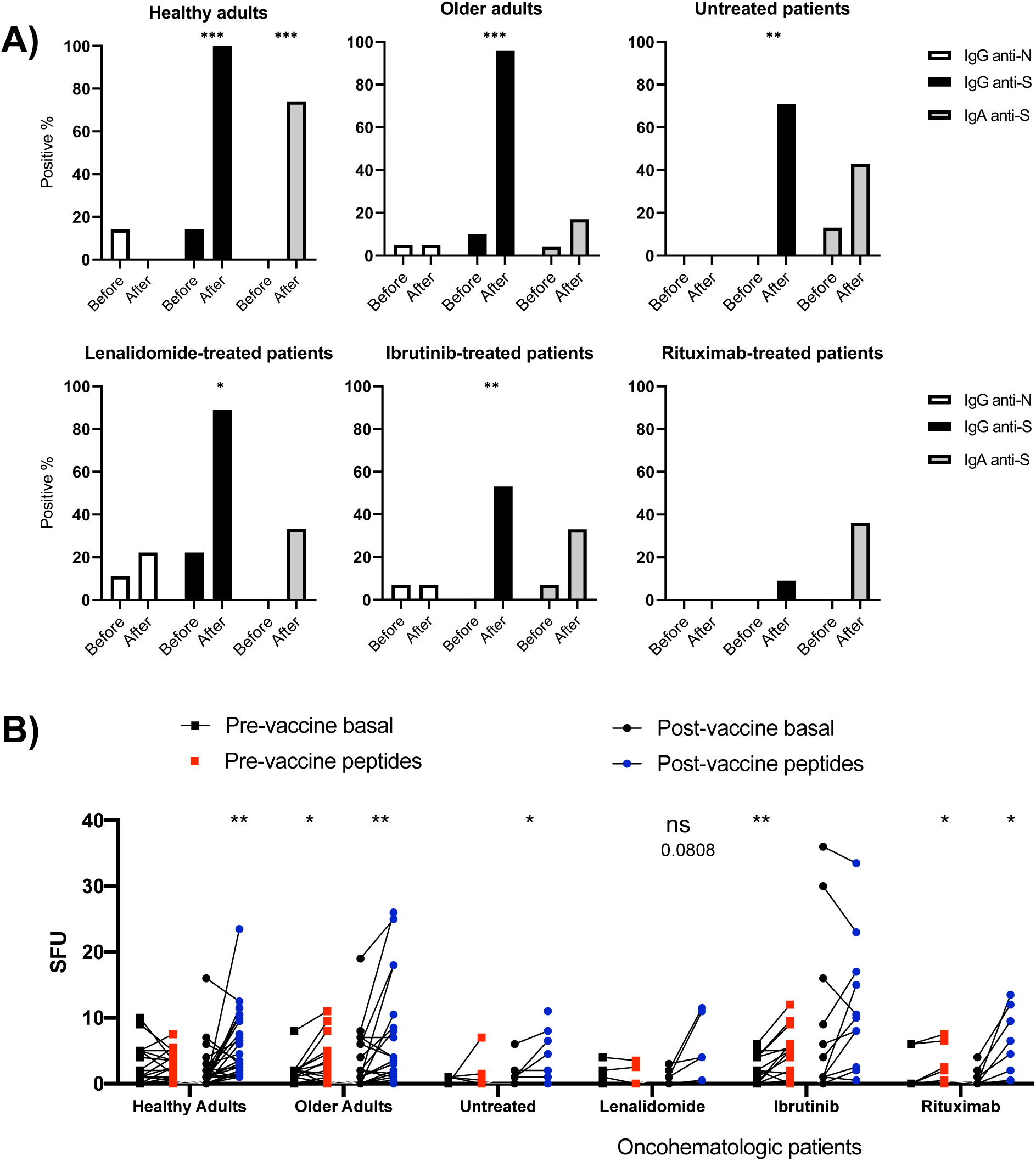
Vaccine-induced humoral and cellular memory. **A)** Humoral memory against SARS-CoV-2 before and after vaccination. Analysis of Immunoglobulin G (black) and A (shaded) anti Spike protein and IgG anti nucleocapsid (white) were performed. Results were based on the number of patients who tested positive in serology. **B)** Cellular memory against SARS-CoV-2 before and after vaccination analyzed by IFN-γ ELISpot assay. Each cohort was analyzed independently by comparing the spot forming units (SFU) under both basal (black dots) and SARS- CoV-2 peptide-stimulated (blue and red dots) conditions. A fisher test was applied in panel A) while a paired One-way ANOVA was applied in panel B). In all cases, a p-value <0.05 was considered significant (*<0.05; **<0.01; ***<0.001), while a p-value <0.10 was considered as not significant (ns) but with a relevant trend displaying the p-value underneath.

### Immunity induced changes following vaccination

Having observed the differences before vaccination in all cohorts, an analysis of the differences before and after full vaccination was performed (Supplementary Figure 2A). Hence, healthy adults expanded the proportion of the subset 3 of the central memory CD4^+^ T-cells (cluster 10 in Table 2) and CD45RA-CD39^+^ T_regs_ (Supplementary Figure 2B) although that could not be further validated by classical gating approaches.

As for the older adults, vaccination induced changes in 15% of the clusters (Supplementary Figure 2C) as it expanded the proportion of circulating CD4^-^CD8^-^ T-cells and early like effector CD4^+^ T-cells and decreased the proportion of classical and non- classical monocytes (Supplementary Figure 2D). Further analysis revealed that vaccination decreased the levels of both subsets of classical monocytes and CD4^-^CD8^-^ T-cells, while expanded the proportion of subset 2 of early effector CD4^+^ T-cells (cluster 3 in Table 2) and subset 2 of early like effector CD4^+^ T-cells (cluster 5 in Table 2).

Referred to untreated oncohematologic patients, vaccination decreased the circulating levels of circulating memory B-cells and plasmablasts (Supplementary Figure 2F) although that could not be further confirmed by hierarchical gating approaches. On the contrary, vaccination induced changes in 4 clusters from the lenalidomide-treated patients revealing a decreased post-vaccination of CD4^-^CD8^-^ T-cells and CD45RA^-^ CD39^+^ T_regs_ (Supplementary Figure 2H). Also, the decrease within the CD4^-^CD8^-^ T-cells was due to a reduction of the second subset of this population (cluster 22 in Table 2) while these patients also expanded subset 5 of the CD45RA^+^CCR7^-^ Tγδ cells (cluster 27 in Table 2). As for the ibrutinib-treated patients, vaccination decreased the levels of circulating monocytes (Supplementary Figure 2J) although that could not be confirmed by classical gating approaches. Finally, vaccination induced changes in 4 clusters of the rituximab-treated patients (Supplementary Figure 2K) confirming that these patients had a trend to decrease the level of non-classical monocytes (Supplementary Figure 2L) and subset 2 of the no memory B-cells (cluster 36 in Table 2) following vaccination (Supplementary Figure 2M).

### Immune variations following vaccination predicts SARS-CoV-2 infection

Having assessed the vaccine-induced immunity after vaccination, a clinical follow-up was performed to all individuals during a period of 18 months. Hence, a total of 30.4% of the healthy adults displayed subsequent SARS-CoV-2 infection as defined by a positive PCR (Table 1). This percentage, however, was much lower in the older adults (15%) as they were protected in a nursing home environment. Finally, 41.12% of the oncohematologic patients had an infection. Of note, given the low number of infected patients, these patients were considered as a single entity in the subsequent analyses irrespectively of their treatment.

The acquired humoral and cellular memory following immunization did not predict subsequent infection for any of the analyzed cohorts (Table 3). Nevertheless, when the cellular immunome post-vaccination of all individuals was studied in the context of subsequent infection, differences in the UMAP analysis were found (Figure 5A) revealing differences in 5 clusters (Figure 5B). Hence, infected individuals had lower levels of CD4^+^CD8^+^ T-cells and a trend towards higher levels CD45RA^+^CCR7^-^ Tγδ cells and terminal effector CD8^+^ T-cells and (Supplementary figure 3A), due to an expansion in the second case of subset 1 (cluster 15 in Table 2) of this population (Supplementary Figure 3B). In order to get a deeper insight into these observations, further analysis was performed on healthy adults and oncohematologic patients, while given the low number of post-vaccinated analyzed infected older adults with enough number cells to perform the analysis, this cohort was excluded.

**Table 3:**
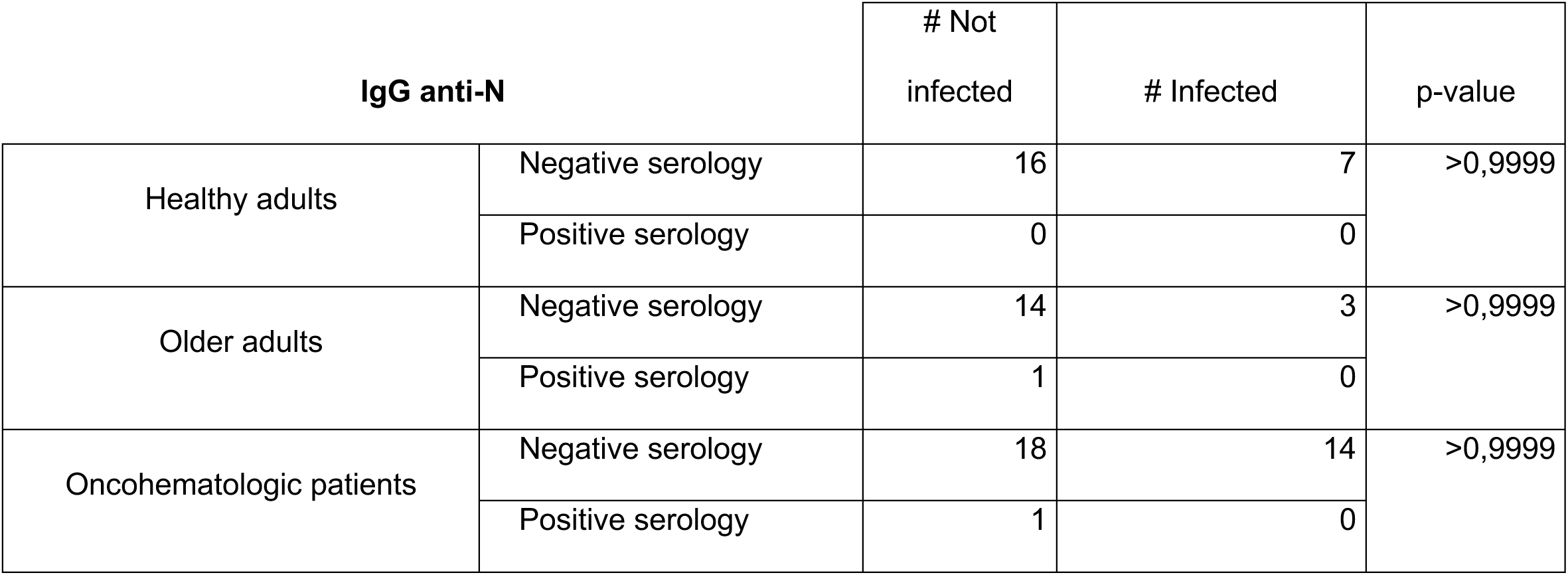
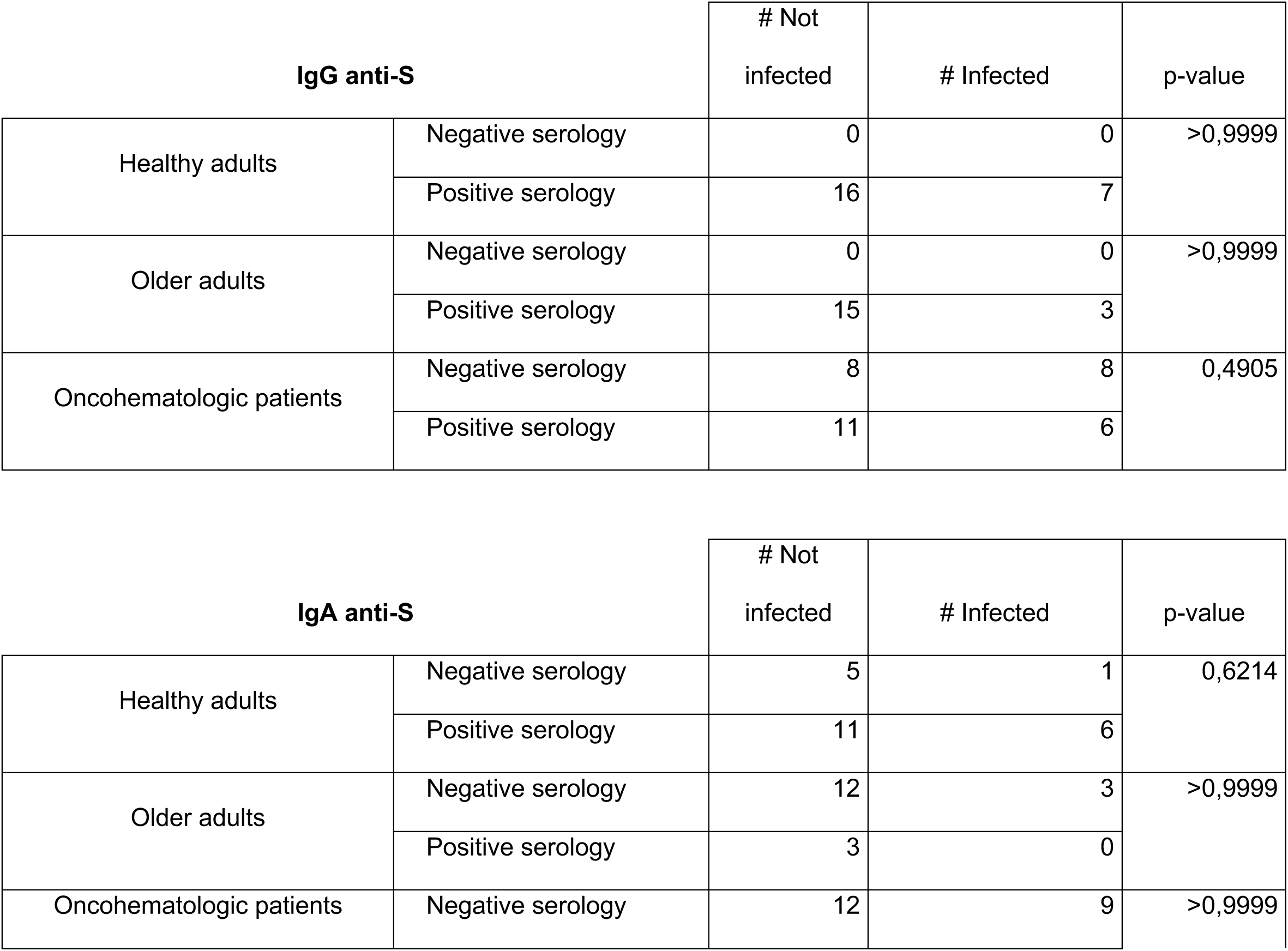
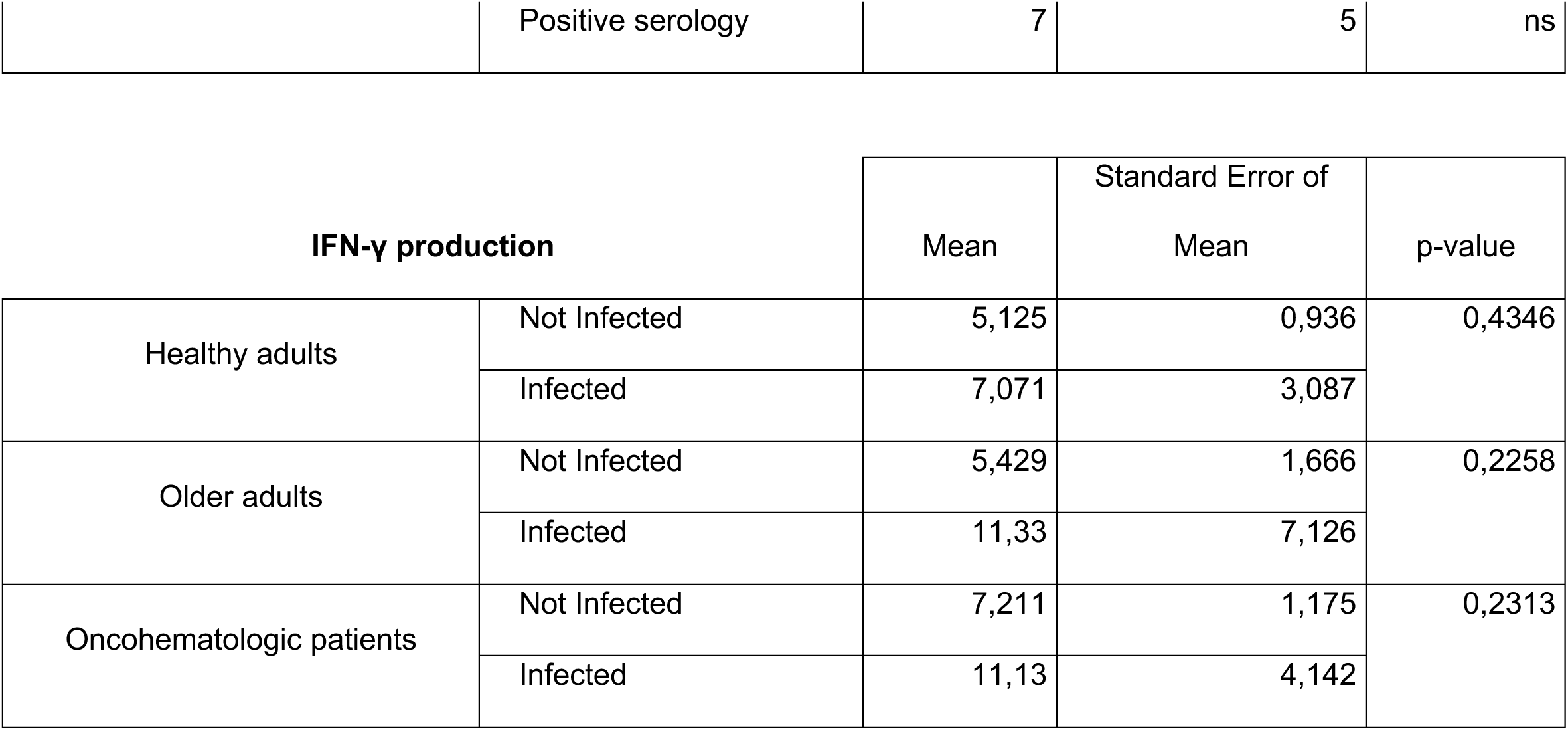
Infection based on humoral and cellular memory. For each cohort, the absolute number of infected and not infected individuals referred to their serology status is shown for immunoglobulin (Ig) G and A towards SARS-CoV-2 Spike protein (S), and IgG against SARS-CoV-2 nucleocapsid protein (N). The mean and standard error for IFNγ production following ELISpot assay towards the S protein is also shown based on subsequent infection. Fisher test was applied for the humoral memory while a t-test was used the cellular one. P-value <0.05 was considered statistically significant

**Figure 5:**
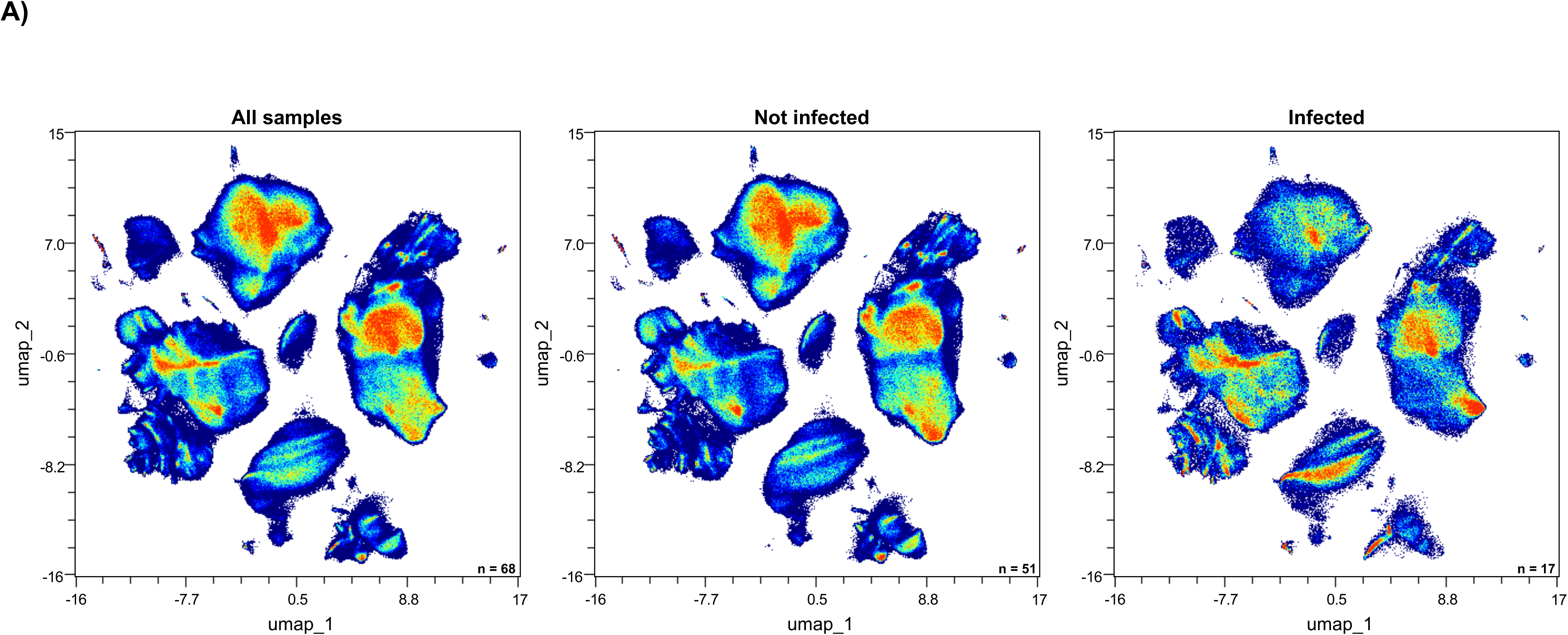
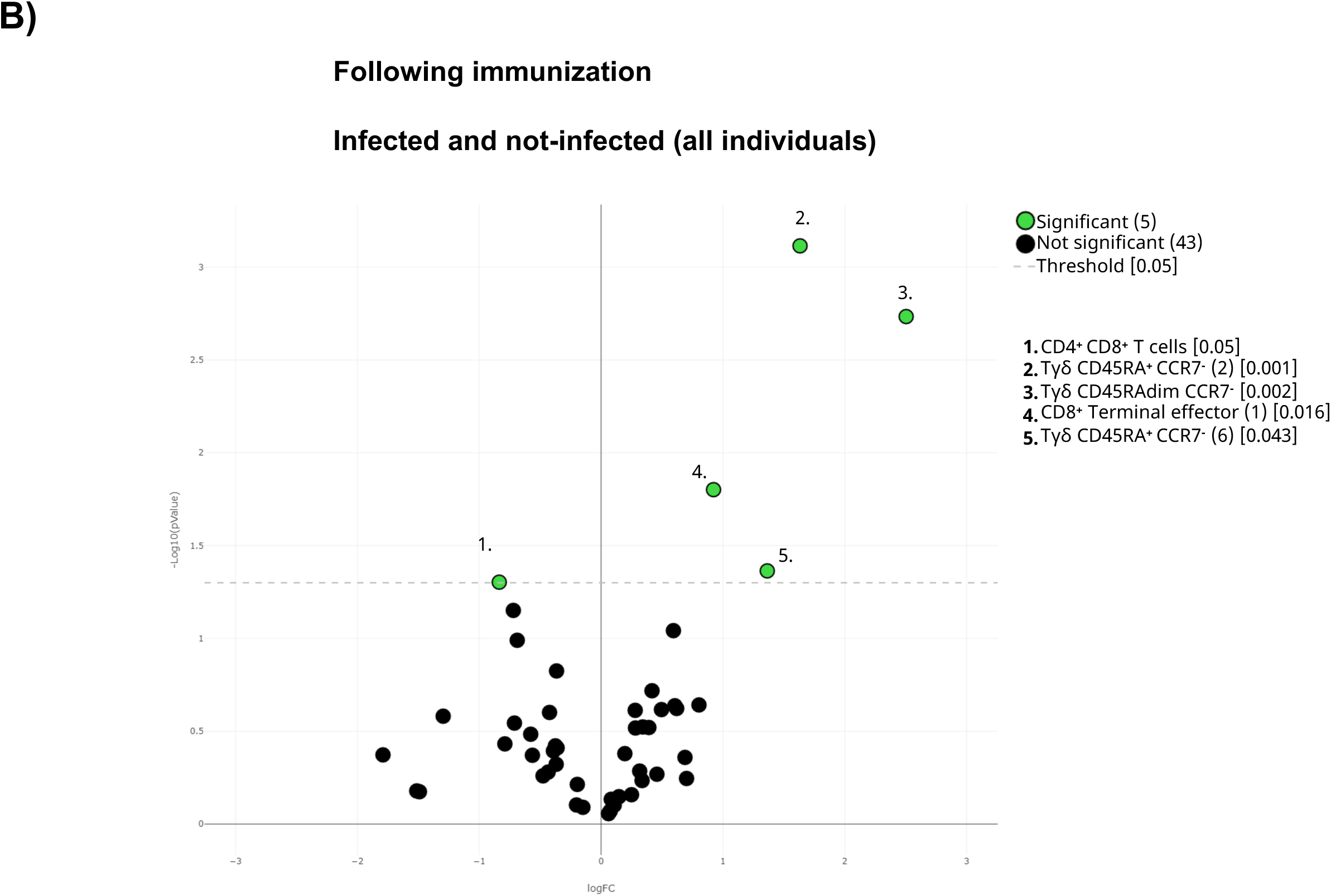
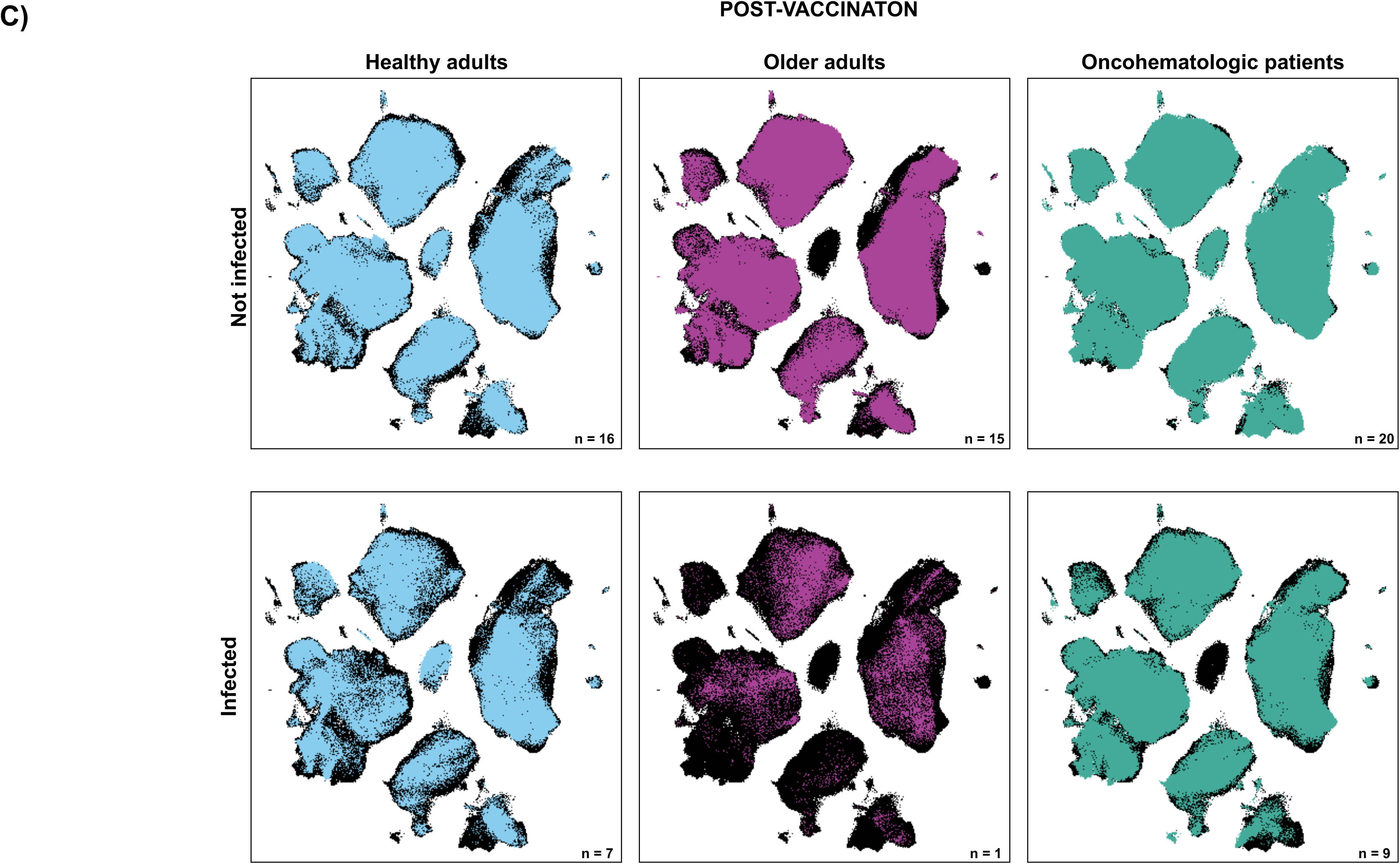
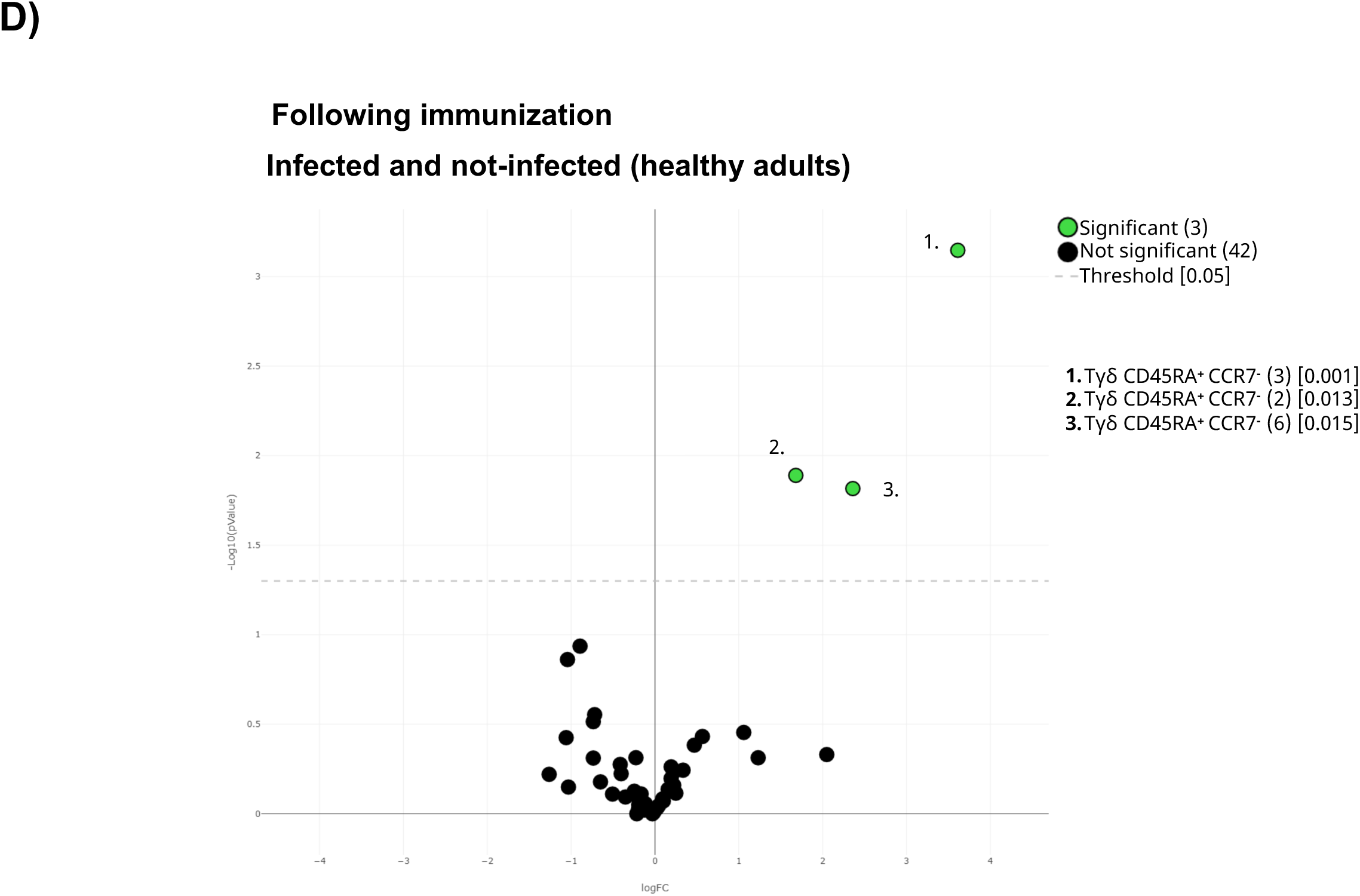
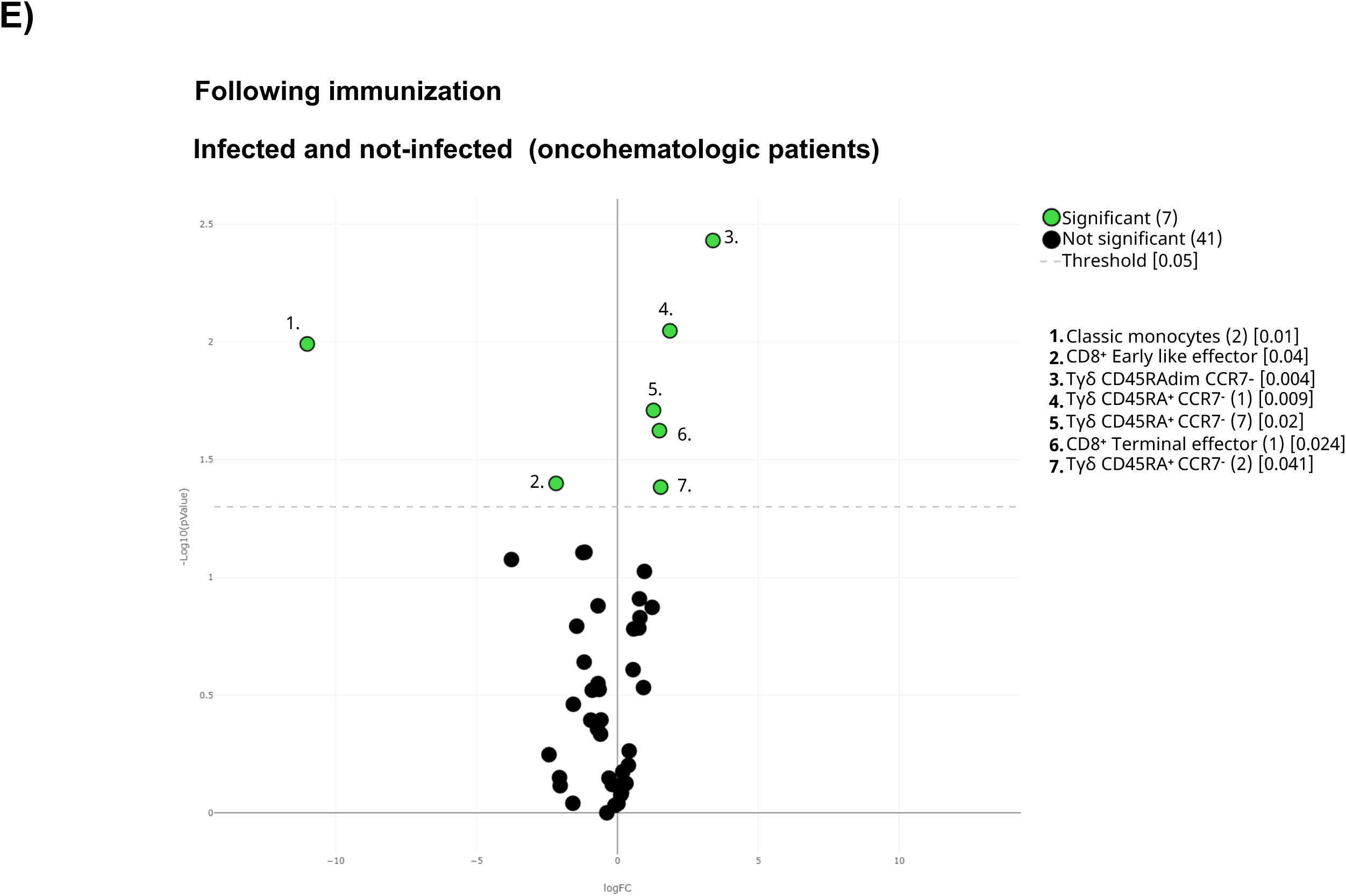
Cellular immunome post-vaccination predicts subsequent SARS-CoV-2 infection. **A)** All samples following full immunization were displayed in the UMAP density plots based on their subsequent infection defined by a positive PCR. **B)** Volcano plot comparing the clusters identified in Table2 and Figure 1 based on subsequent infection. **C)** UMAP plot of the infected and not-infected samples were colored in blue (healthy adults), purple (older adults), and green (oncohematologic patients) referred to all samples (shown in black). Volcano plots of the healthy adults and oncohematologic patients are shown in **D)** and **E)** respectively. For the volcano plots, those clusters which were differentially expressed (p-value <0.05) in the comparisons are highlighted in green. Due to the low number of events, some clusters could not be analyzed in panel D) (plasmablasts, CD4^-^CD8^-^ T-cells (2) and Tγδ CD45RA^+^/CCR7^-^ (4)).

The UMAP analysis revealed differences between the infected and not infected older adults and oncohematologic patients (Figure 5C). Hence, infected healthy adults had higher levels of 3 different subsets of CD45RA^+^CCR7^-^ Tγδ cells (Figure 5D) although that could not be further confirmed by hierarchical gating approaches. In a similar manner, infected oncohematologic patients had differences in 15% of the total clusters (Figure 5E) confirming, as in the healthy adults, and expansion of CD45RA^+^CCR7^-^ Tγδ cells (Supplementary figure 3C) due to an expansion of subset 1, 2 and 7 (clusters 23, 24 and 29 in Table 2) of this population (Supplementary Figure 3).

### Pre-vaccine immunome predicts subsequent SARS-CoV-2 infection

Given that we could predict subsequent SARS-CoV-2 infection based on the circulating immunome following vaccination, we next addressed whether these observations could be also observed before vaccination. Hence, there were differences in the immunome composition before vaccination between subsequently infected and not infected individuals, even before vaccination (Figure 6A) as 21% of the total clusters were differentially expressed between infected and not-infected individuals (Figure 6B). Hence, before vaccination subsequently infected individuals had higher levels of circulating terminal effector CD8^+^ T-cells and plasmablasts, coupled with a trend towards higher levels of CD45RA^+^CCR7^-^ Tγδ, and lower levels of early NK and total ILC (Supplementary Figure 4A). Further analysis confirmed that subset 3 of the CD45RA^+^CCR7^-^ Tγδ (cluster 25 in Table 2) was expanded in the infected individuals even before vaccination (Supplementary Figure 4B).

**Figure 6:**
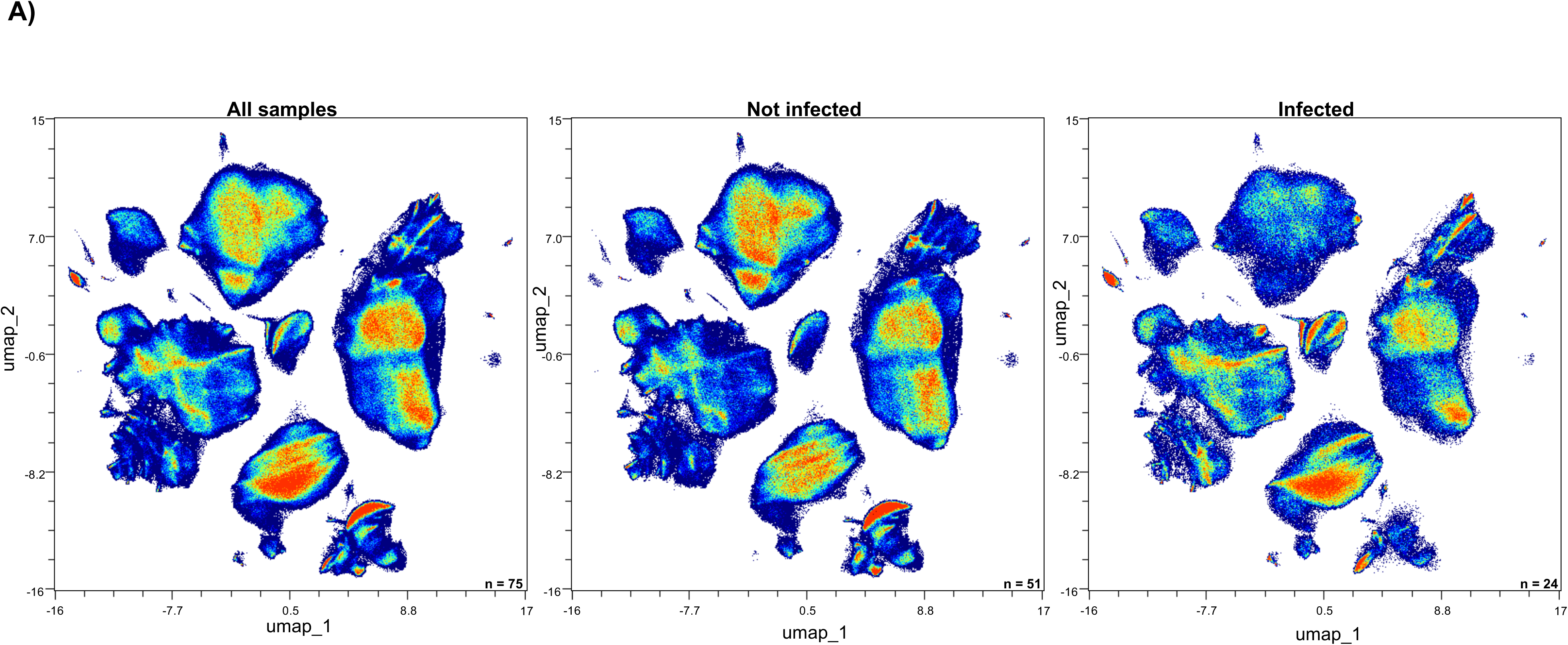
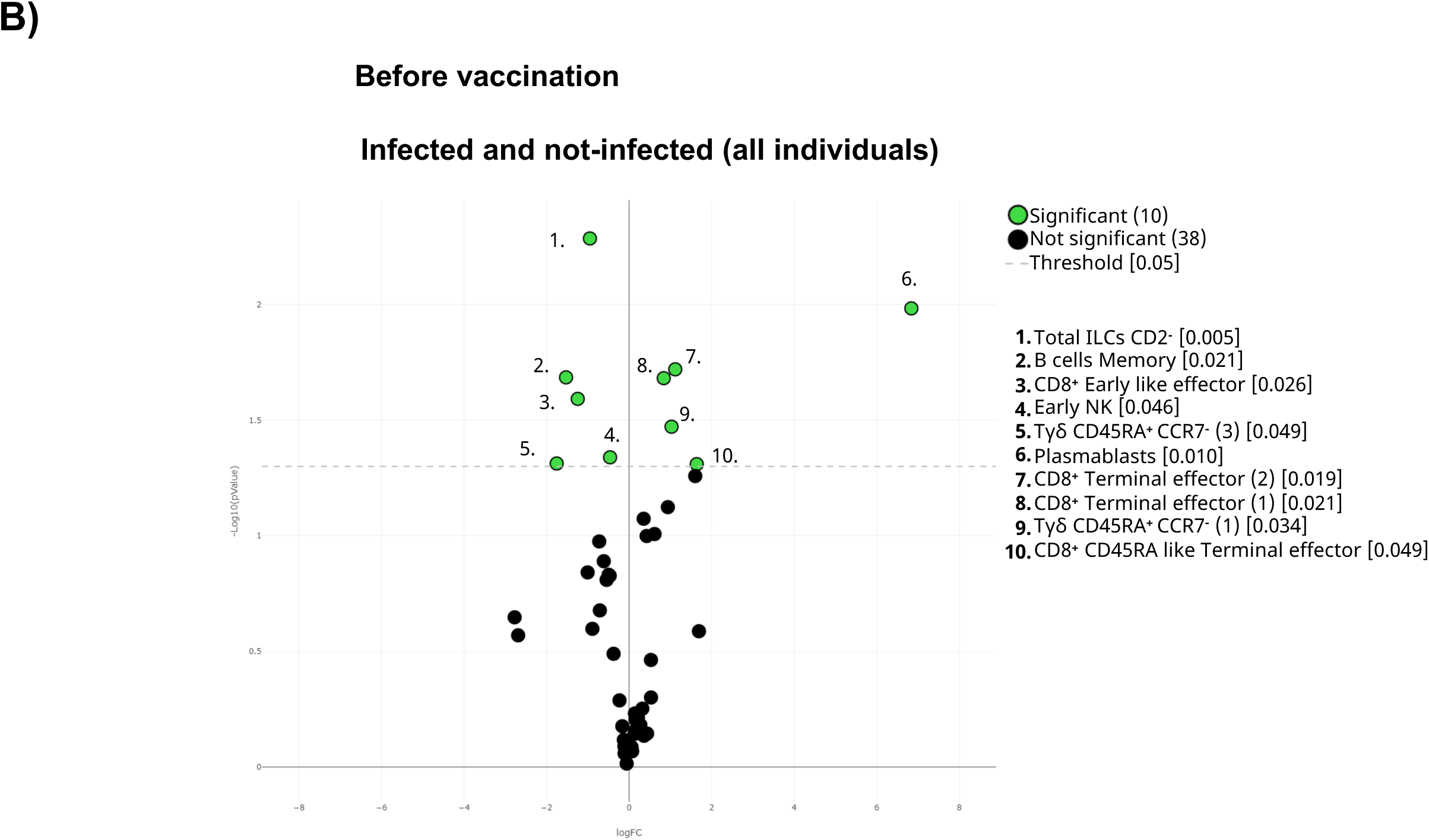
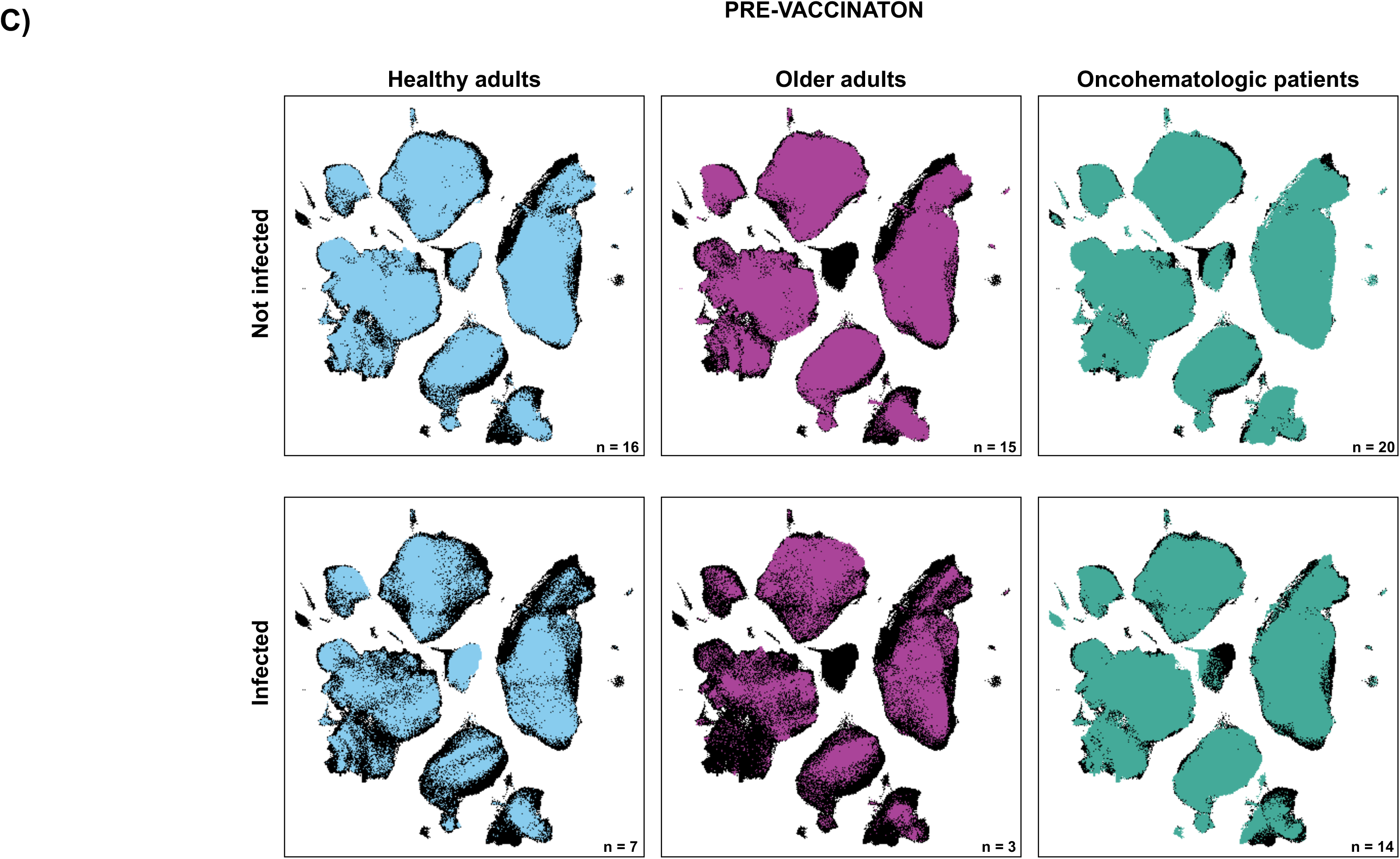
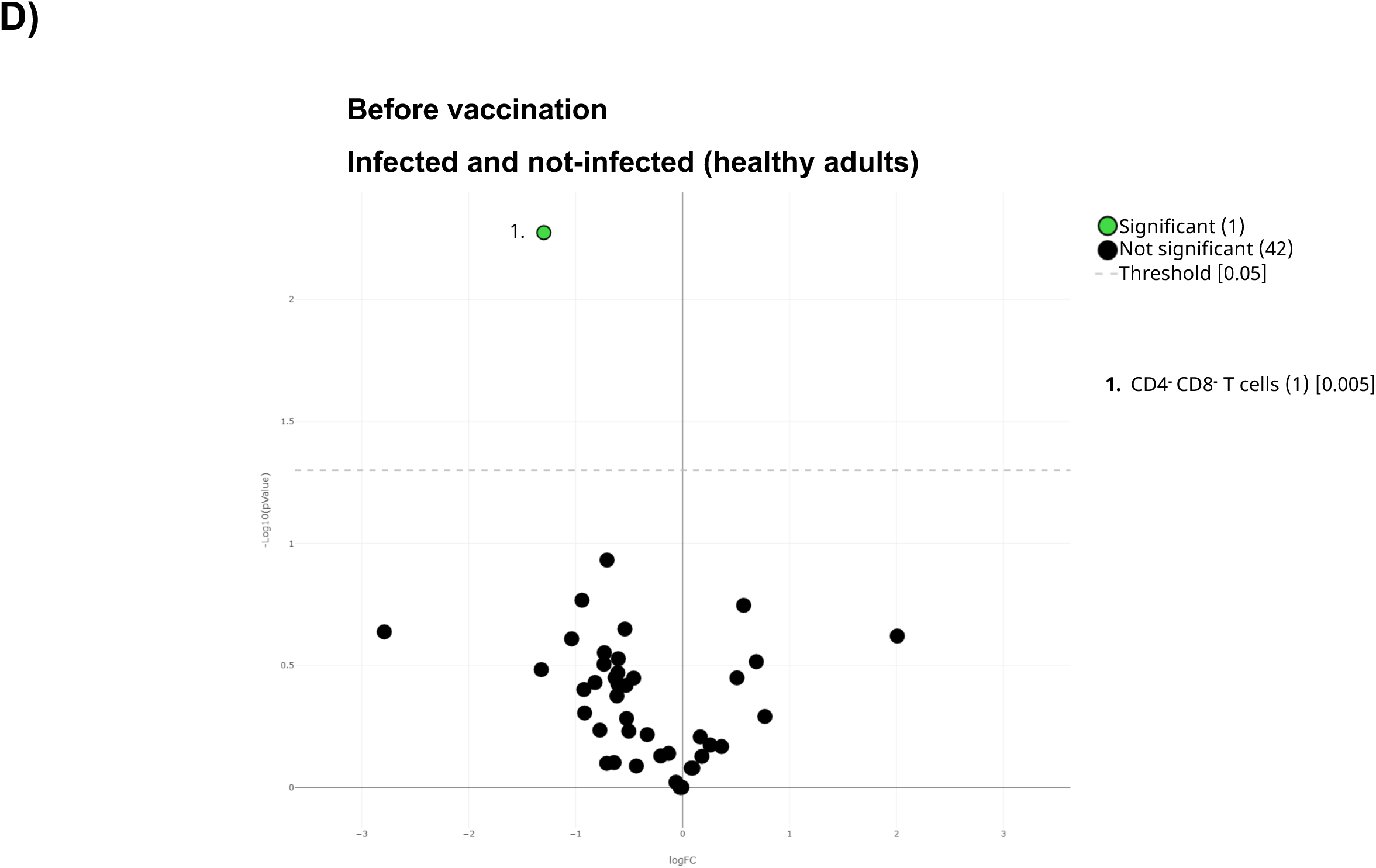
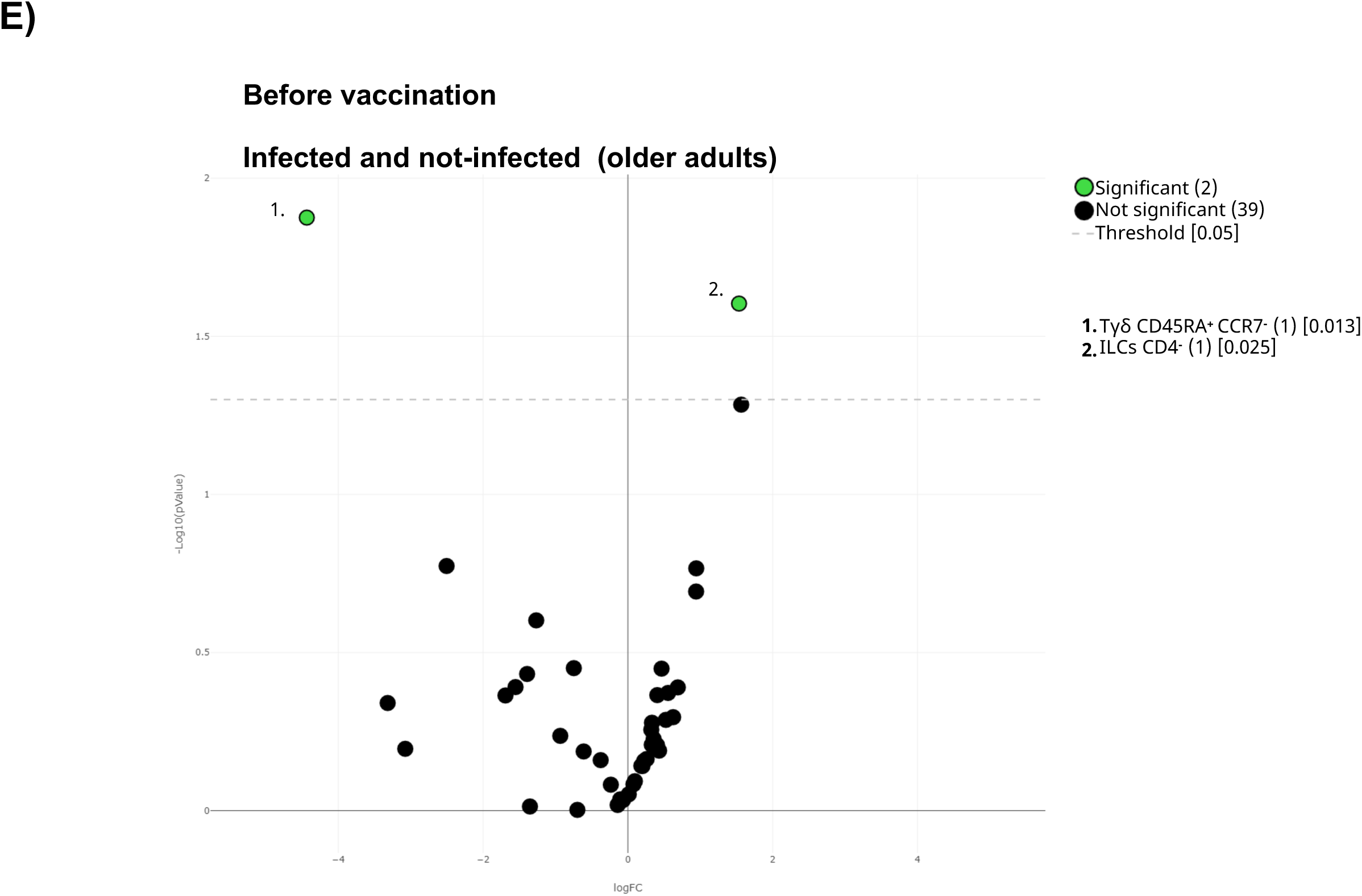
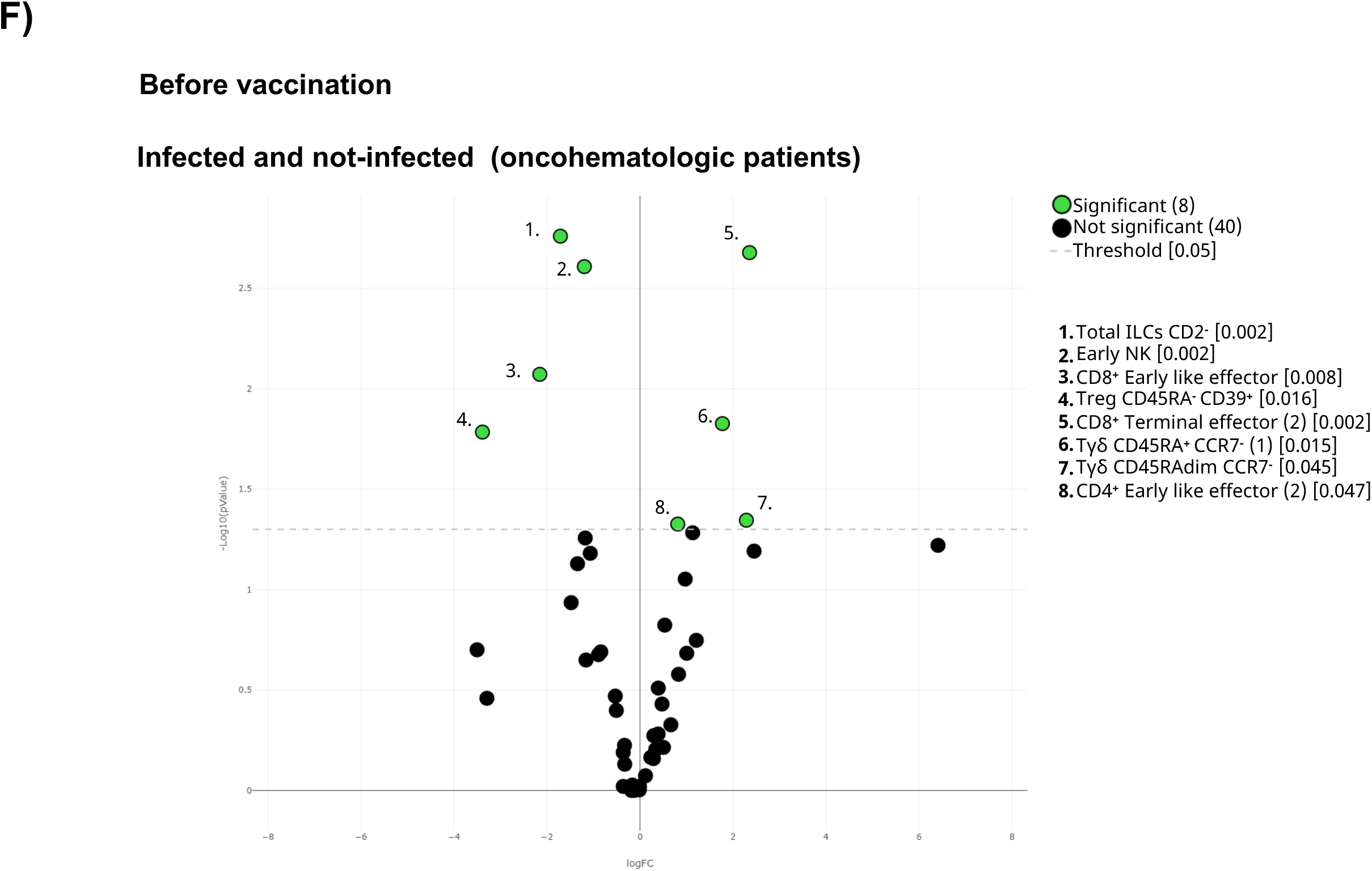
Pre-vaccination cellular immunome predicts SARS-CoV-2 infection. **A)** All samples before the first dose were displayed in the UMAP density plots based on their subsequent infection. **B)** General Volcano plot comparing the clusters identified in Table2 and Figure 1 before vaccination based on subsequent infection. **C)** UMAP plot of the cohorts before being vaccinated were colored by blue (healthy adults), purple (older adults) and green (oncohematologic patients), referred to all samples (shown in black). Specific volcano plot of **D)** healthy controls, **E)** older adults and **F)** oncohematologic patients based on subsequent infection were also shown. For the volcano plots, those clusters which were differentially expressed (p-value <0.05) n the comparisons are highlighted in green. Due to the low number of events, some clusters could not be analyzed in panel D) (T_regs_ CD45RA^-^/CD39^+^, non-classical monocytes, plasmablasts, CD4^-^/CD8^-^ T cells (2) and Tγδ CD45RA^+^/CCR7^-^ (4)) and E) (classic monocytes (2), T_regs_ CD45RA^-^/CD39^+^, non-classical monocytes, plasmablasts, CD4^-^/CD8^-^T cells (2), Tγδ CD45RA^+^/CCR7^-^ (4) and Tγδ CD45RA^+^/CCR7^-^ (5)).

**Figure 7:**
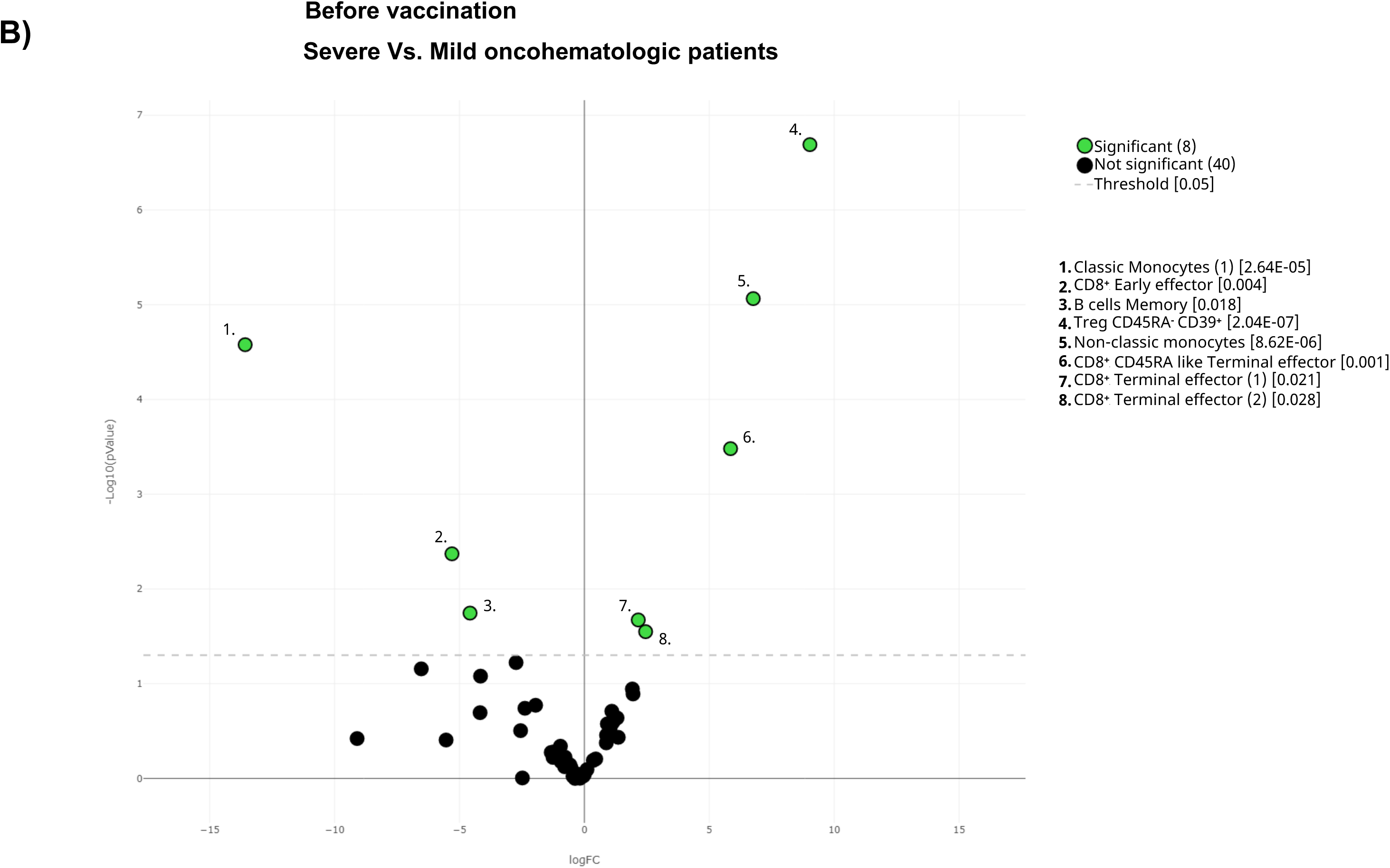
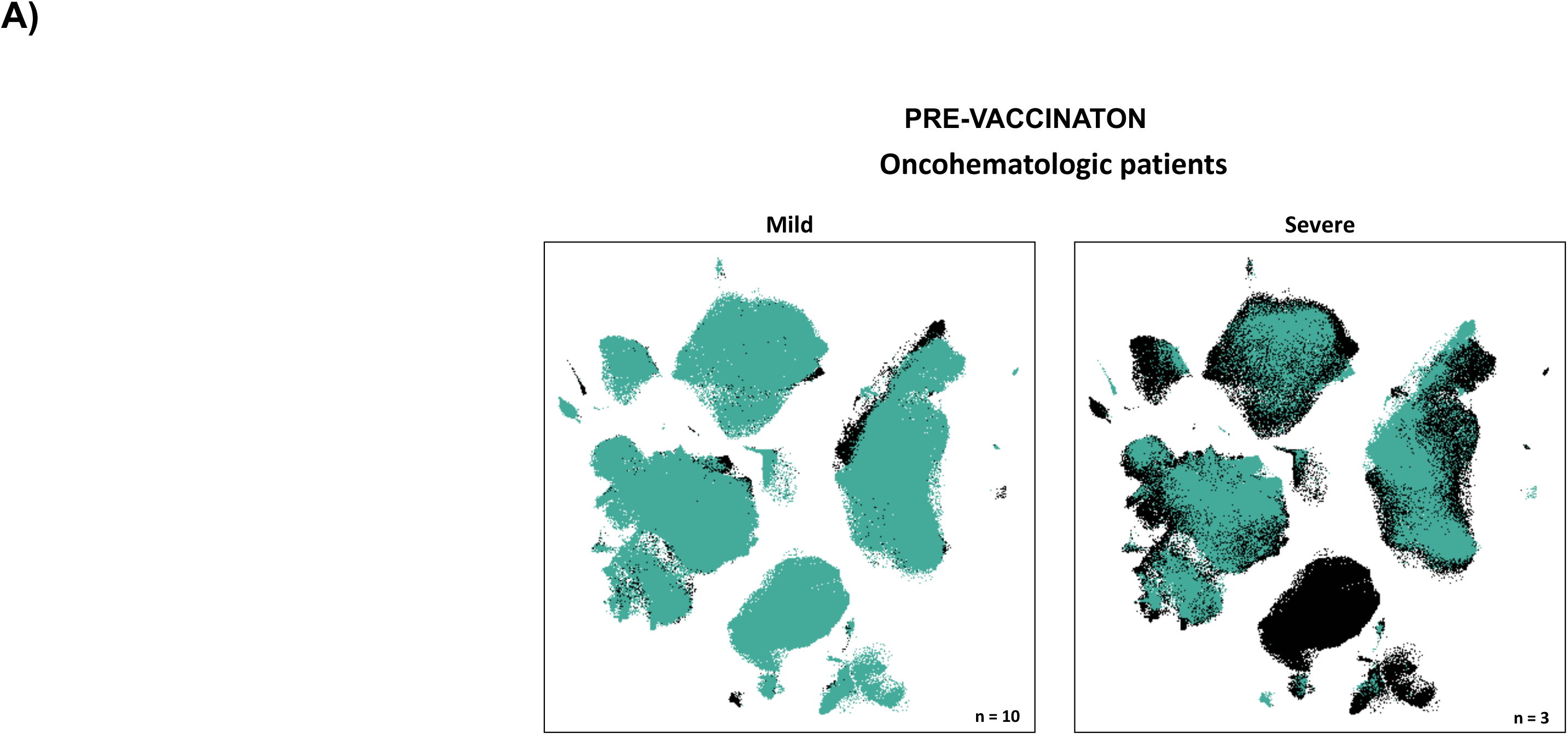
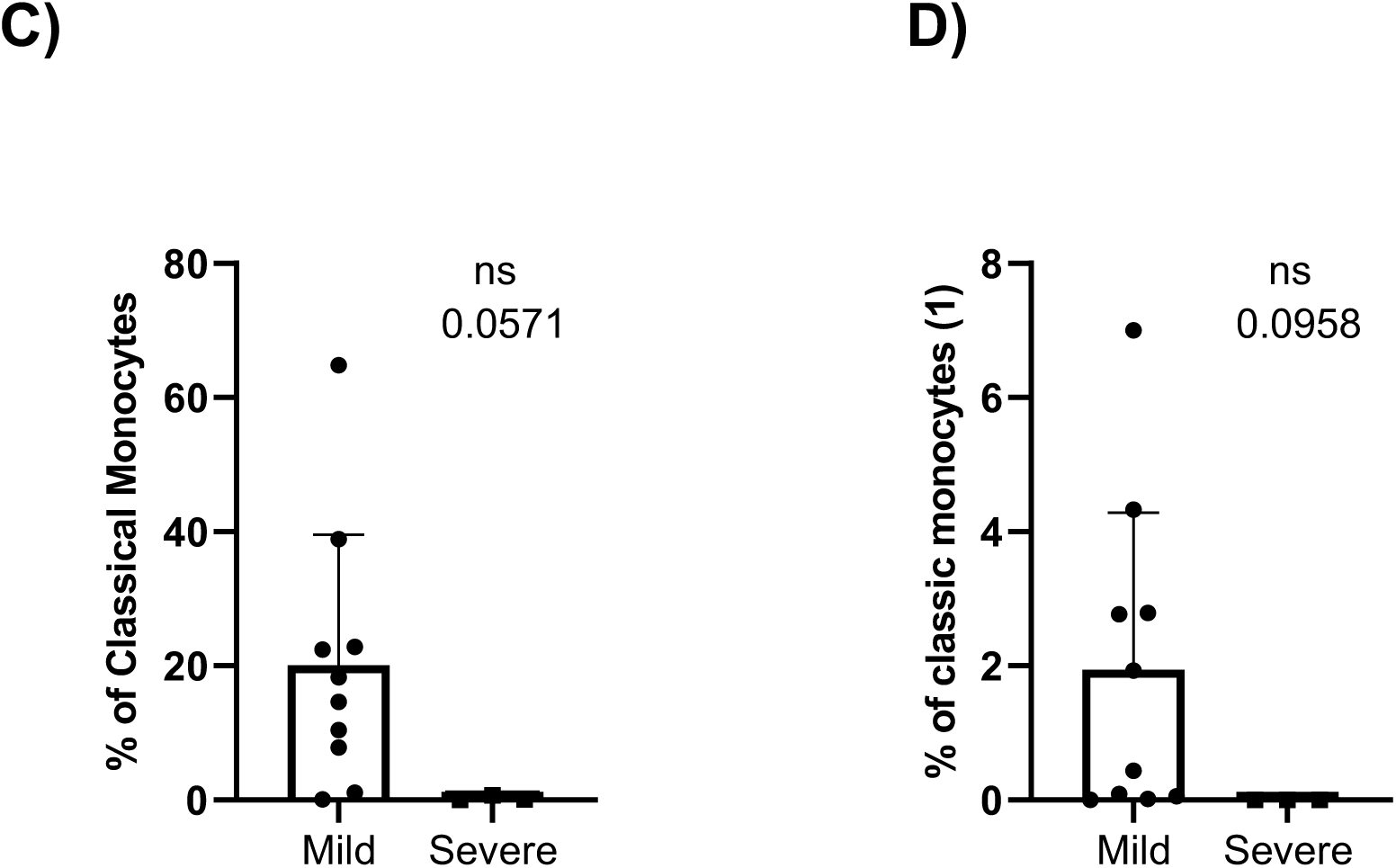
Cellular immunome before vaccination predicts COVID-19-induced hospitalization. **A)** UMAP plot of the pre-vaccination samples from infected oncohematologic patient based disease outcome as mild (left) and severe (right) defined as the absence or need of subsequent COVID-induced hospitalization respectively. **B)** Volcano plot analysis comparing the clusters identified in Table2 and Figure 1, within infected oncohematologic patients based on disease outcome. **C)** Classical validation, following the gating strategy shown in Figure 2, of total monocytes, and **D)** the classic monocyte (1) cluster. In panel B), green dots represent those clusters that showed significant differences (p- value<0.05). A one-tailed t-test was applied on panels C) and D), showing the p-value in the figure (ns, -not statistically significant-).

Having seen these differences, the analysis was subsequently performed within the 3 different cohorts (Figure 6C). Although healthy adults only had differences in 1 cluster based on the subsequent infection (Figure 6D), hierarchical gating revealed a trend towards higher levels of circulating plasmablasts and non-classical monocytes (Supplementary Figure 4C), with a deficit of subset 1 of CD4^-^CD8^-^ T-cells (cluster 21 in Table 2) and a trend to lower levels of subset 4 of the CD45RA^+^CCR7^-^ Tγδ (cluster 26 in Table 2) in the subsequently infected individuals (Supplementary Figure 4D). As for older adults, only 2 cluster were differentially expressed before vaccination between subsequent infected and not-infected individuals (Figure 6E), with this cohort showing a trend towards lower levels of subset 5 of CD45RA^+^CCR7^-^ Tγδ cells (cluster 27 in Table 2) in the infected individuals. Finally, 17% of the clusters were differentially expressed in the oncohematologic patients (Figure 6F) as these patients had higher levels of CD45RA^+^CCR7^-^ Tγδ and a trend towards lower levels of early NK cells (Supplementary Figure 4F), within a specific expansion before vaccination of subset 1 of the CD45RA^+^CCR7^-^ Tγδ (cluster 23 in Table 2) in the subsequently infected patients (Supplementary Figure 4).

### Pre-vaccine immunome analysis in oncohematologic patients predicts COVID- induced hospitalization

A total of three out of the thirteen infected oncohematologic patients had vaccine failure as defined by SARS-CoV-2 induced hospitalization (Table 1). Hence, given that the immunome composition before vaccination can predict subsequent infection, we finally addressed whether the same could be true, within the oncohematologic cohort, to predict vaccine failure. UMAP immunome analysis of this cohort divided by mild (no hospitalization) and severe disease (required hospitalization) displayed evident differences between them (Figure 6A), revealing a total of 8 clusters differentially expressed in these patients even before vaccination (Figure 6B). Hence, patients with a severe outcome displayed an expansion of T_regs_, non-classic monocytes and CD8^+^effector T-cells although these findings could not be confirmed by classical gating approaches likely due to the low number of patients with severe outcome (data not shown). Nevertheless, patients with a severe outcome displayed a trends towards a deficit of circulating monocytes (figure 6C) due to a deficit of its cluster 1 (cluster 40 in Table 2) confirming therefore the relevance of this population to control subsequent infection.

## DISCUSSION

For the past three years of pandemic, the immune system of the vast majority of humans came into contact with SARS-CoV-2 through vaccination, infection, or both. Vaccination has been crucial to contain the impact of the COVID-19 pandemic^14^. New evidence suggests that ‘hybrid’ immunity as result of both vaccination and natural infection, can provide partial protection against reinfection for at least eight months^15^. However, long- term immune protection has proved to be more complex than initially suggested. To date, only two systematic reviews give meta-analytical evidence on the duration of COVID-19 vaccine effectiveness^16, 17^ with both of them displaying a general decrease of vaccine effectiveness over time against infections, hospitalizations, and mortality. This seems especially true for humoral response elicited by mRNA vaccines that can be escaped by variants of concern rather than for T-cell mediated responses^18^. Accordingly, adaptive immune response is known to play an important role in viral clearance, disease containment and resolution^19, 20^.

In order to get a deeper insight into the duration of the acquired immunity, we hereby have performed an in depth and unbiased characterization of the immune system before and after immunization. Hence, and although the acquired levels of humoral and cellular memory could not predict subsequent infection, the immunome analysis predicted not just subsequent PCR-confirmed infection following immunization, but also prior to that. Nevertheless, it is true that PCR was not systemically done on regular bases to all the individuals during the length of the study. Therefore, it is currently unknown whether those patients who did not become infected is because they certainly did not or whether, on the contrary, they were also exposed to the virus but they were more efficient at controlling the infection so they did not develop any symptom and, as a consequence, no PCR was performed. Nevertheless, an obvious consequence of vaccine failure is COVID-induced hospitalization which, in our case, was restricted to the ibrutinib-treated oncohematologic patients revealing a unique immune fingerprint in these patients even before vaccination.

We are nonetheless aware that this is a pilot study with a restricted sample size. Hence, it is important to highlight that following the computational cytometry analysis pipeline, although several clusters were found to be differentially expressed between the several comparisons performed, these results could not be always validated following hierarchical gating approaches. This is likely due to low number of available samples since, indeed, many of the performed comparison already display a trend. Therefore, we cannot discard the possibility that most of the clusters identified in the volcano plots could had been validated by classical gating approaches if larger number of individuals had been enrolled in this pilot study. Having said that, those cell populations and clusters subsets that were validated following classical hierarchical approaches provide further strength to our findings. In this regard, the use of unsupervised analyses reveals the presence of several subsets within a given population which, otherwise, could had remained unnoticed. A clear example of that is the CD45RA^+^CCR7^-^ Tγδ population, where up to 7 subsets can be found based on the differential expression of several surface markers (Table 2 and Figure 2D). In this regard, it is important to highlight that several subsets of this population could predict vaccine failure in the different cohorts, not just following vaccination but also before that. For instance, subset 1 of this population (cluster 23 in Table 2) was expanded in subsequently infected oncohematologic patients, both before and following immunization. In a similar manner, subsets 2 (cluster 24 in Table 2) and 7 (cluster 29 in Table 2) were also expanded in the oncohematologic patients before vaccination, while subset 4 (cluster 26 in Table 2) and 5 (cluster 27 in Table 2) were decreased in healthy and older adults respectively before vaccination. Hence, these findings confirm the relevance of the different Tγδ populations to control SARS-Cov-2 infection. Therefore, based on these results it is clear that this population and their subsets should be further addressed in depth not just to understand the specific mechanisms by which the control SARS-CoV-2 infection, but also to be used as novel biomarkers to monitor and predict infection in immune compromised individuals.

More generally, an important consideration when studying vaccine responses is prior immunologic encounters. For example, prior mild SARS-CoV-2 infection followed by complete clinical recovery poises individuals, particularly in males, to mount a more robust response to subsequent FLU vaccination^21^. This is owed to long-lasting antigen- agnostic trained innate immunity mechanisms^22^ and bystander activation (non-SARS- CoV-2 specific) of virtual memory (VM) and VM-like CD8+ T cells^23^. As such, any immune challenge may establish new baseline immune statuses with the potential to impact future responses in both antigen-specific and antigen-agnostic ways^24^. In this regard, our findings add further insight to these mechanisms as we could also predict vaccine failure even before vaccination.

In addition to the inherent properties of vaccines, other factors can contribute to their overall effectiveness, such as gender, age, co-morbidities, pre-existing diseases or socio-economic background^25–28^. Hence, throughout the lifespan, sex and age are fundamental transformers of immunity to infectious diseases and to vaccination’s responses^29^. Females tend to mount stronger immune responses than males^30^, and immunosenescence leads to impaired immune function and a heightened inflammatory state in older adults^31^. There is an important intersection between these host factors, whereby the impact of aging on the immune system differs in males and females^32^. The implications of the interaction between sex and age were clearly demonstrated by the epidemiology and clinical manifestations of respiratory viral diseases, such as influenza and COVID-19. In this latter case, sex differences in immune responses were shown to underlie COVID-19 disease outcomes^33^. Female bias in immunity to influenza vaccines has been consistently reported, however understanding of sex differences in the response to COVID-19 vaccines in older adults is incomplete due to small sample sizes and failure to disaggregate clinical trial data by both sex and age^34^. Nevertheless, in our case there were not enough data to segregate data on sex, an issue to be addressed in future studies.

In summary, we here have proved that although the vaccine-induced humoral and cellular memory cannot predict subsequent infection in immune-compromised patients, an unbiased characterization of the circulating immunome can predict it even before vaccination. Hence, future studies should expand this pilot study focusing of the relevance of the already identified cell populations which seem to play a pivotal role controlling SARS-CoV-2 infection, with a particular input on the CD45RA^+^CCR7^-^ Tγδ subsets.

## Data Availability

All data produced in the present study are available upon reasonable request to the authors

## SUPPLEMENTARY FIGURES

**Supplementary figure 1:**
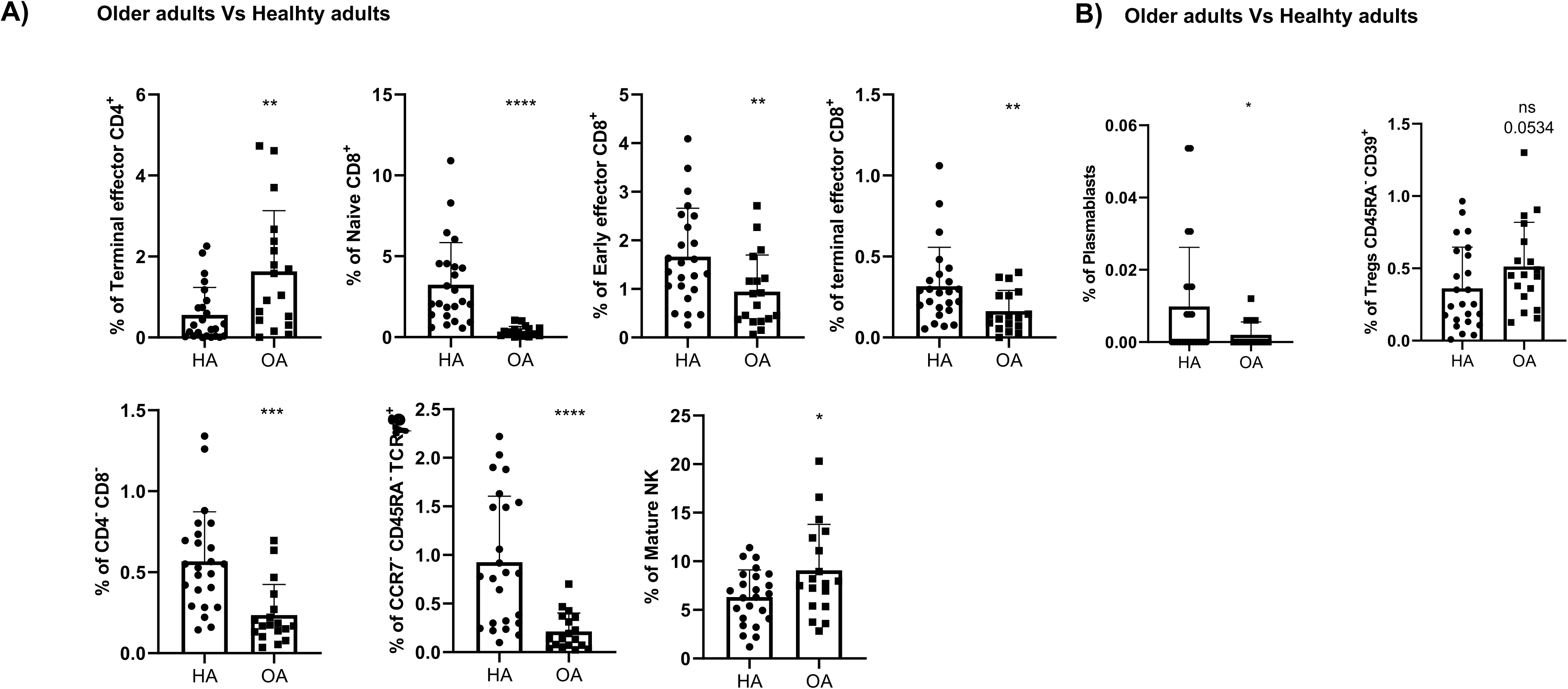
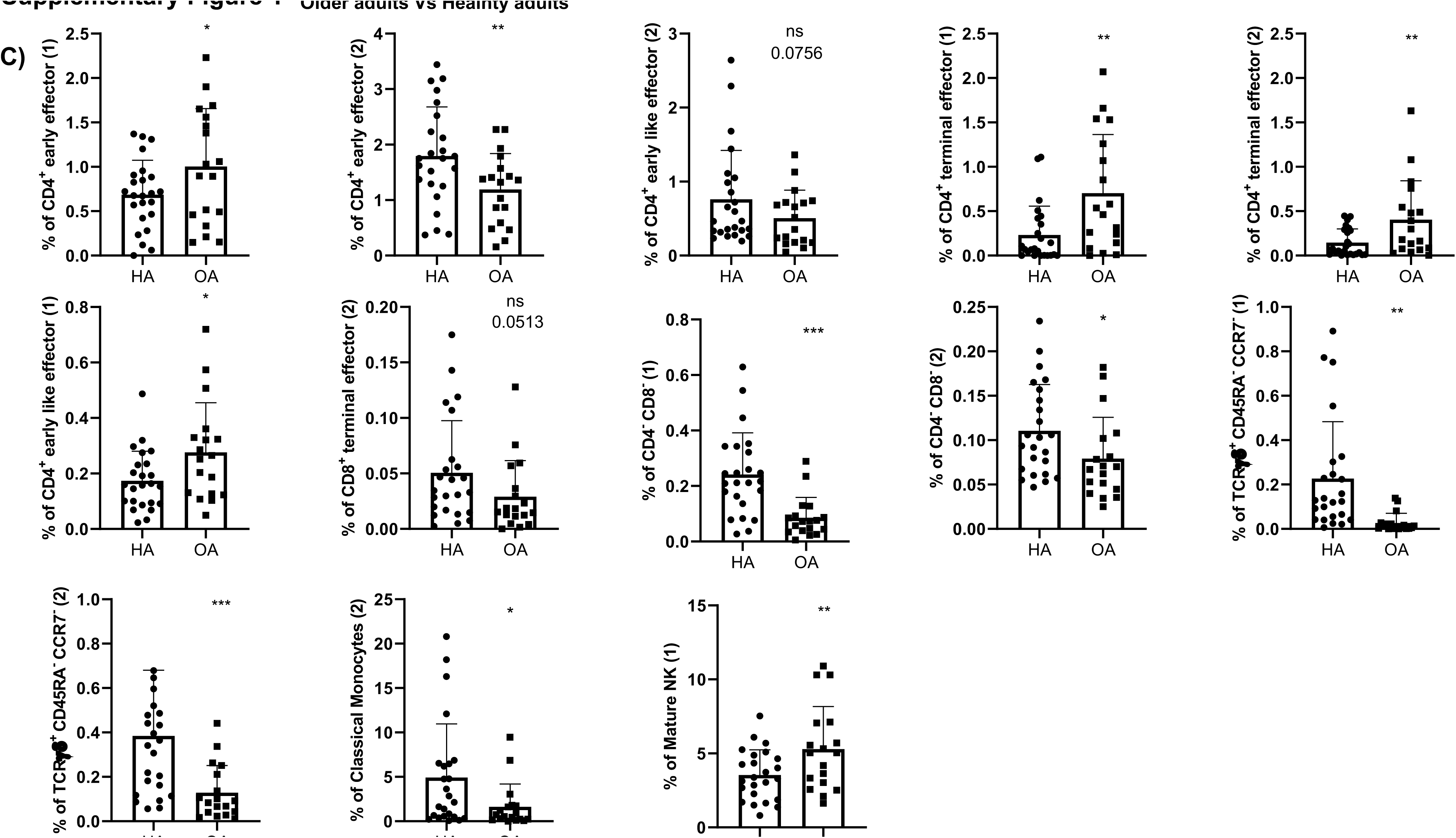
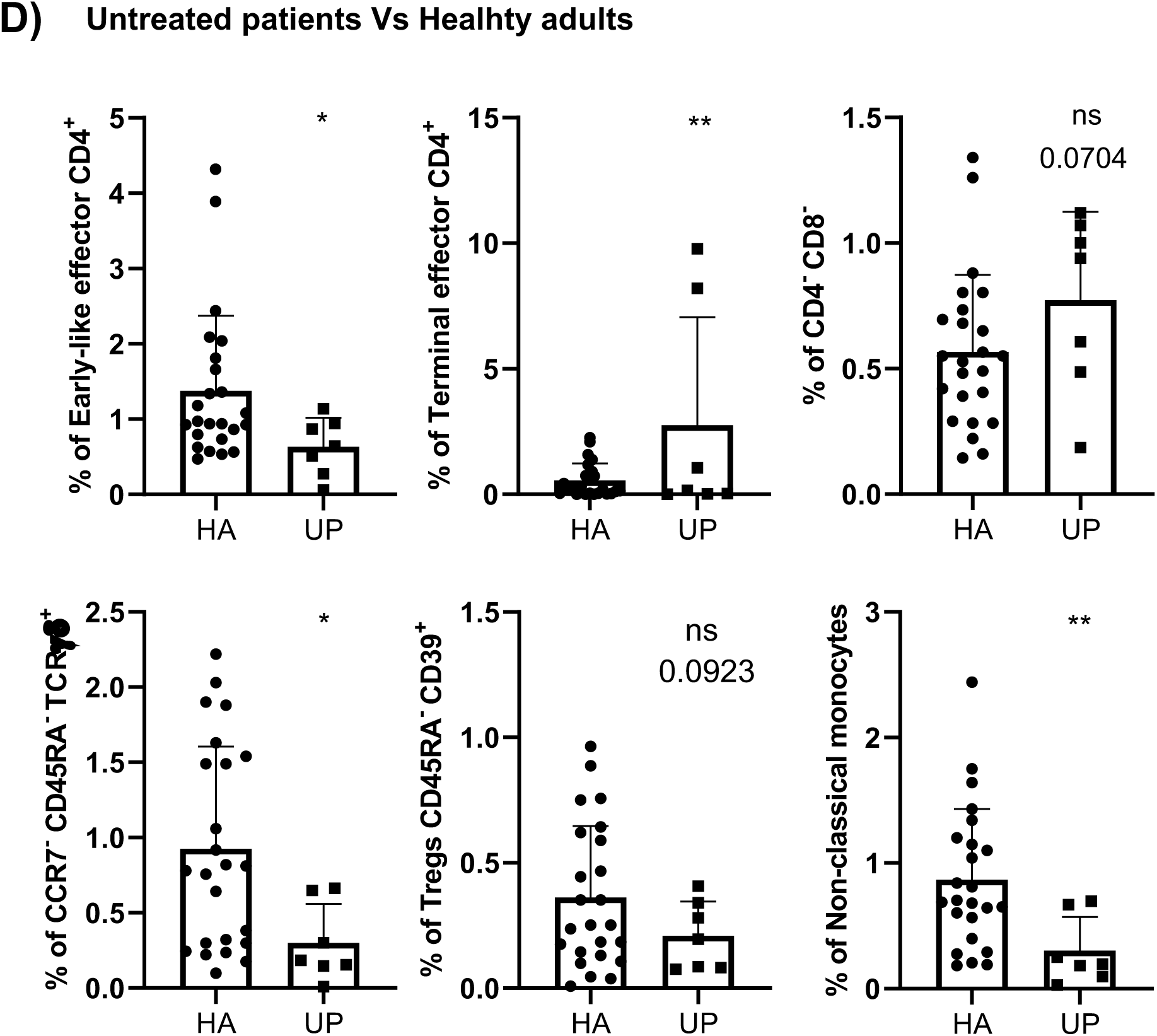
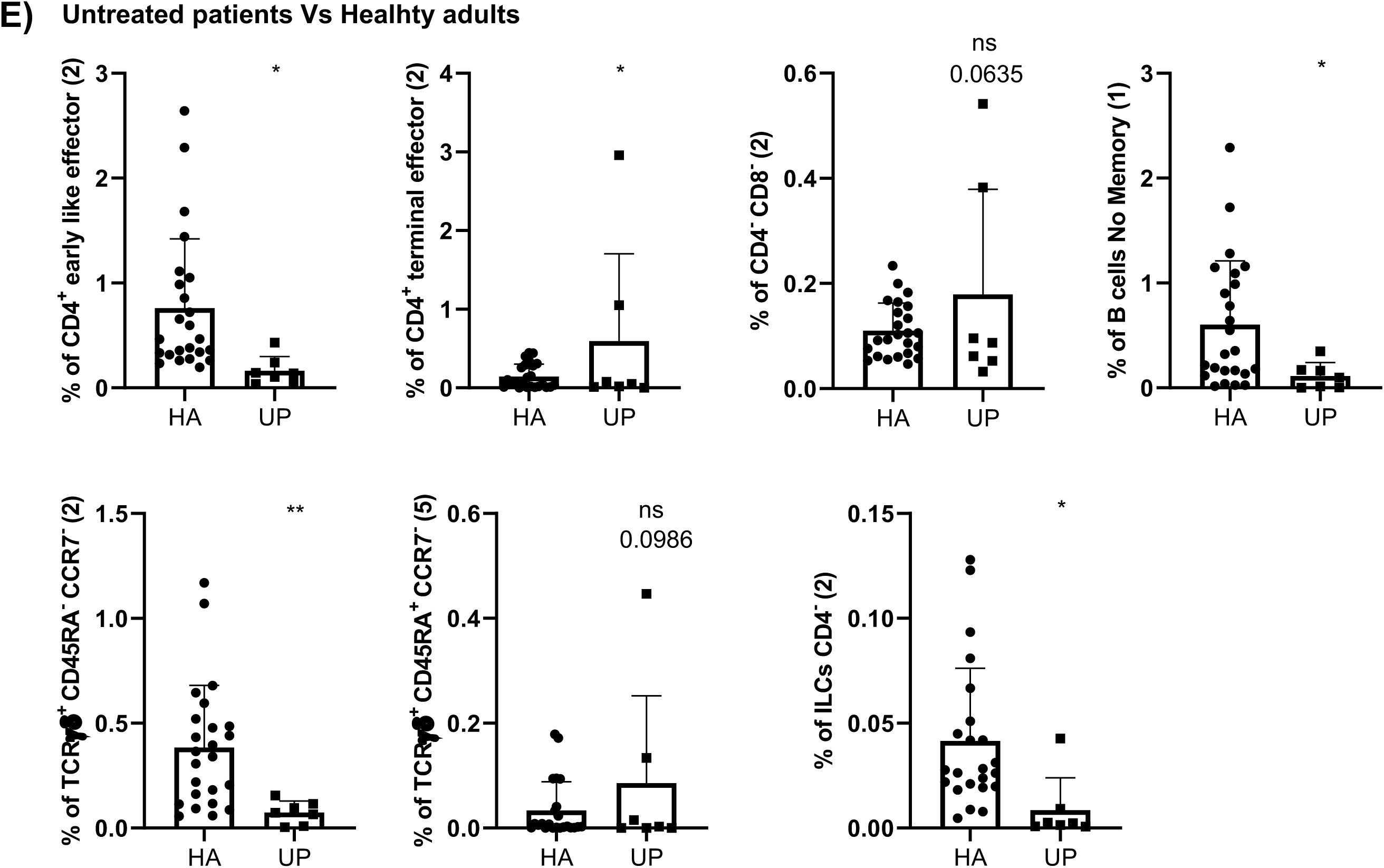
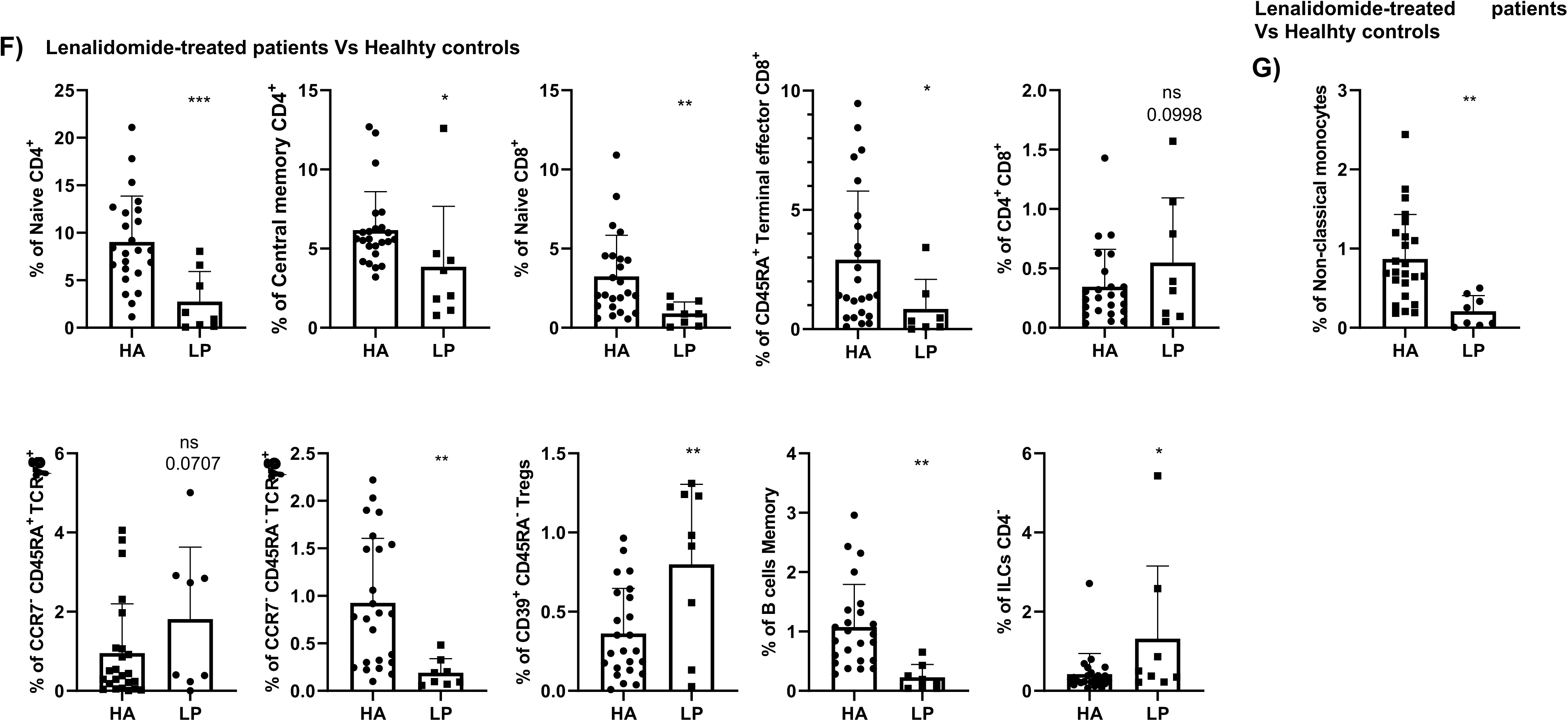
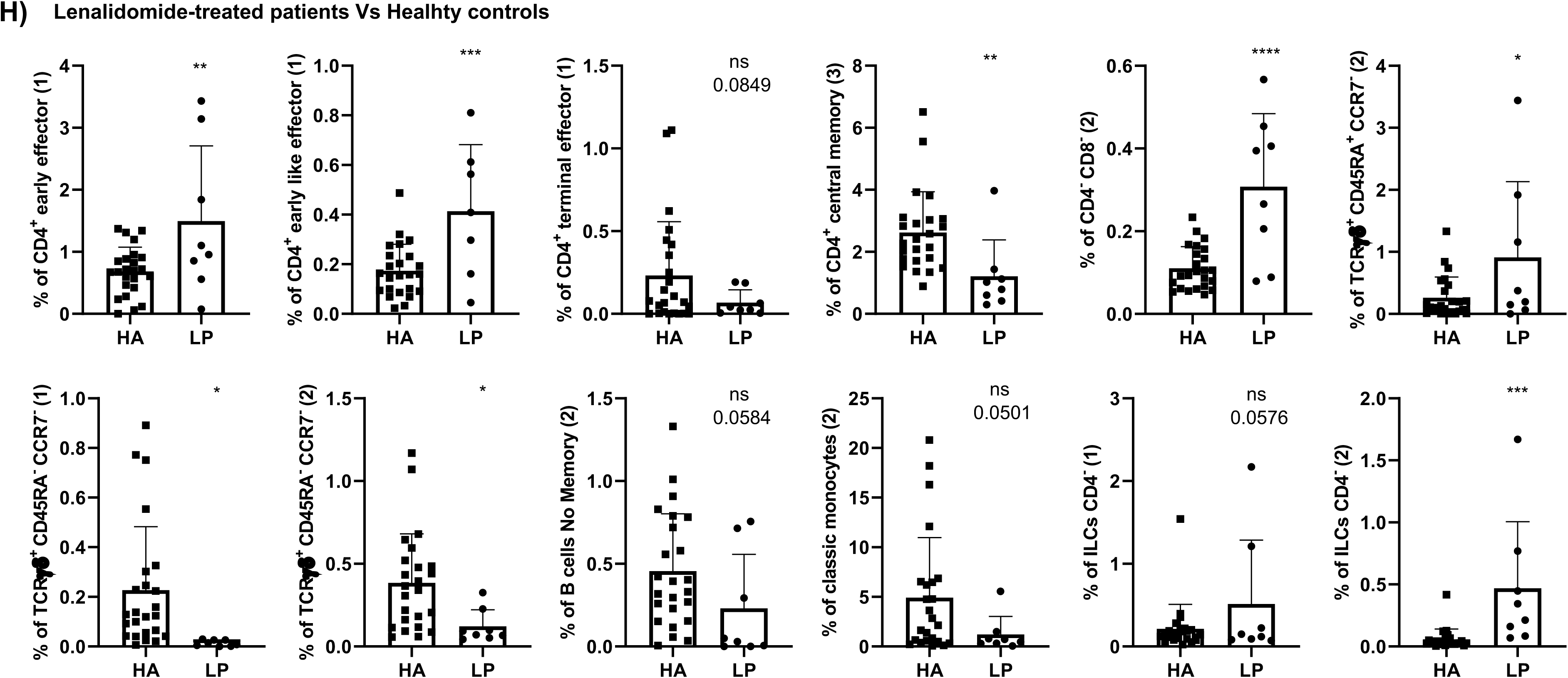
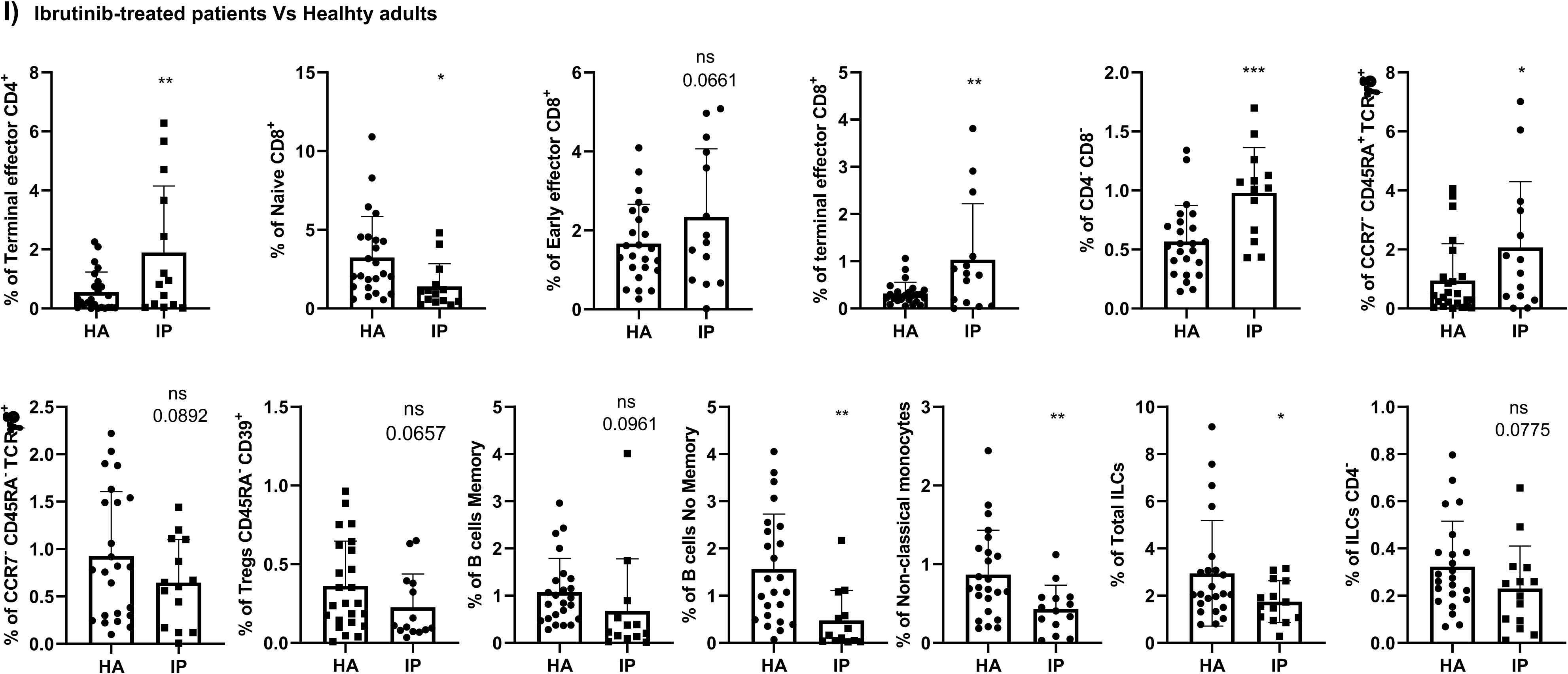
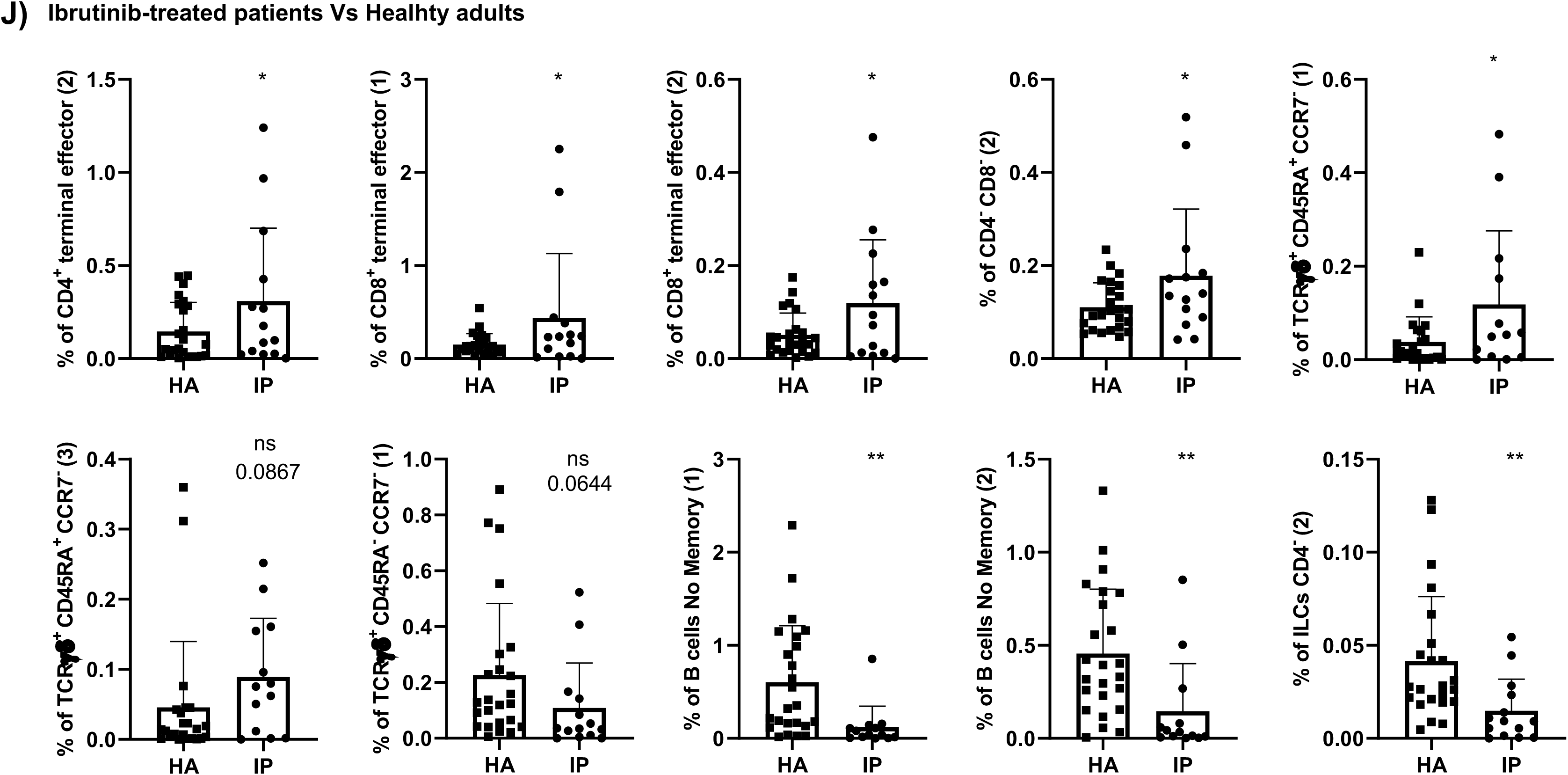
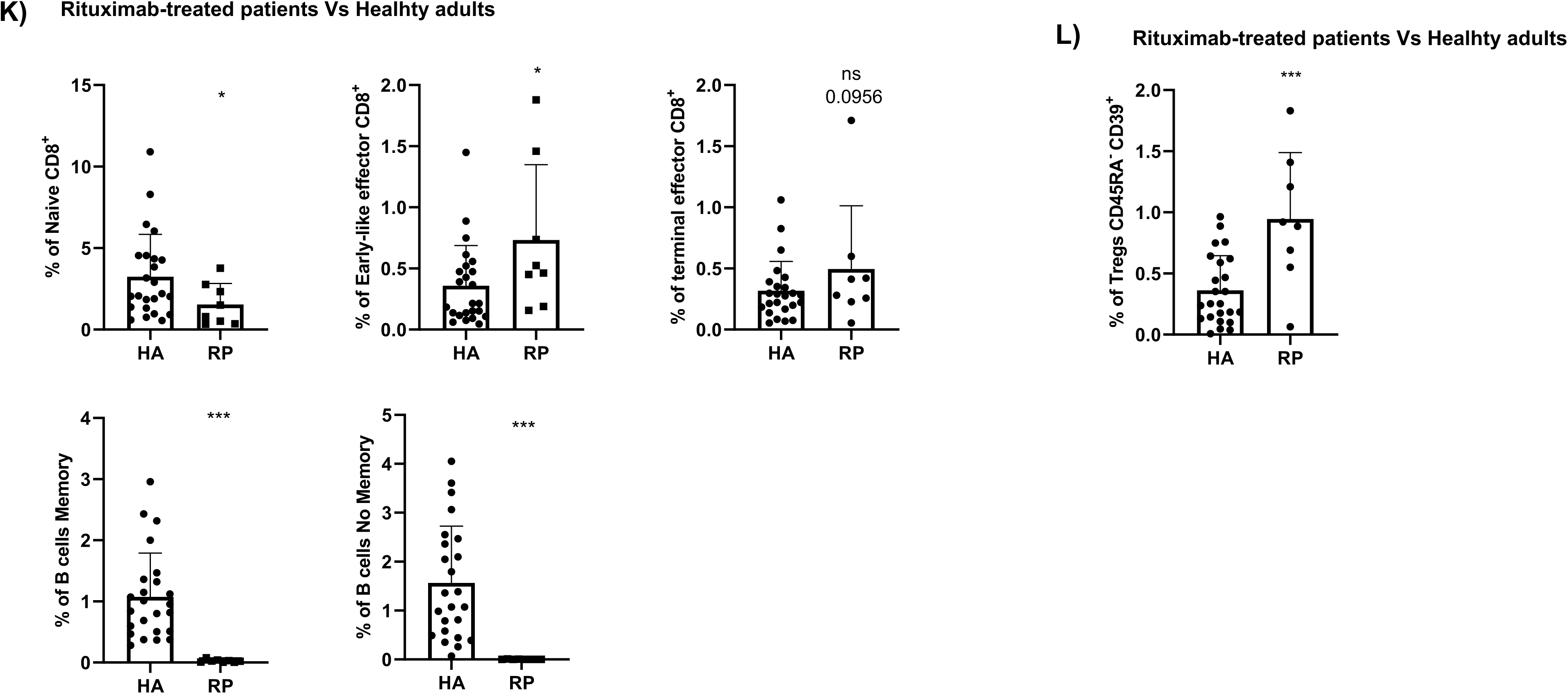
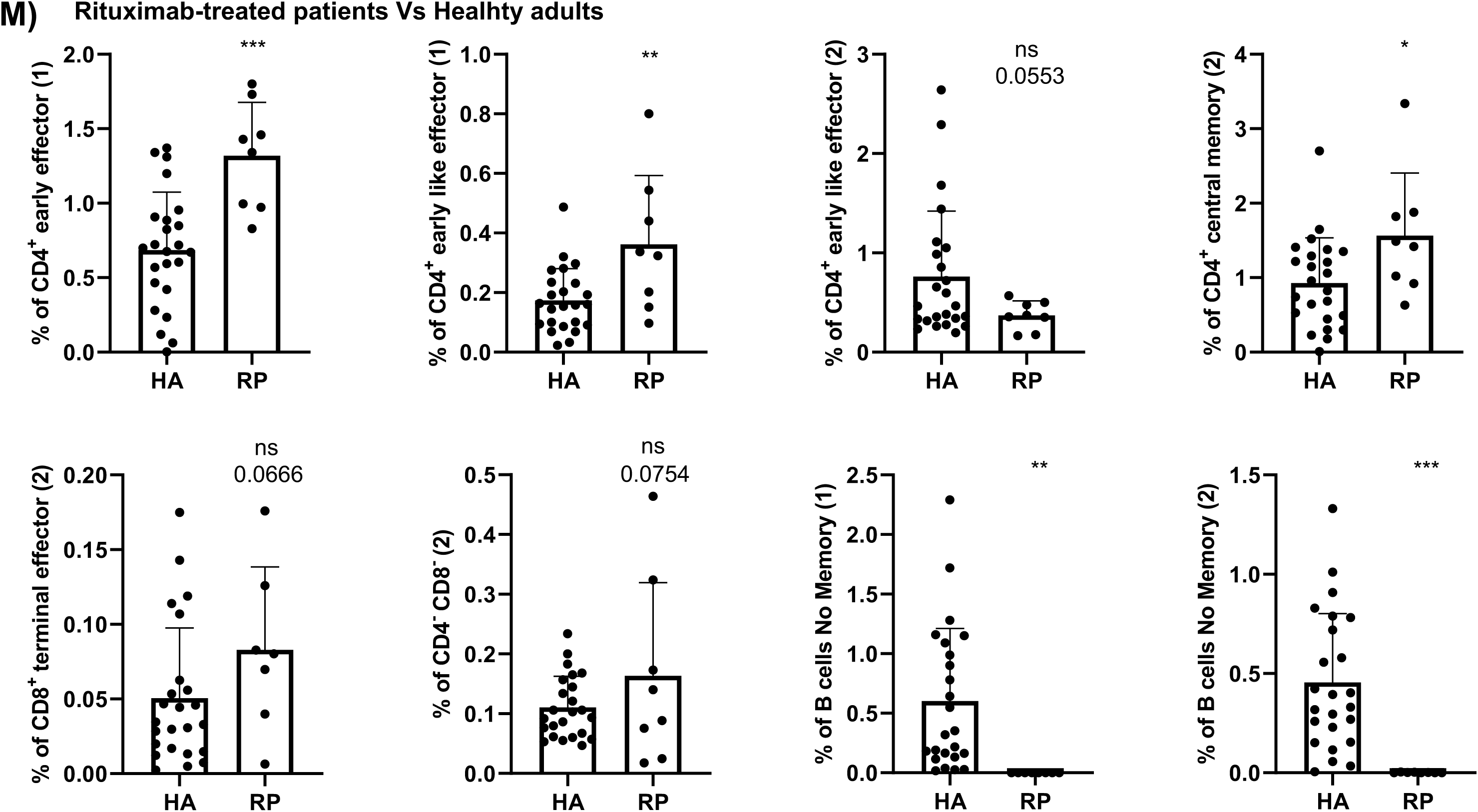
In depth analysis of the different cohorts at baseline. **A)** Validation by hierarchical gating, as shown in Figure 2, of the cell populations differentially expressed between healthy adults (HA) and older adults (OA)in the in the volcano plot from Figure 3C. Those samples that could not be analyzed by the volcano plots due the low number of events are shown in **B).** Analysis of the specific cluster subsets within a given population is shown in **C)**. **D)** Comparison by hierarchical gating of the populations differentially expressed between HA and untreated oncohematologic patients (UP) as revealed in Figure 3D. The specific cluster subset comparison is shown in **E)**. **F)** Study of the cell populations differentially expressed between HA and lenalidomide-treated oncohematologic patients (LP) as shown in Figure 3E. Those samples that could not be analyzed by the volcano plots due the low number of events are shown in **G).** Analysis of the specific cluster subsets within a given population is shown in **H)**. **I)** Comparison by hierarchical gating of the cell populations differentially expressed between HA and ibrutinib-treated oncohematologic patients (IP) as shown in Figure 3F. Analysis of the specific cluster subsets within a given population is shown in **J)**. **K)** Validation by hierarchical gating of the cell populations differentially expressed between HA and rituximab-treated oncohematologic patients (RP) as shown in Figure 3G. Those samples that could not be analyzed by the volcano plots due the low number of events are shown in **L).** Analysis of the specific cluster subsets within a given population is shown in **M)**. In all cases, a t-test analysis was performed where a p-value <0.05 (*<0.05; **<0.01; ***<0.001) was considered significant. A p-value between 0.05 and 0.1 was considered as not significant (ns) but with a relevant trend displaying the p- value underneath.

**Supplementary figure 2:**
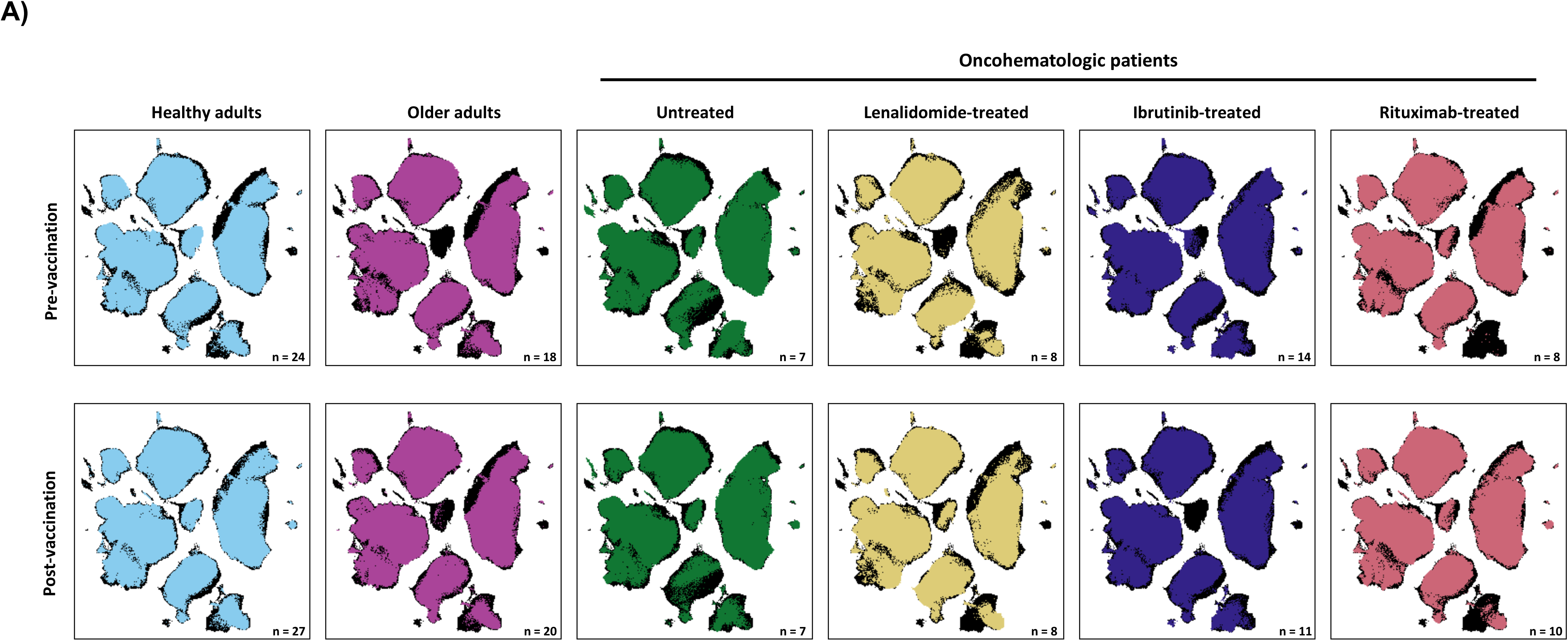
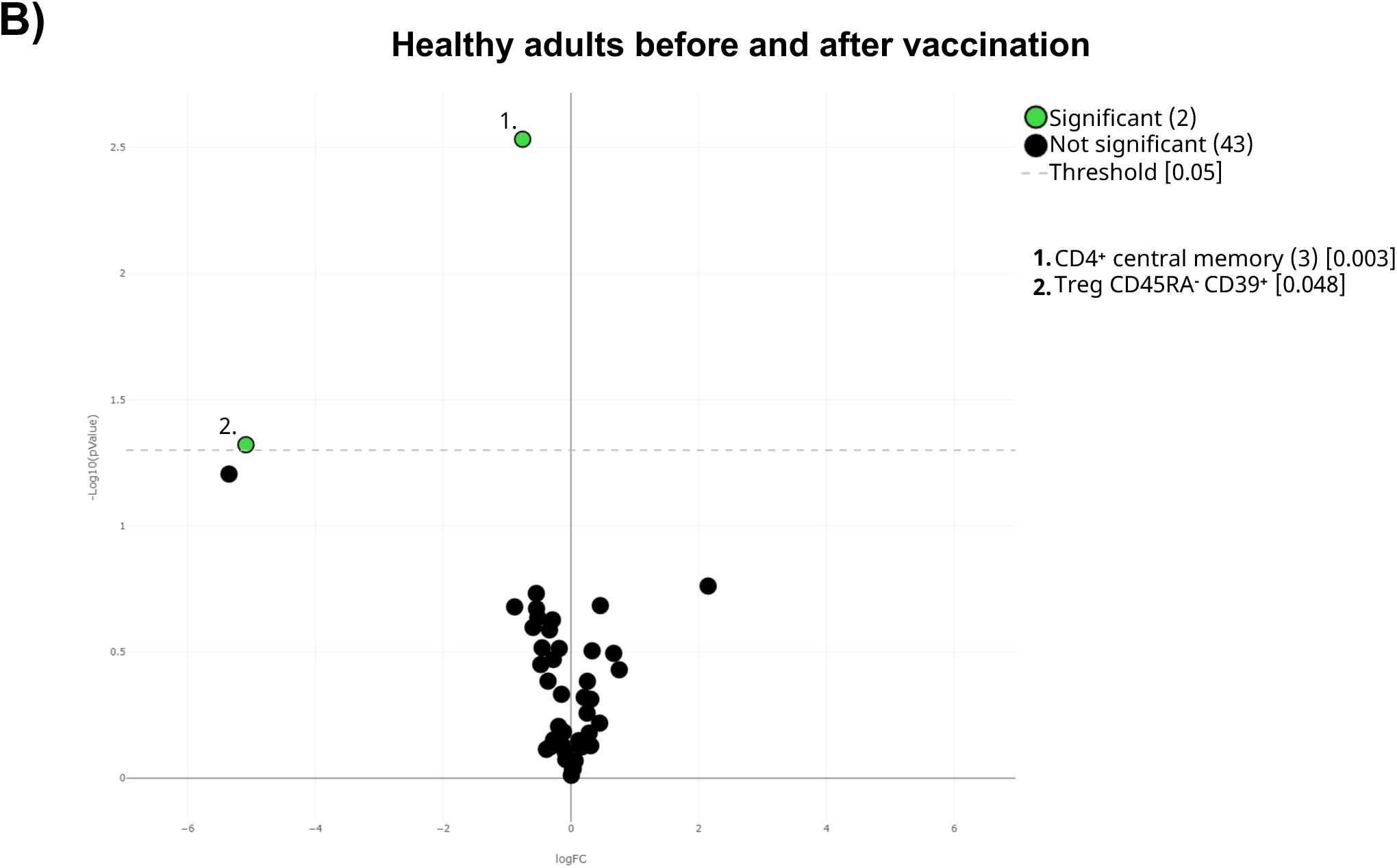
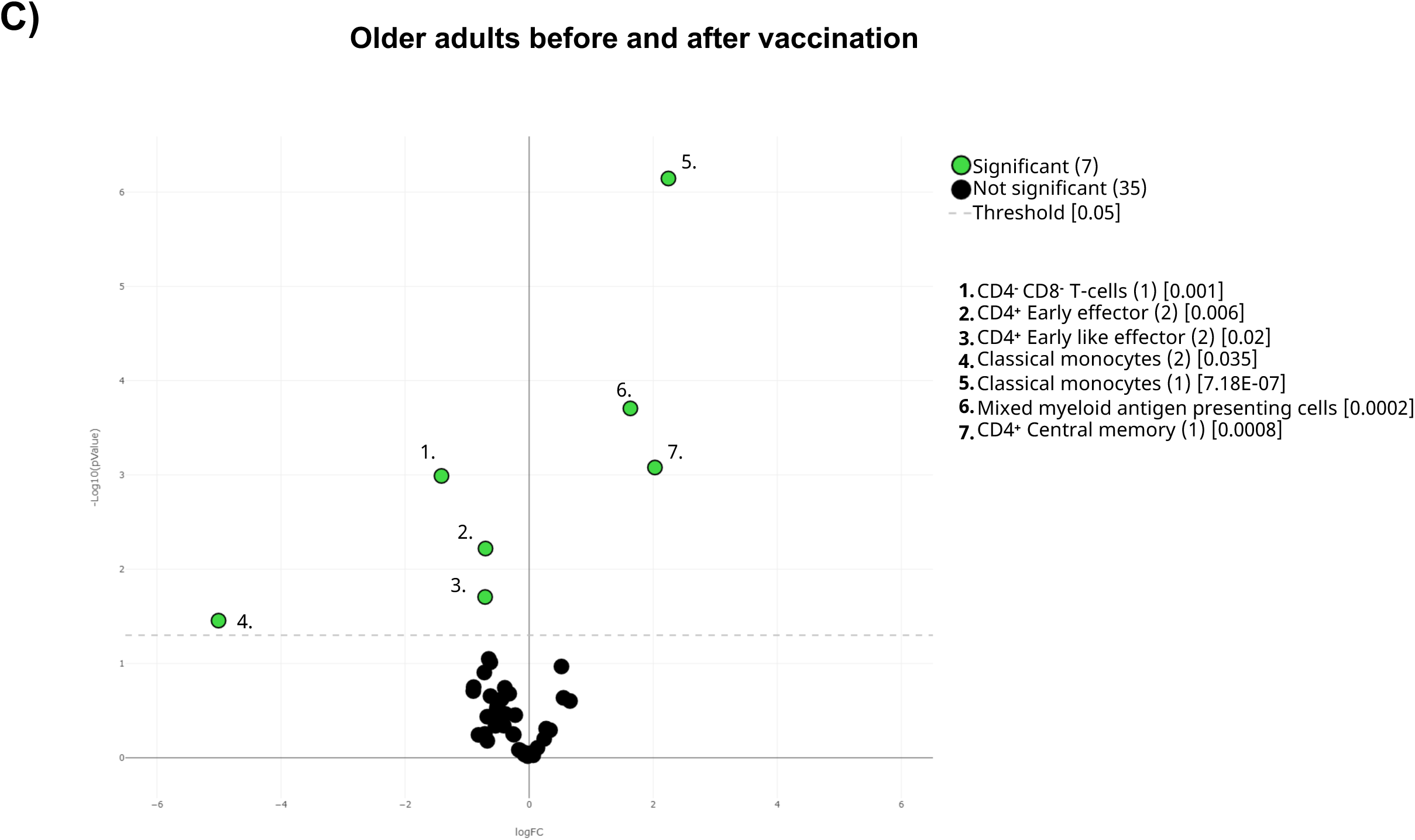
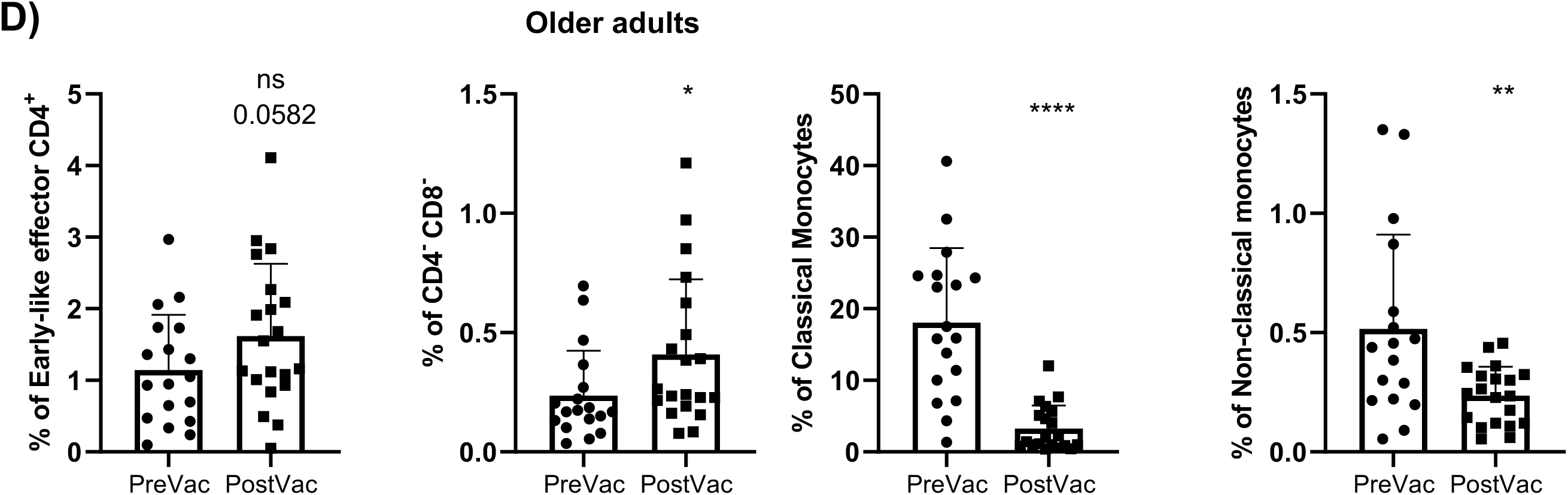
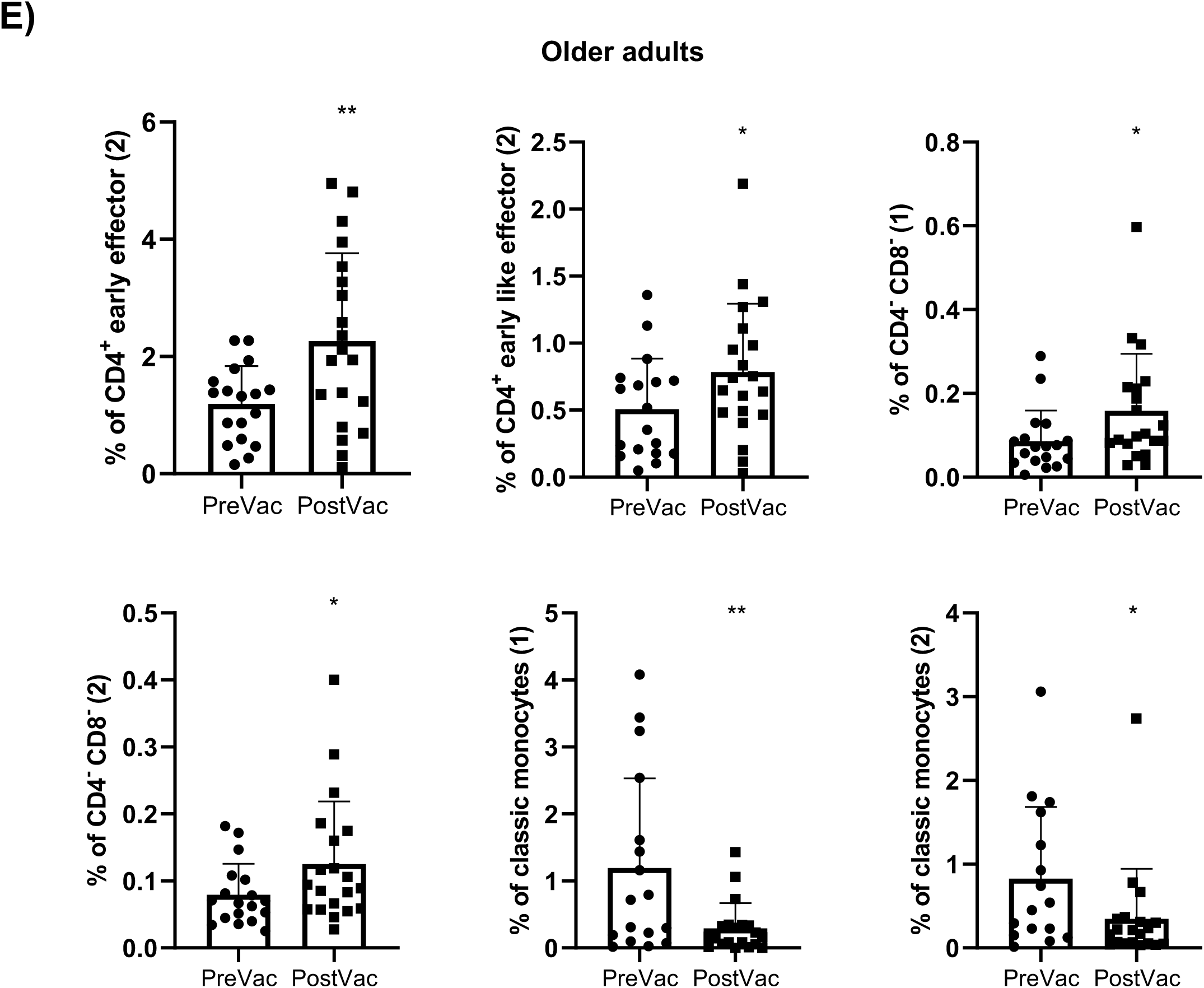
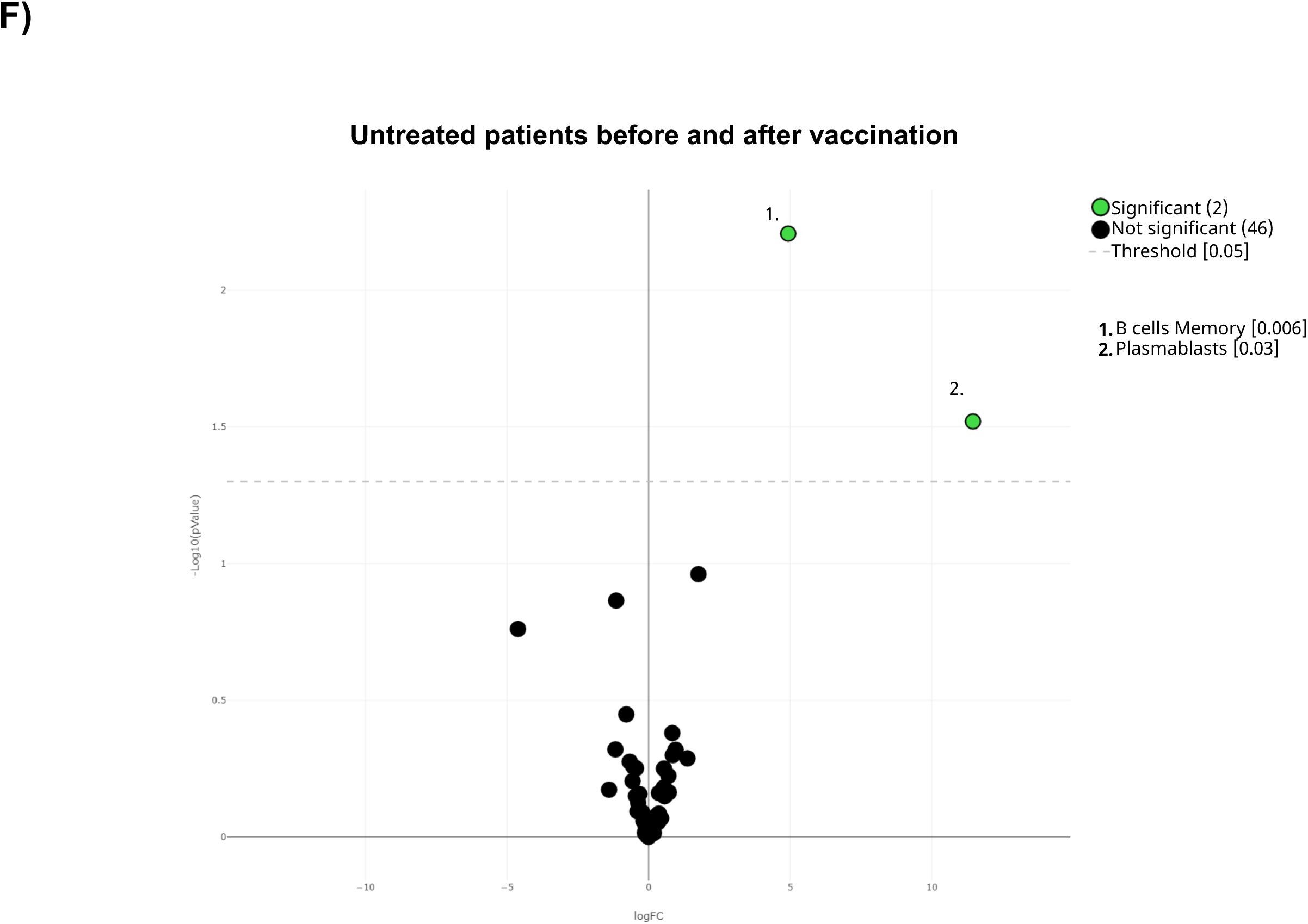
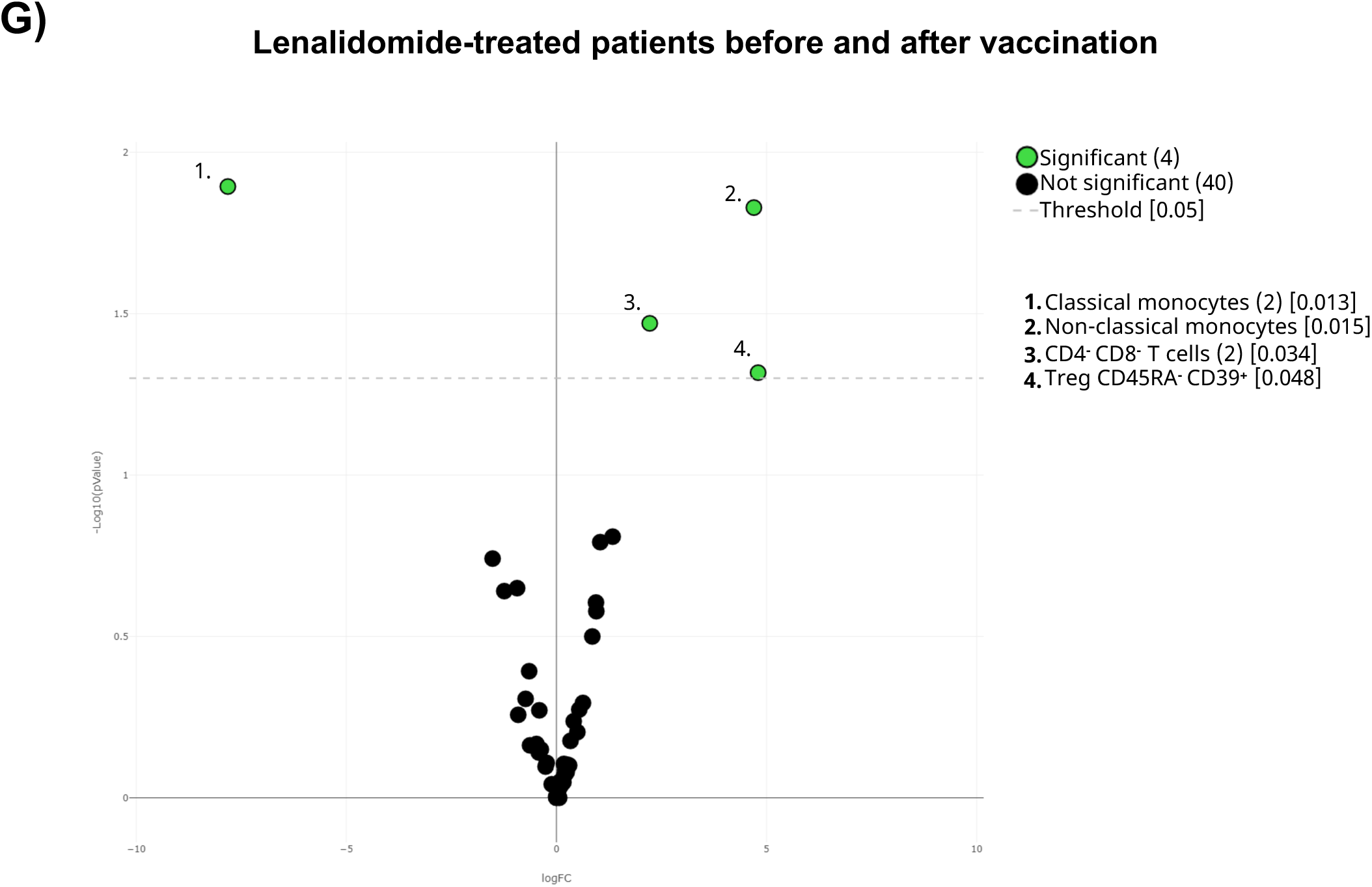
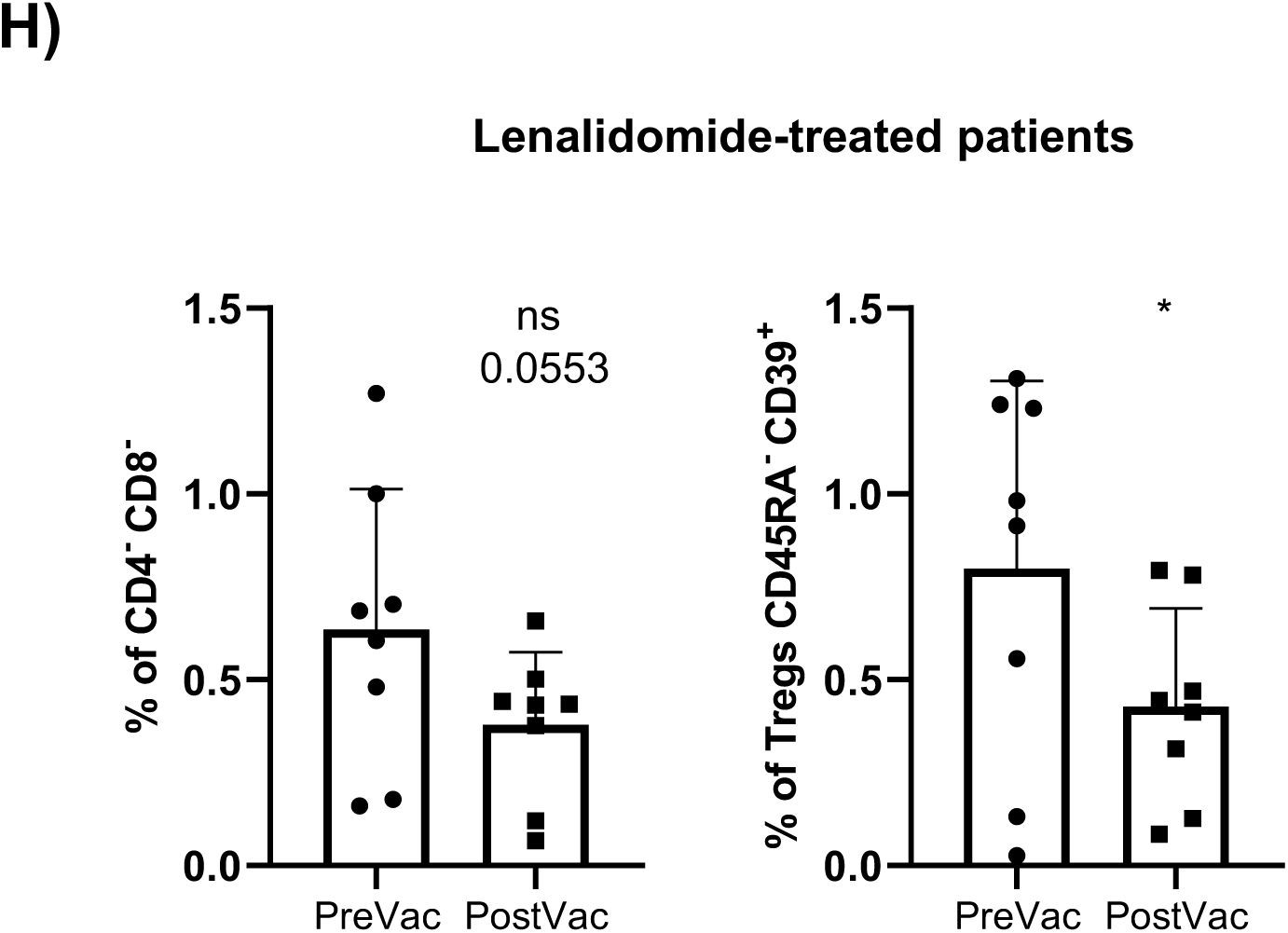
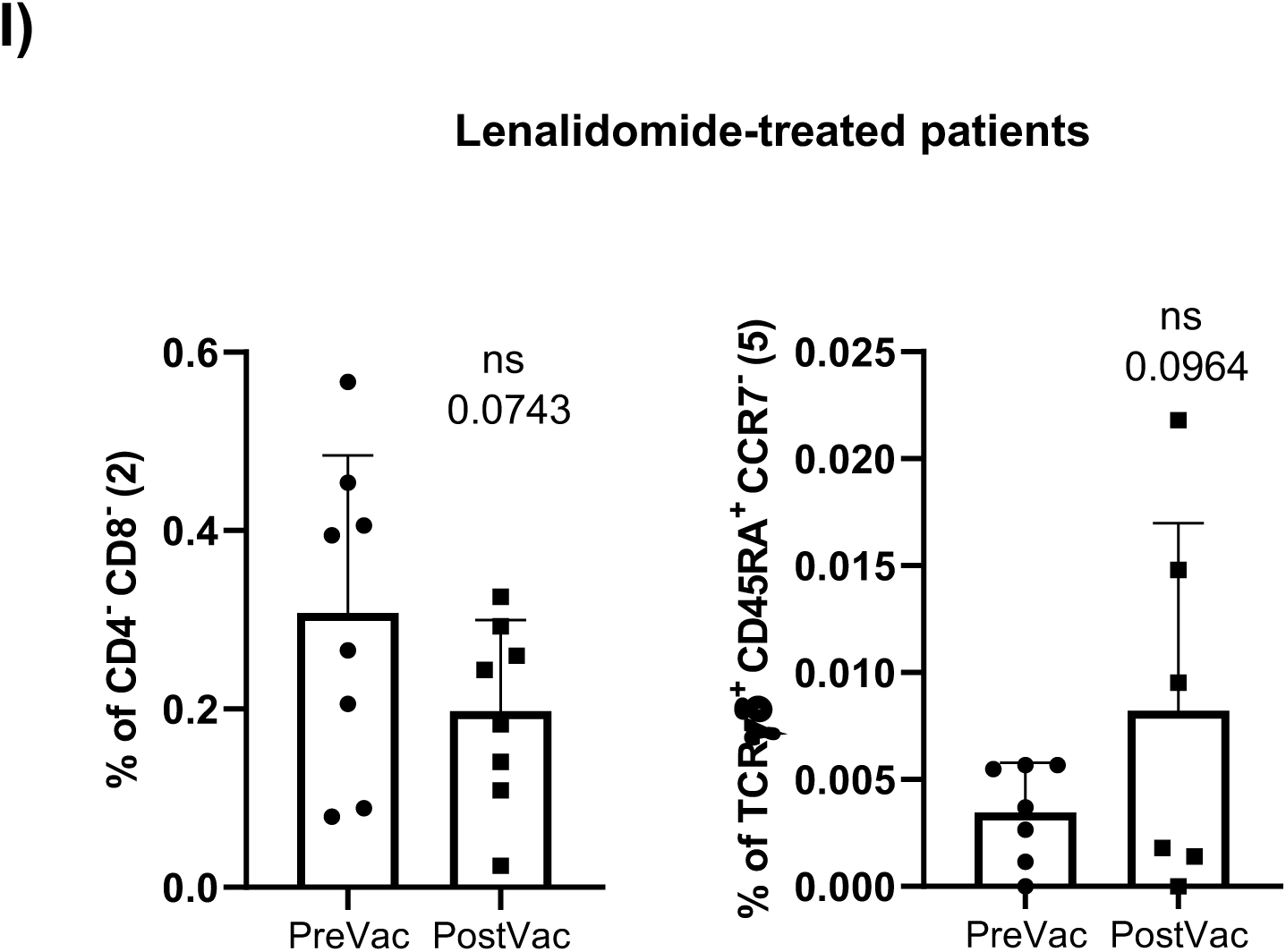
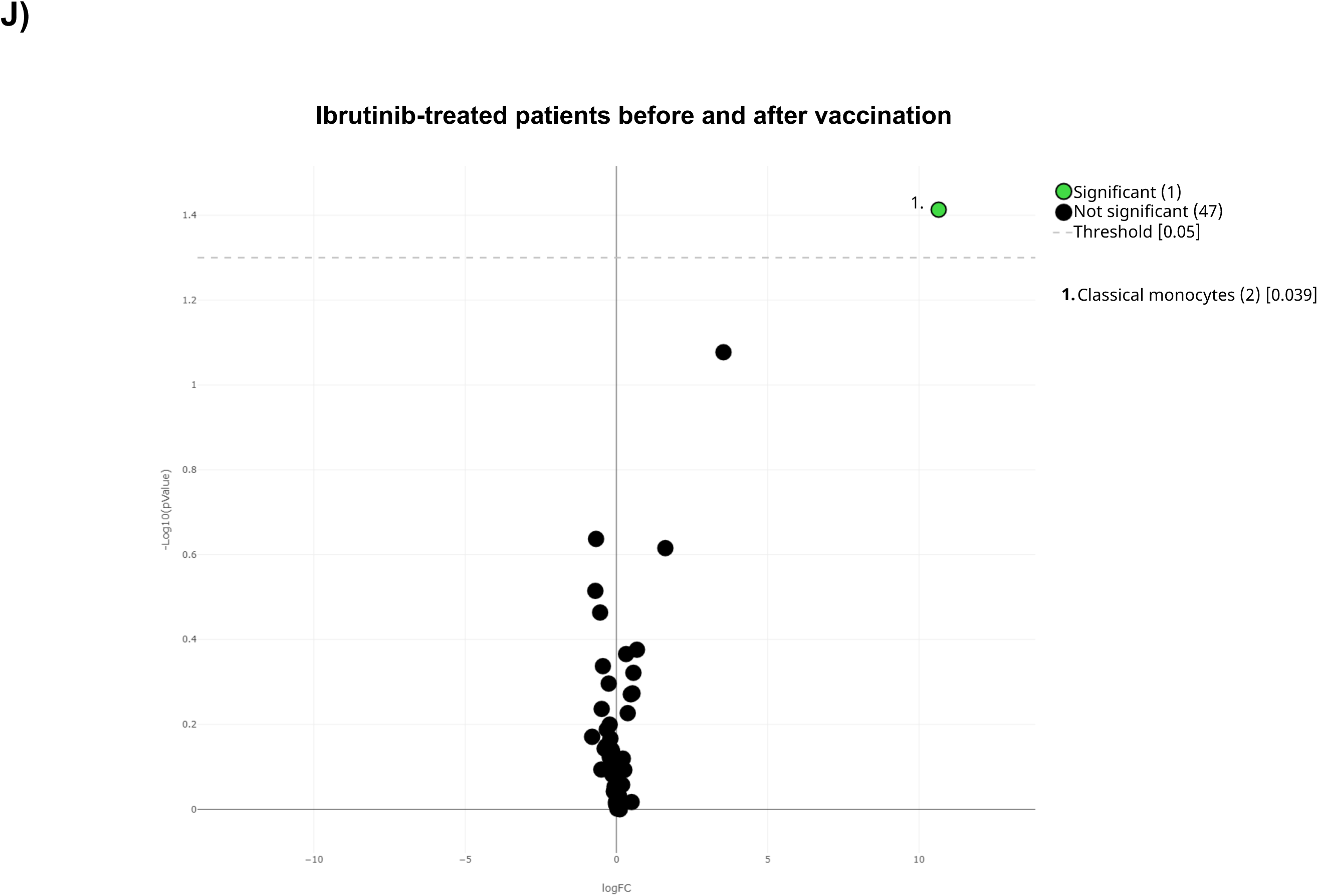
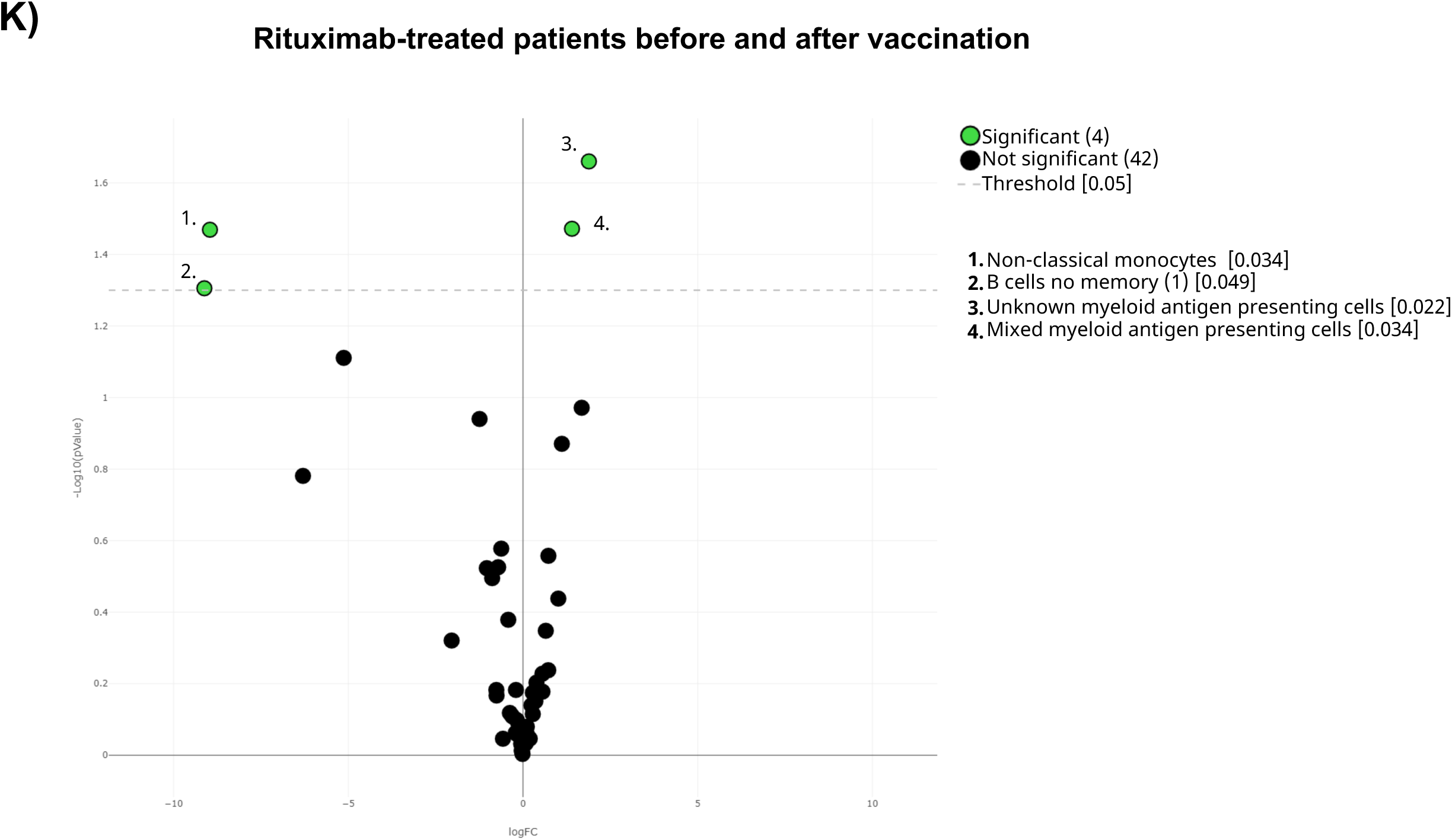
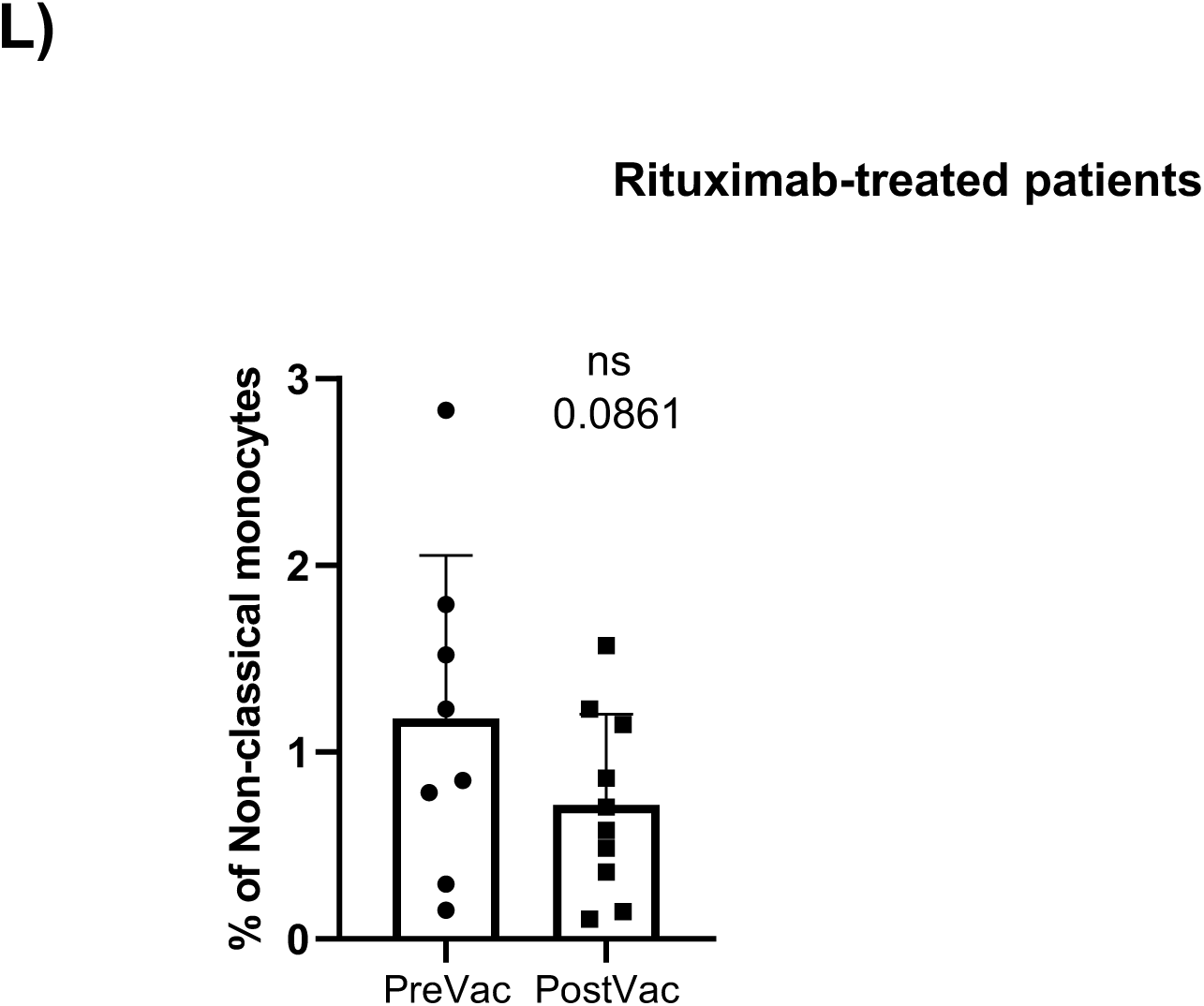
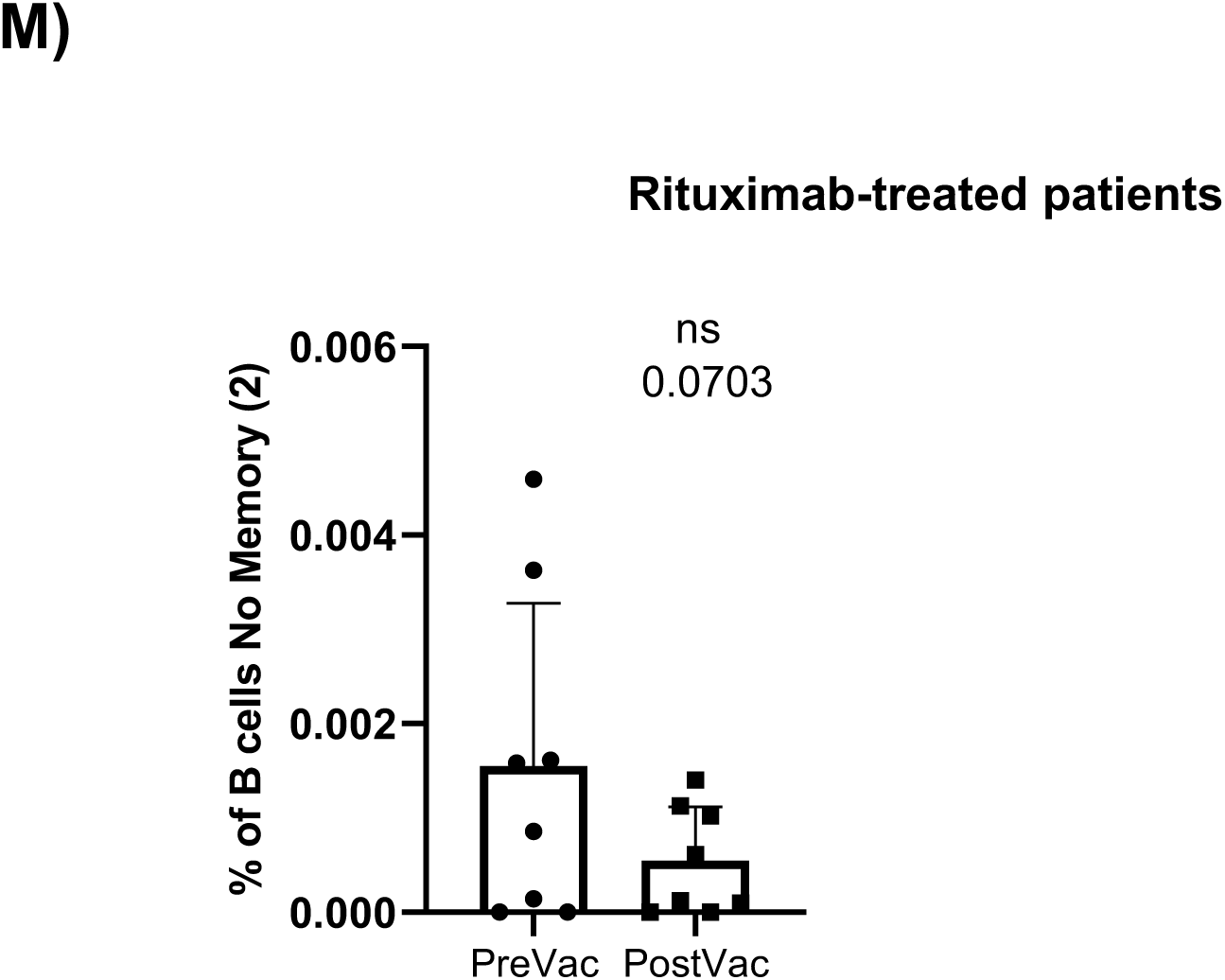
In deep immune characterization of the cohorts before and after vaccination. **A)** UMAP plot of the cohorts before (above) and after (below) vaccination colored by blue (healthy adults), purple (older adults), green (untreated oncohematologic patients), yellow (lenalidomide-treated oncohematologic patients), navy blue (ibrutinib-treated oncohematologic patients) and light red (rituximab-treated oncohematologic patients) referred to all samples (shown in black). **B)** Volcano plot analysis comparing healthy adults before and after vaccination. Clusters that could not be analyzed due the small number of events (SNE) were plasmablasts, CD4^-^/CD8^-^ T cells (2), and Tγδ CD45RA^+^/CCR7^-^ (4). **C)** Volcano plot analysis comparing Older adults before and after vaccination. Clusters that could not be analyzed due to SNE were non-classical monocytes, plasmablasts, CD4^-^/CD8^-^ T cells (2), Tγδ CD45RA^+^/CCR7^-^ (4), Tγδ CD45RA^+^/CCR7^-^ (5) and T_regs_ CD45RA^-^/CD39^+^. **D)** Classical validation, as in Figure 2, of the populations significantly expressed in the volcano plot from C and **E)** the specific cell cluster subsets. **F)** Volcano plot analysis comparing Untreated patients before and after vaccination. **G)** Volcano plot analysis comparing Lenalidomide-treated patients before and after vaccination. Clusters that could not be analyzed due to SNE were plasmablasts, Tγδ CD45RA^+^/CCR7^-^ (4), Tγδ CD45RA^+^/CCR7^-^ (5) and B cells no memory (2). **H)** Classical validation of the populations significantly expressed in the volcano plot from G) and **I)** their specific cell cluster subsets. **J)** Volcano plot analysis comparing Ibrutinib-treated patients before and after vaccination. **K)** Volcano plot analysis comparing Rituximab-treated patients before and after vaccination. Clusters that could not be analyzed due to SNE were plasmablasts and B cells no memory (2). **L)** Classical validation of the populations significantly expressed in the volcano plot K) and **M)** the specific cell cluster subsets. In the volcano plots, green dots represent those clusters that showed significant differences (p-value<0.05). A t-test analysis was performed in panels D), E), H), I), L) and M). In all cases, a p-value<0.05 (*<0.05; **<0.01; ***<0.001) was considered significant, and a p-value between 0.05 and 0.1 was considered as not significant (ns) but with a relevant trend displaying the p-value underneath. “PreVac” and “PostVac” refer respectively to the circulating levels of each cell subset before and after vaccination.

**Supplementary figure 3:**
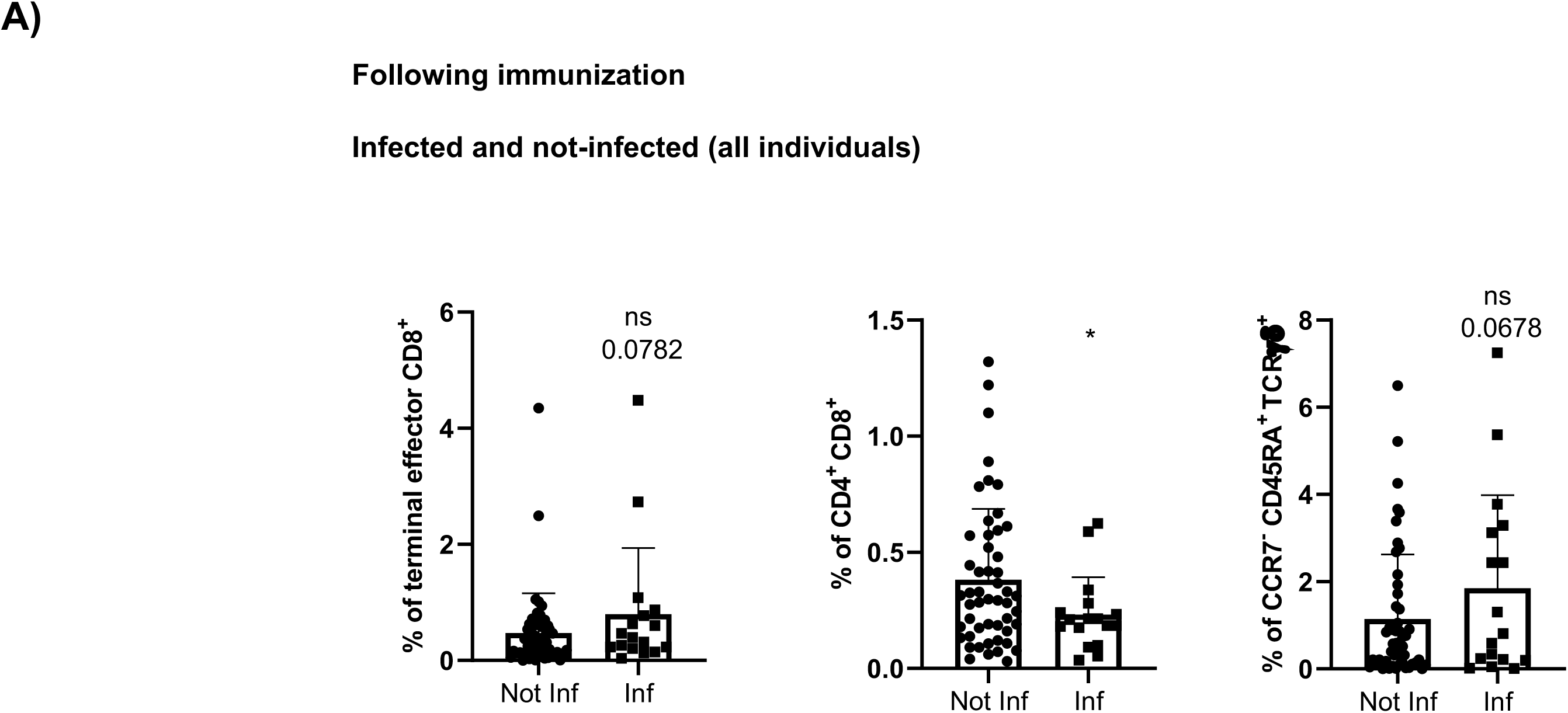
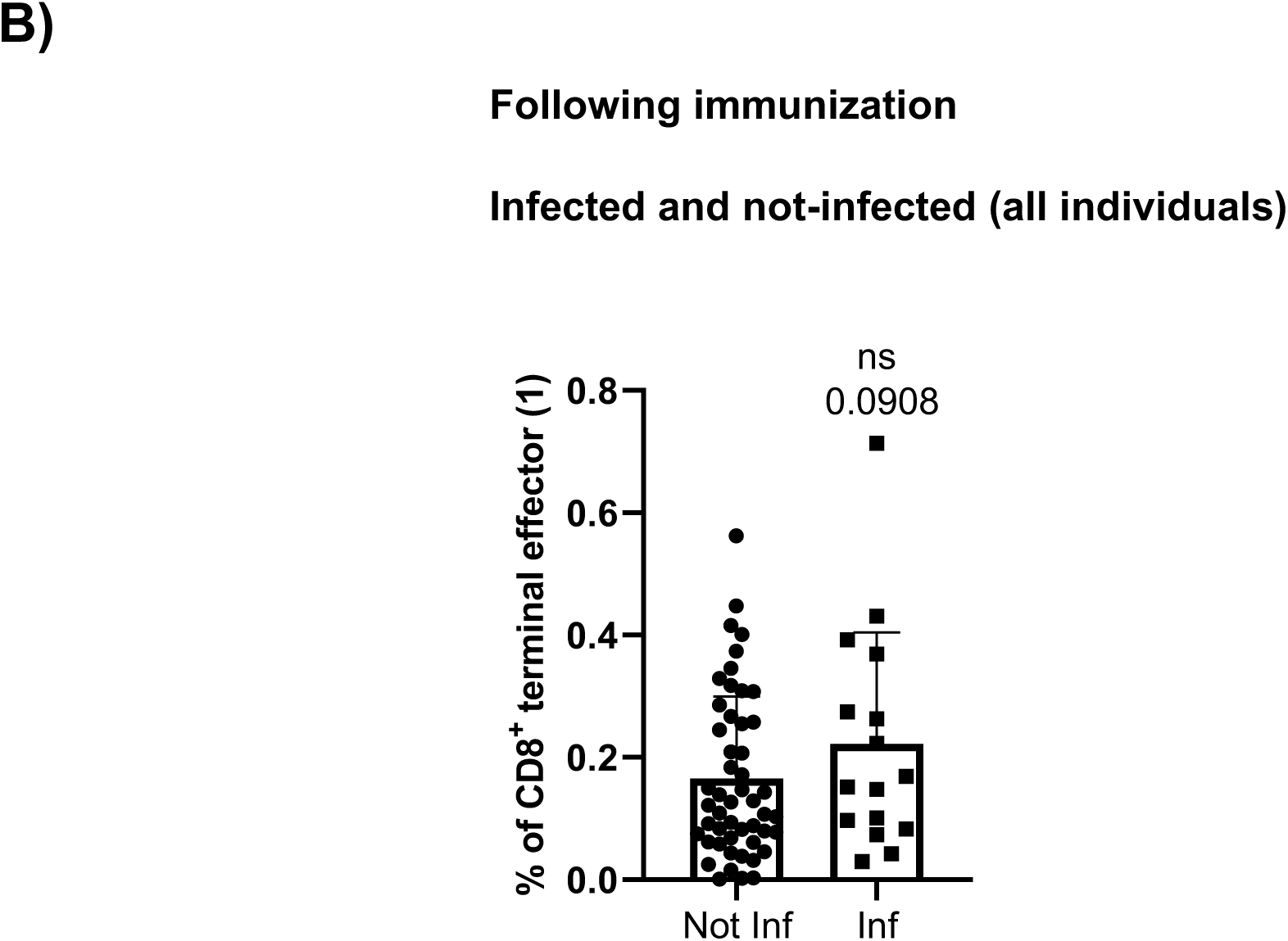
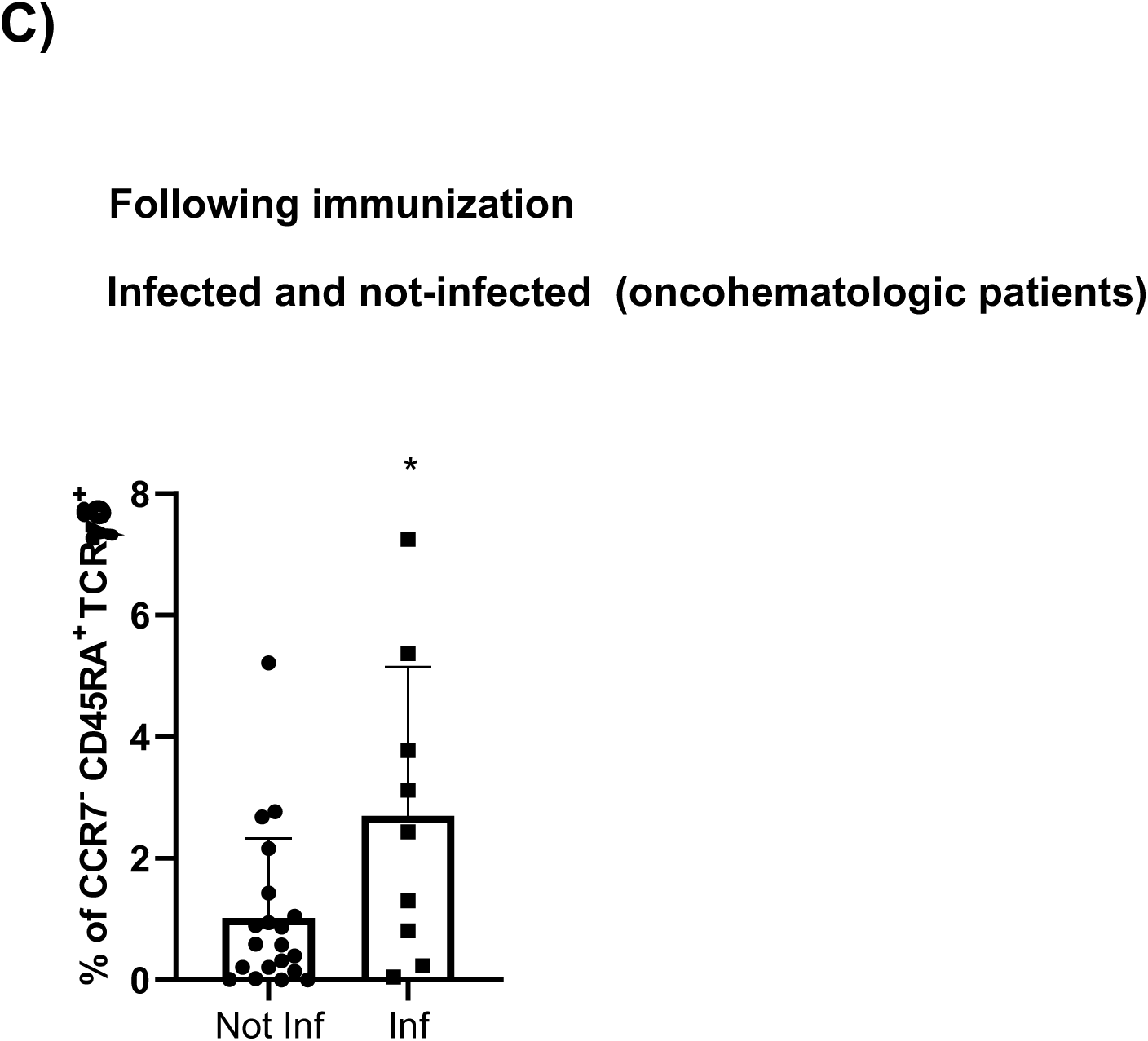
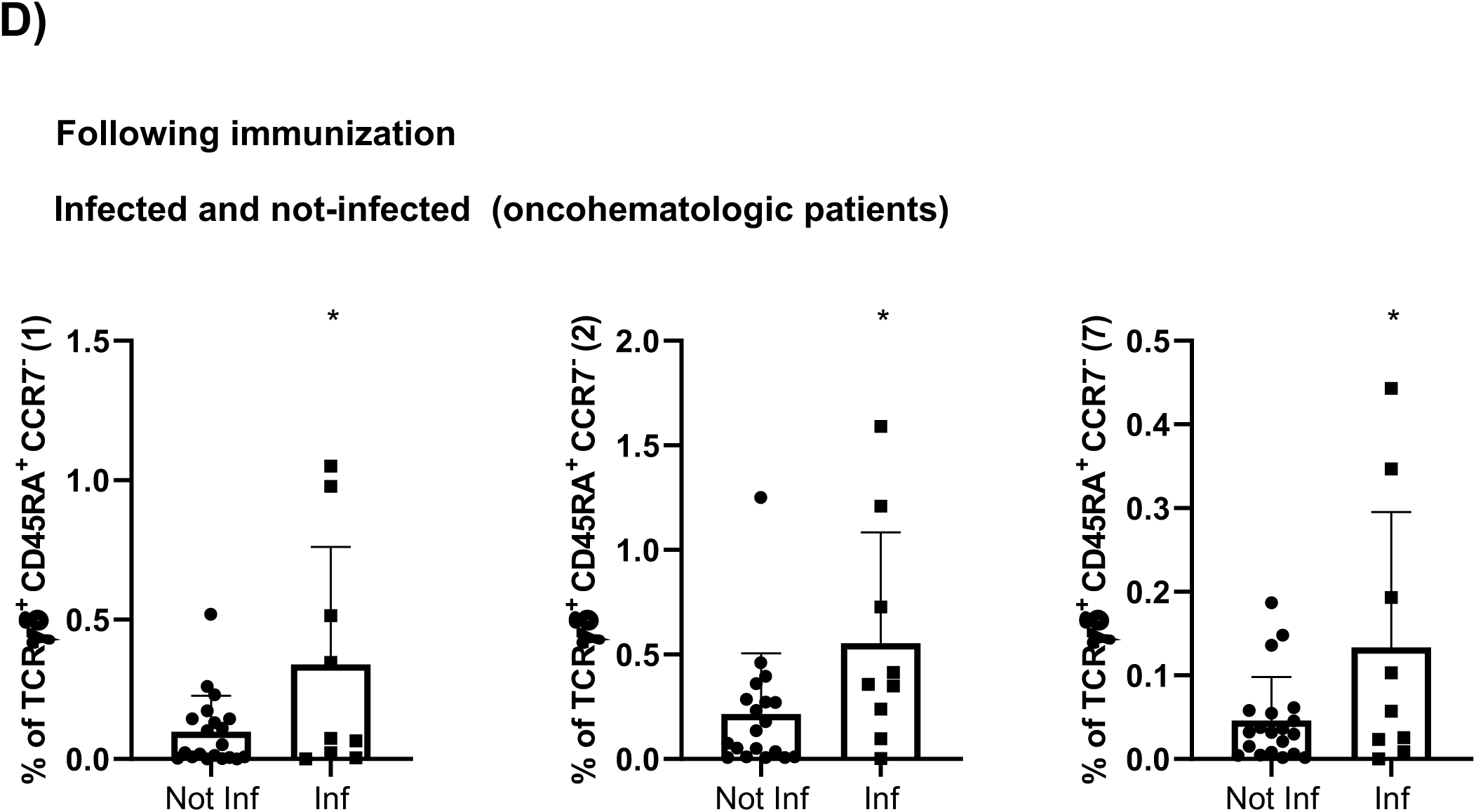
Immune variations following vaccination predicts SARS-CoV-2 infection. **A)** Classical validation, as in Figure 2, of populations significantly expressed in the volcano plot from figure 4B comparing the immune system of all individuals following immunization based on their subsequent infection and **B)** their specific cluster subsets. **C)** Hierarchical gating of the populations significantly expressed in the volcano plot from figure 4E comparing infected and non-infected oncomehatologic patients, and subsequent study in **D)** of the statistically significant cell cluster subsets. In all cases, a t-test analysis was performed where a p-value<0.05 (*<0.05) was considered significant, and a p-value between 0.05 and 0.1 was considered as not significant (ns) but with a relevant trend displaying the p-value underneath. “Inf” refers to subsequently infected individuals, while “Not Inf” are those who were not infected.

**Supplementary figure 4:**
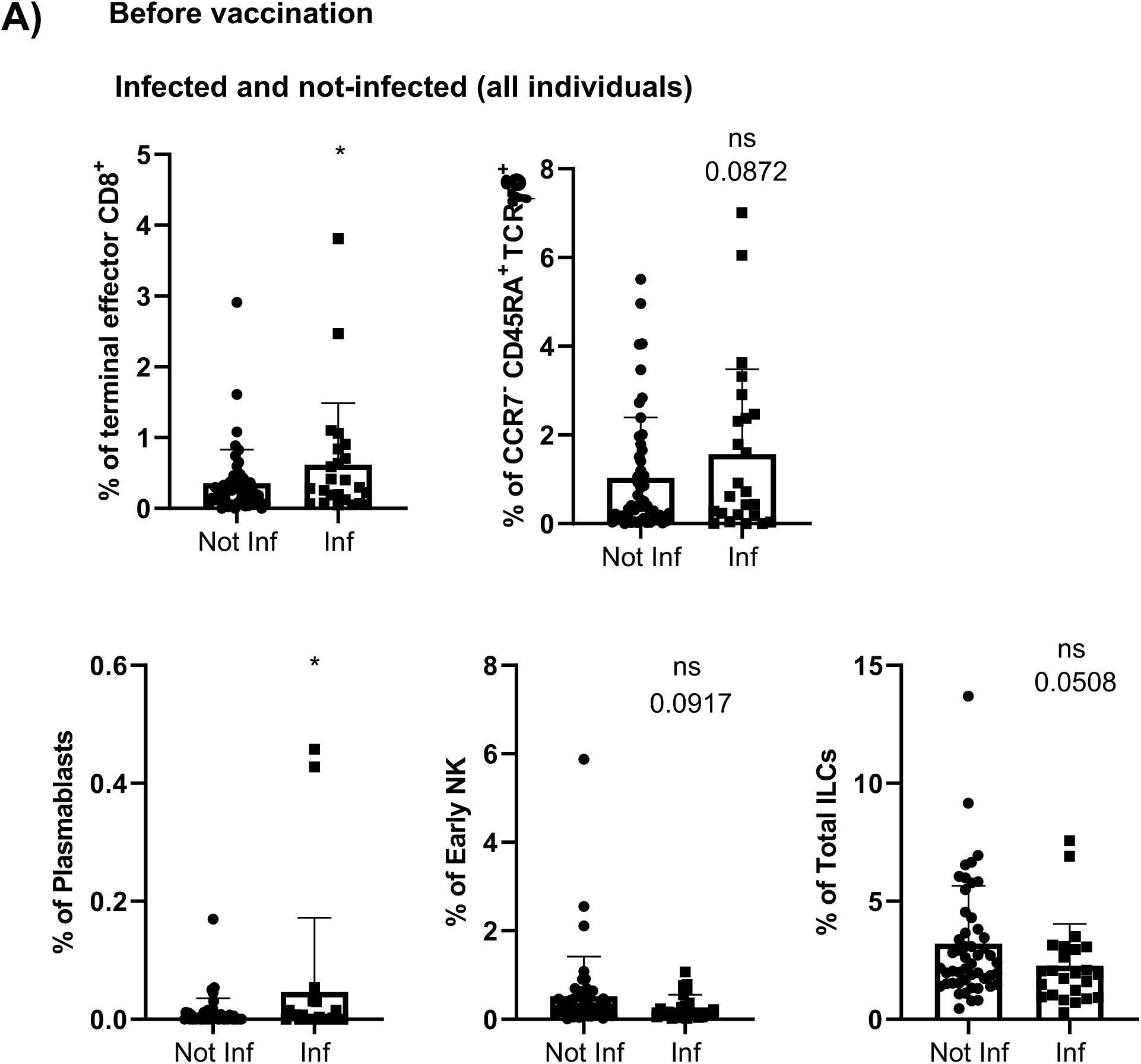
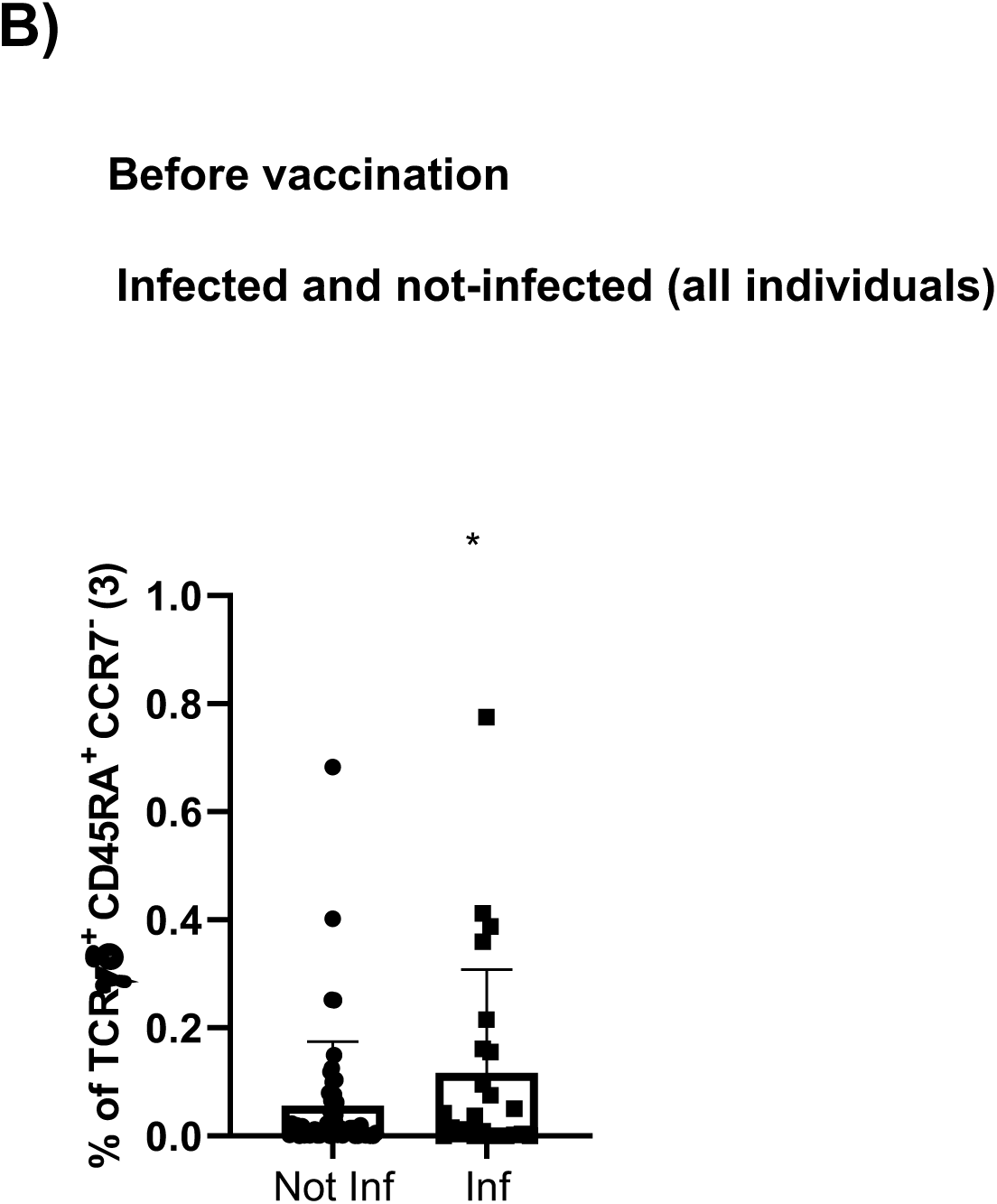
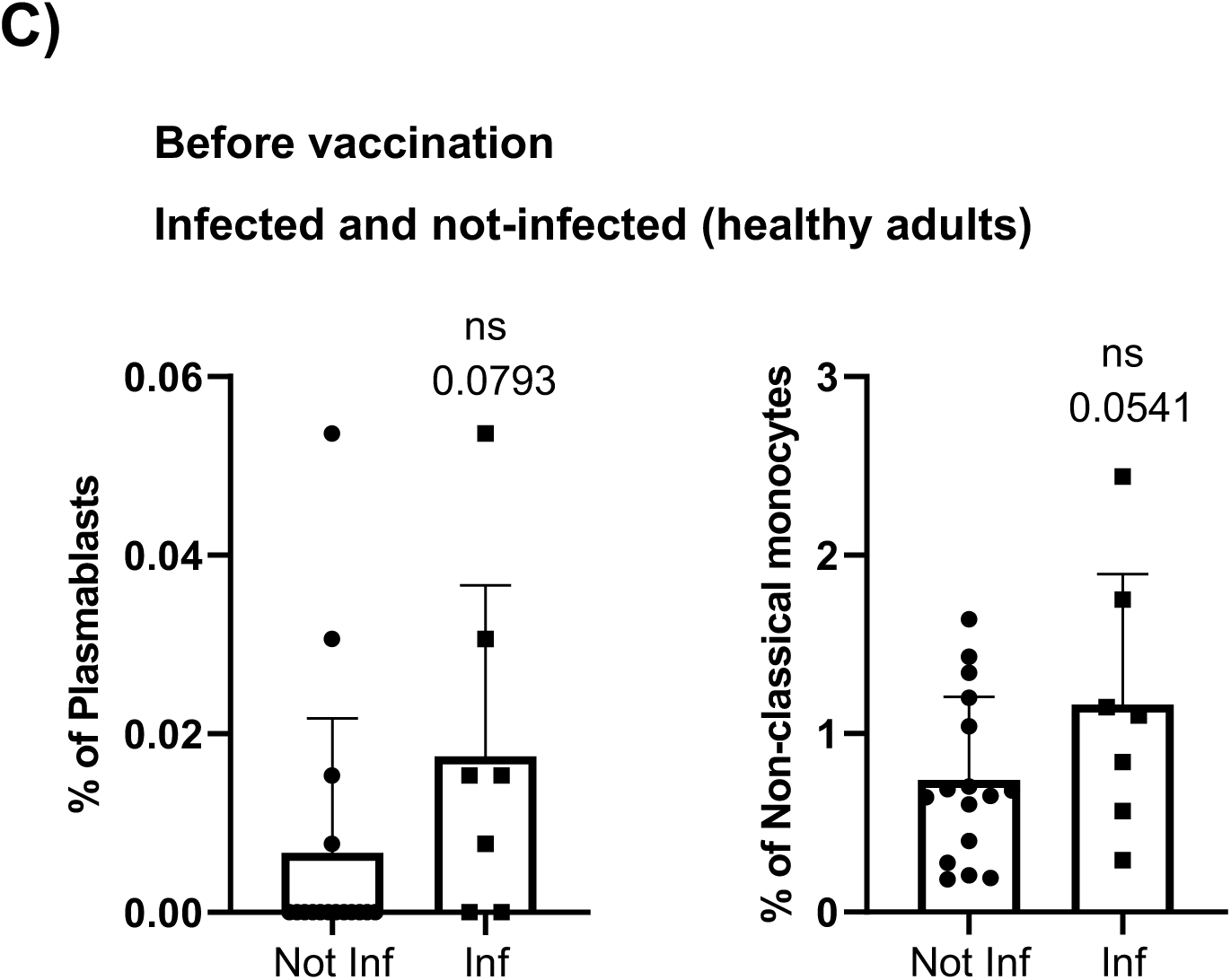
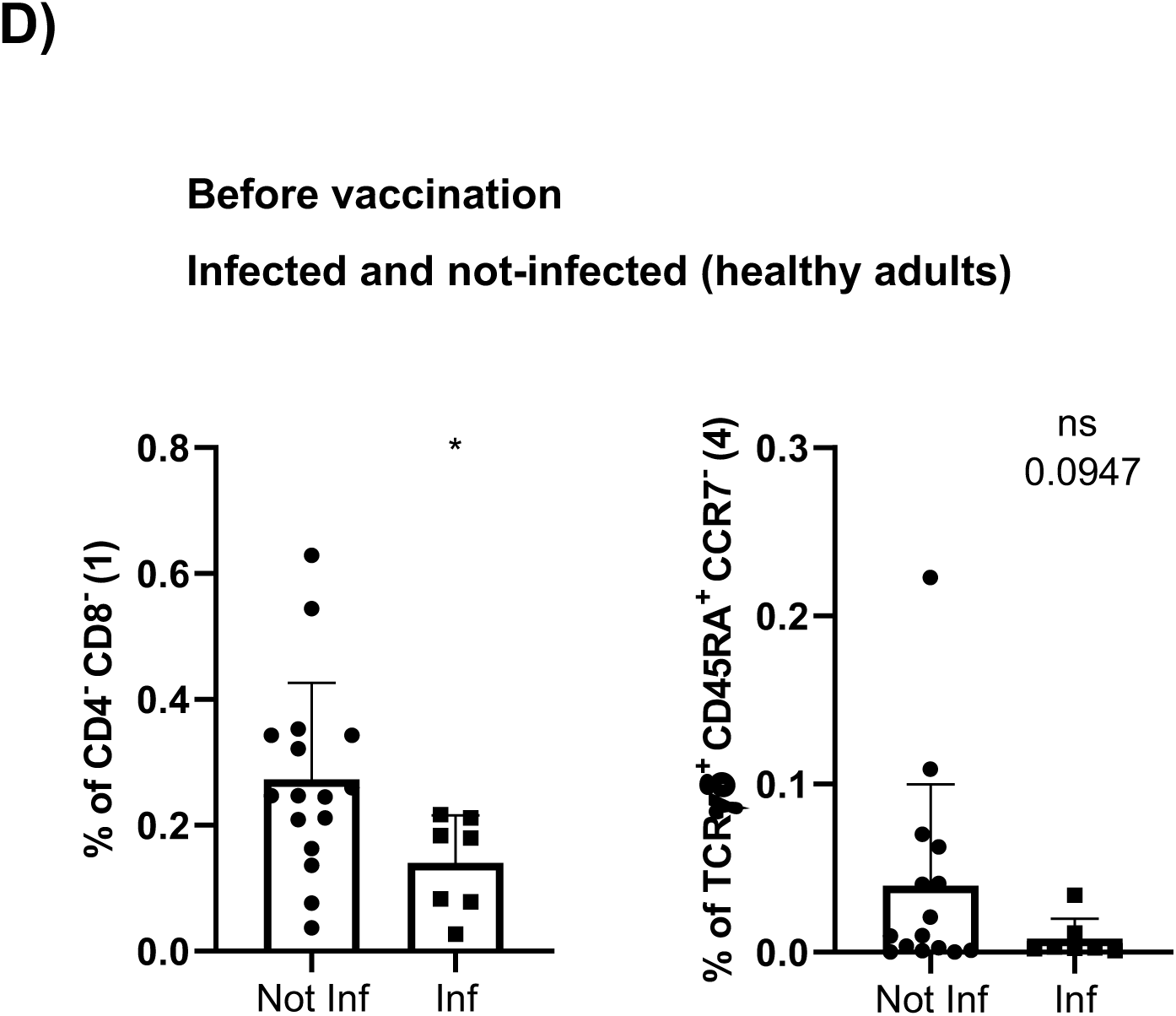
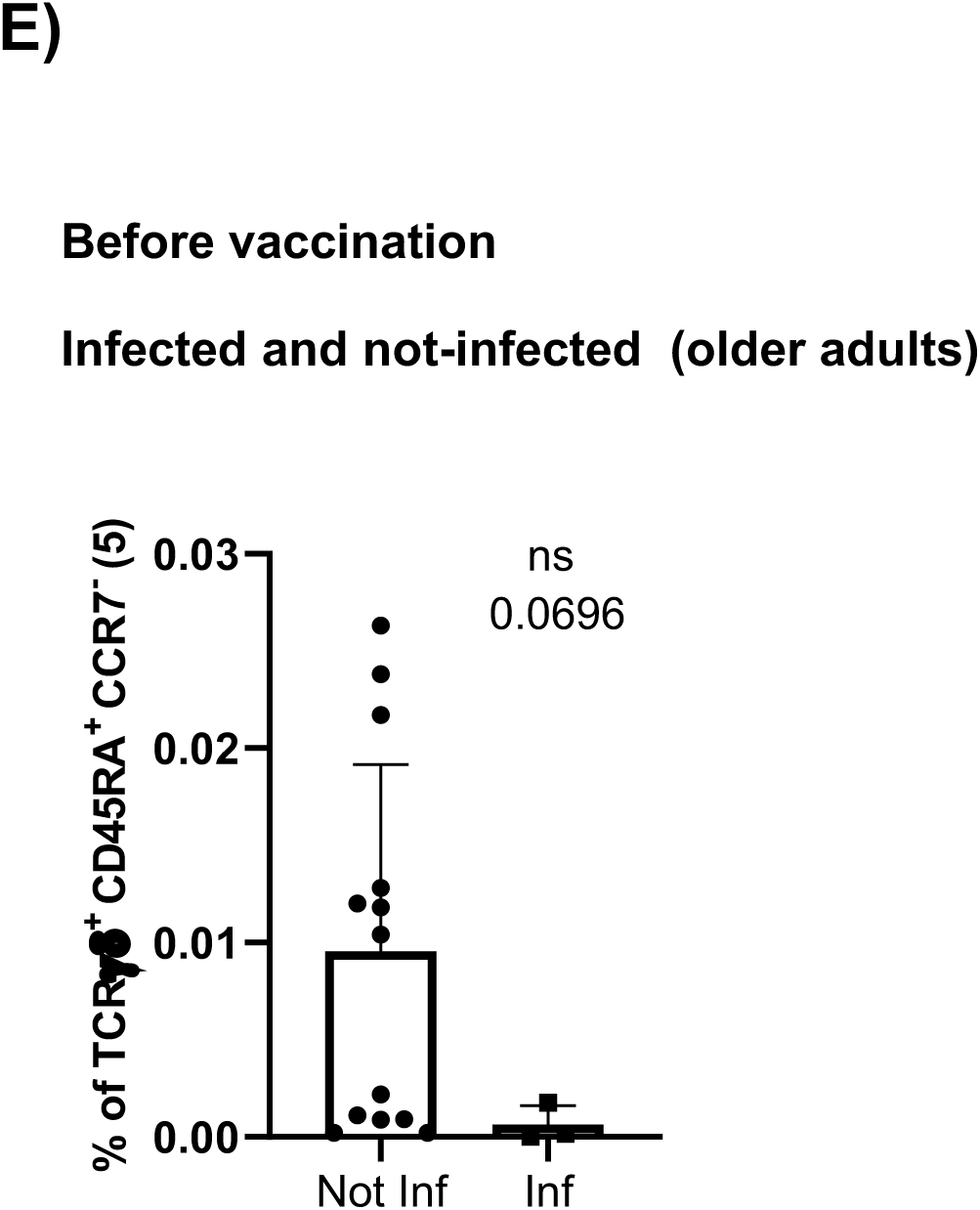
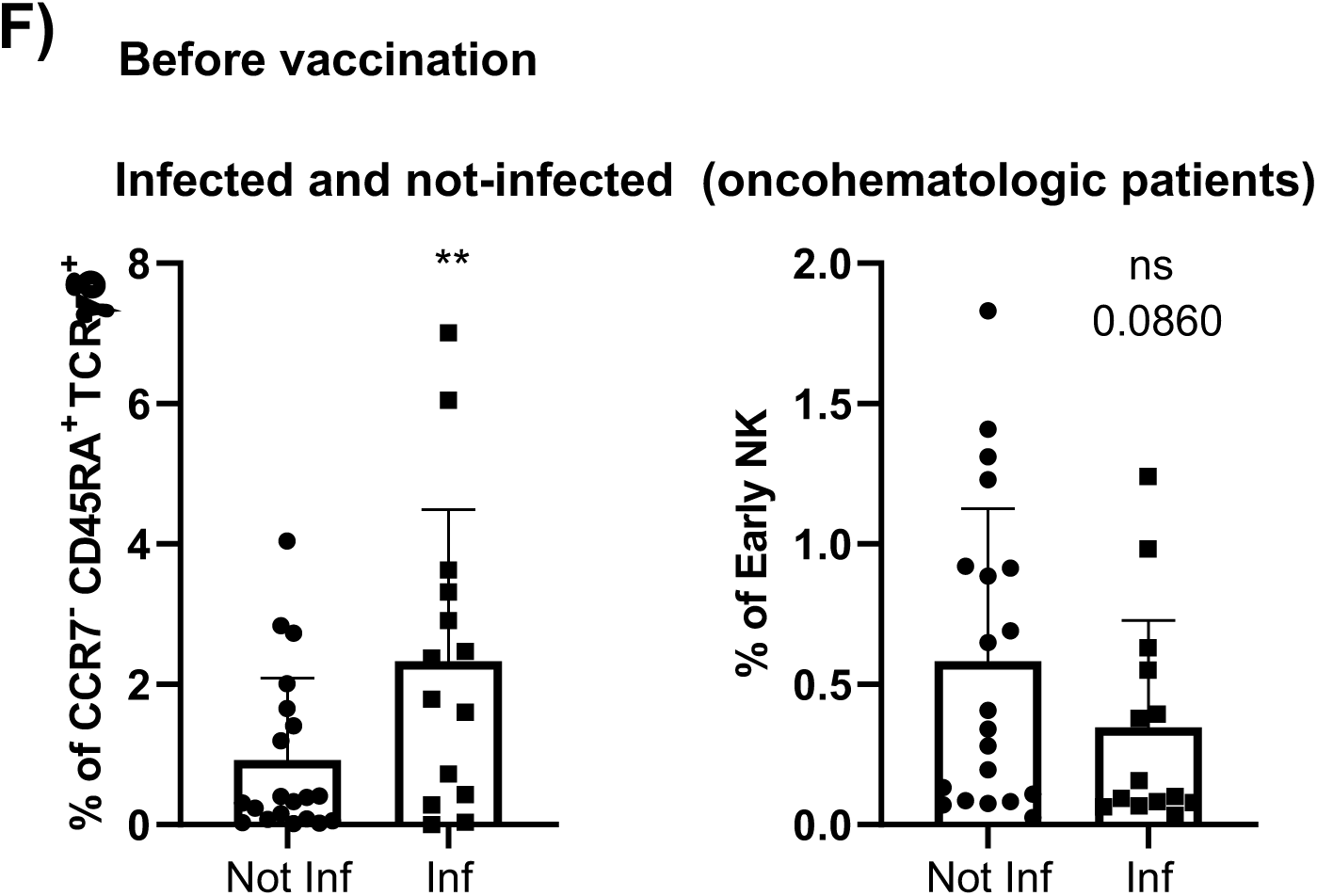
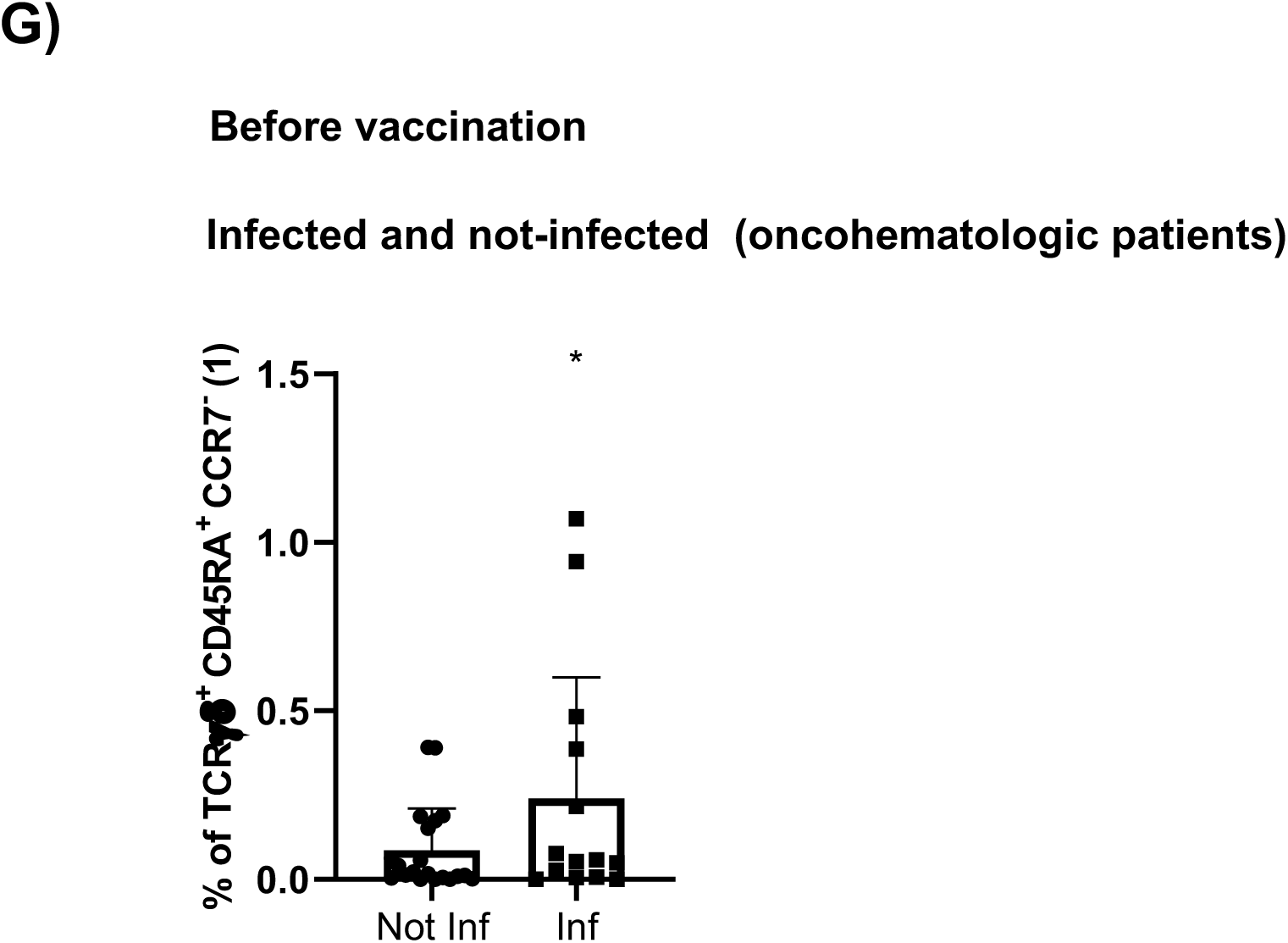
Immunome differences before vaccination predicts SARS-CoV-2 infection. **A)** Classical validation, as in Figure 2, of the populations significantly expressed in the volcano plot from figure 5B comparing the immune systems of all individuals before vaccination based on their subsequent infection following immunization, and **B)** their specific cell cluster subsets. **C)** Comparison of the populations significantly expressed in the volcano plot from figure 5D comparting the immune system of infected and non- infected healthy adults before vaccination, and **D)** their specific cell cluster subsets. **E)** Analysis of the cell cluster subsets differentially expressed in the volcano plot from figure 5E comparting infected and non-infected older adults before vaccination. **F)** Proportion of the populations significantly expressed in the volcano plot from figure 5F comparing infected and non-infected oncohematologic patients before vaccination, and **G)** their specific cluster subsets. In all cases, a t-test analysis was performed where a p- value<0.05 (*<0.05; **<0.01) was considered significant, and a p-value between 0.05 and 0.1 was considered as not significant (ns) but with a relevant trend displaying the p- value underneath. “Inf” refers to subsequently infected individuals, while “Not Inf” are those who were not infected.

